# Viral sequencing reveals US healthcare personnel rarely become infected with SARS-CoV-2 through patient contact

**DOI:** 10.1101/2021.01.28.21250421

**Authors:** Katarina M. Braun, Gage K. Moreno, Ashley Buys, Max Bobholz, Molly A. Accola, Laura Anderson, William M. Rehrauer, David A. Baker, Nasia Safdar, Alexander J. Lepak, David H. O’Connor, Thomas C. Friedrich

## Abstract

**Background:** Healthcare personnel (HCP) are at increased risk of infection with the severe acute respiratory coronavirus 2019 virus (SARS-CoV-2). Between 12 March 2020 and 10 January 2021, >1,170 HCP tested positive for SARS-CoV-2 at a major academic medical institution in the Upper Midwest of the United States. We aimed to understand the sources of infections in HCP and to evaluate the efficacy of infection control procedures used at this institution to protect HCP from healthcare-associated transmission.

**Methods:** In this retrospective case series, we used viral genomics to investigate the likely source of SARS-CoV-2 infection in 96 HCP where epidemiological data alone could not be used to rule out healthcare-associated transmission. We obtained limited epidemiological data through informal interviews and review of the electronic health record. We combined viral sequence data and available epidemiological information to infer the most likely source of HCP infection.

**Findings:** We investigated 32 SARS-CoV-2 infection clusters involving 96 HCP, 140 possible patient contacts, and 1 household contact (total n = 237). Of these, 182 sequences met quality standards and were used for downstream analysis. We found the majority of HCP infections could not be linked to a patient or co-worker and therefore likely occurred in the outside community (58/96; 60.4%). We found a smaller percentage could be traced to a coworker (10/96; 10.4%) or were part of a patient-employee cluster (12/96; 12.5%). Strikingly, the smallest proportion of HCP infections could be clearly traced to a patient source (4/96; 4.2%).

**Interpretation:** Infection control procedures, consistently followed, offer significant protection to HCP caring for COVID-19 patients in a representative American academic medical institution. Rapid SARS-CoV-2 genome sequencing in healthcare settings can be used retrospectively to reconstruct the likely source of HCP infection when epidemiological data are not available or are inconclusive. Understanding the source of SARS-CoV-2 infection can then be used prospectively to adjust and improve infection control practices and guidelines.

**Funding:** This project was funded in part through a COVID-19 Response grant from the Wisconsin Partnership Program at the University of Wisconsin School of Medicine and Public Health to T.C.F. and D.H.O. Author N.S. is supported by the National Institute of Allergy and Infectious Diseases Institute (NIAID) Grant 1DP2AI144244-01.

**Research in context:** *Evidence before this study:* On 16 January 2021 we searched for “SARS-CoV-2” AND “healthcare workers” AND “viral sequencing” in Google Scholar. This search returned 57 results, and included a number of preprint articles. We found two studies that used viral sequencing to investigate healthcare-associated outbreaks in the Netherlands ^1^ and the United Kingdom ^2^. To our knowledge, no study has used viral sequencing to specifically investigate the source of SARS-CoV-2 infections in healthcare workers in the United States. Although we and others have written about the potential utility of sequencing as an infection control asset ^3–6^, few have demonstrated the practical application of such efforts.

*Added value of this study:* Our study suggests infection control measures in place at the institution evaluated in this case series are largely protecting healthcare personnel (HCP) from healthcare-associated SARS-CoV-2 infections. Even so, the majority of healthcare-associated infections we did identify appeared to be linked to HCP-to-HCP spread so additional messaging and guidelines to reduce HCP-to-HCP spread in and out of the workplace may be warranted. In addition, we demonstrated how rapid viral sequencing can be combined with, even limited, epidemiological information to reconstruct healthcare-associated SARS-CoV-2 outbreaks.

*Implications of all the available evidence:* Healthcare-associated SARS-CoV-2 infections negatively affect HCP, patients, and communities. Infections among HCP add further strain to the healthcare system and put patients and other HCP at risk. We found the majority of HCP infections appeared to be acquired through community exposure so measures to reduce community spread are critical. This further emphasizes the importance of mask-wearing, physical distancing, robust testing programs, and the rapid distribution of vaccines.

## Introduction

Despite the use of personal protective equipment (PPE) and other strategies to mitigate risk, front-line healthcare workers are at increased risk for infection with severe acute respiratory syndrome coronavirus 2 (SARS-CoV-2) compared to the general population ^7^. Healthcare-associated transmission of SARS-CoV-2 poses a serious risk to healthcare workers as well as to other hospital staff and patients ^8,9^. Here we use rapid viral sequencing and forensic genomics to uncover the likely source of infection in 96 confirmed cases of coronavirus-disease 2019 (COVID-19) in healthcare personnel (HCP). We further describe how the results of these investigations informed infection control recommendations within a large academic medical system in the midwestern United States.

Healthcare-associated SARS-CoV-2 infections negatively affect HCP through direct health impacts, lost wages, and secondary consequences for their close contacts ^10^. Healthcare-associated SARS-CoV-2 infections can also negatively impact patient care through staffing shortages, environmental contamination, low morale and other mental health impacts on HCP; each of these can secondarily impact the overall quality of care ^2,11^.

The US Centers for Disease Control and Prevention (CDC) have released guidelines for infection prevention for HCP interacting directly with patients with suspected or confirmed infection with SARS-COV-2 ^12^. These guidelines include recommendations for the proper use of PPE, hand hygiene, precautions to be taken during aerosol-generating procedures, collection of diagnostic respiratory specimens, environmental infection control practices and many others. These guidelines in addition to institution-specific infection control measures – described in detail in Lepak et al ^13^ – were in place at the institution evaluated here. We posit that these guidelines are generally successful in protecting HCP from SARS-CoV-2 infection in a healthcare setting. In this report, we test this hypothesis using viral sequencing to investigate the likely sources of infection in a series of HCP in the United States.

Sequencing has been used to explore the origin and the path of spread of various nosocomial *bacterial* pathogens, such as vancomycin resistant-enterococcus (VRE), *Listeria monocytogenes*, and *Klebsiella pneumoniae* ^14–17^. With a few exceptions ^18–20^, viral sequencing is not currently standard practice for investigating healthcare-associated SARS-CoV-2 infections, although we and others have highlighted the potential utility of this approach ^3–6^. Data suggest SARS-CoV-2 accumulates approximately one fixed mutation every other transmission event ^21,22^. Therefore, if SARS-CoV-2 is directly transmitted from one individual to another, this “transmission pair” is expected to share identical, or nearly identical (≤1 consensus SNV difference), viral sequences. The most parsimonious explanation for identical or near-identical genomes generated from individuals with known contact is that they are transmission pairs ^23^. In contrast, two people who became infected at similar times, but from different sources, would be expected to be infected with viruses that differ from each other by two or more nucleotides. This is especially true at this stage of the pandemic in the United States, when transmission rates are high and multiple viruses of distinct genetic lineages cocirculate in many areas ^24^. By increasing the resolution of inference, rapid viral sequencing can facilitate a targeted approach to examine SARS-CoV-2 nosocomial outbreaks at the level of the individual and the institution, which others have referred to collectively as “precision epidemiology” ^25^.

## Results

HCP began testing positive for SARS-CoV-2 at a major academic biomedical institution in the American Upper Midwest in early March 2020. From 12 March, 2020 to 10 January, 2021 ∼1,172 HCP tested positive for SARS-CoV-2 at this center. In collaboration with this institution’s infection control team, we began sequencing viral genomes from residual nasopharyngeal specimens from the individuals involved in these infection clusters. We focused our analyses on HCP infections and infection clusters that were highest risk for nosocomial transmission, as when healthcare-associated transmission could not be ruled out using epidemiological data alone. Each investigation included at least one HCP, all known direct and indirect SARS-CoV-2-positive patient contacts where residual swab was available, and occasionally extended to epidemiologically-linked household contacts. Relevant patient contacts were identified in the Epic electronic medical record using a comprehensive caregiver trace. This function identifies all patient records accessed by a HCP.

We consider three main potential sources of healthcare-associated infection with SARS-CoV-2: “outside community,” “patient source” (via HCP-patient interactions), and “employee source” (via HCP-HCP interactions). A few HCP infections did not fit neatly into these categories so we have included two additional categories. First, “combined patient and employee cluster”, where a patient-to-HCP transmission event likely initiated a cluster of infections, but we were unable to pinpoint the first HCP to become infected. Second, “inconclusive”, where a consensus sequence was not available, there were no appropriate comparator samples, or epidemiological information were insufficient to interpret the sequence data.

For us to conclude person A was a likely source of infection for person B, persons A and B must have had known contact with each other, must have been tested within 0-14 days of each other, and must have been infected with identical or near-identical (≤1 consensus intrahost single nucleotide variant (iSNV) difference) viruses. In cases where we found the HCP virus diverged from the patient- and employee-contact viruses, we concluded infection most likely occurred outside of the healthcare setting, or in the “outside community”.

In total, we investigated 32 SARS-CoV-2 infection clusters involving 96 HCP, 140 possible patient contacts, and 1 household contact (total n = 237). This accounted for approximately ∼8.2% (96/1,172) of all documented infections in the HCP at this institution. In total, we sequenced 237 samples collected between 15 March and 27 December, 2020. Of these, 182 met quality standards (as described in methods) and were used for downstream analysis. We did not find a closely related virus among the epidemiologically linked contacts in 58 HCP infections, so we concluded the most likely source of infection in these cases could be traced to the outside community (58/96; 60.4%). We find a smaller percentage could be traced to a coworker (10/96; 10.4%) or were part of a patient-employee cluster (12/96; 12.5%). Strikingly, the smallest proportion of HCP infections could be clearly traced to a patient source (4/96; 4.2%). The remaining HCP infections could not be definitively traced to a single source and were therefore inconclusive (12/96; 12.5%) (**Table 1**). Below, we describe one representative example of three distinct transmission scenarios – outside community-to-HCP, HCP-to-HCP, and patient-to-HCP. A brief overview of all cases included in this study can be found in **Supplementary File 1**.

**Table 1.**
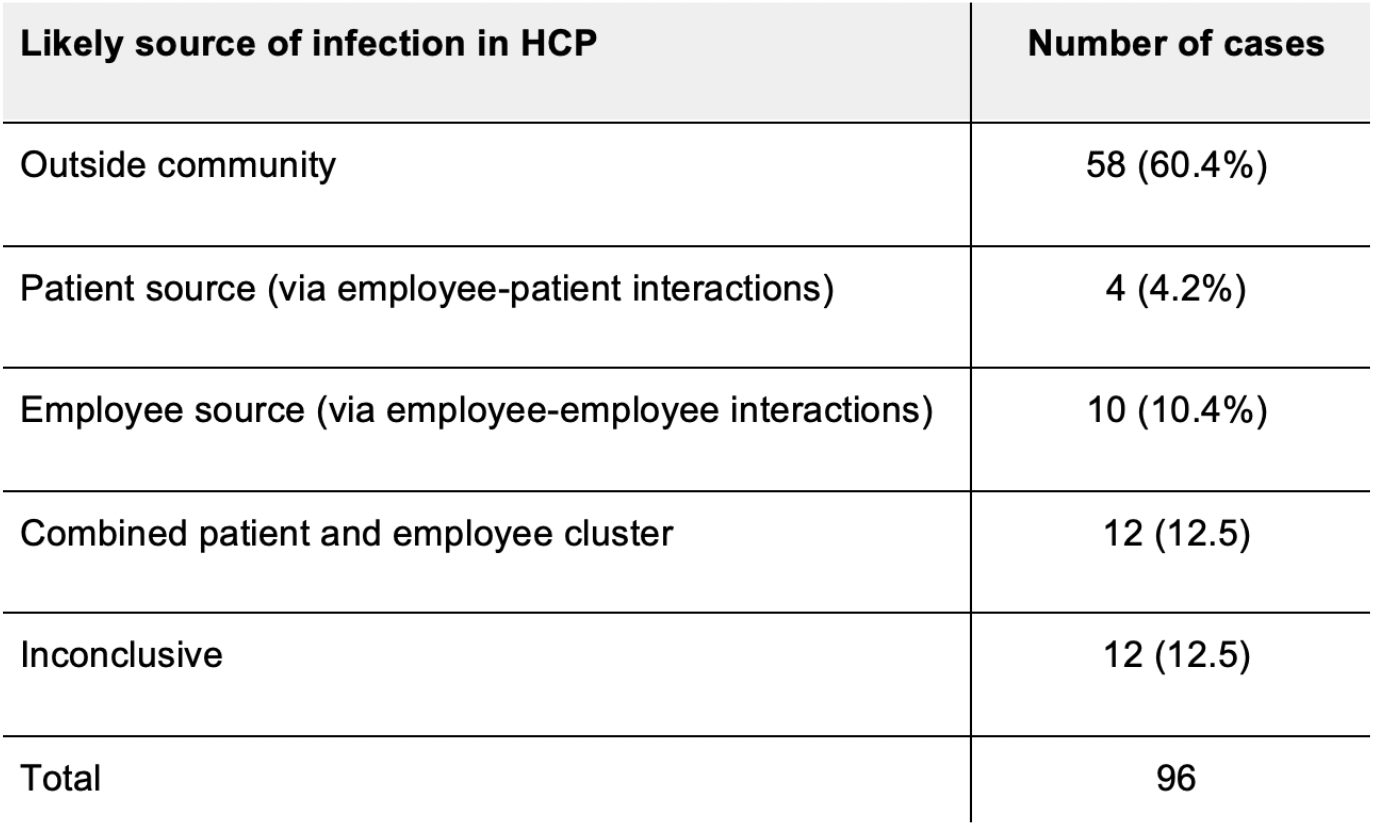
Summary of the likely source of infection in the HCP evaluated in this study.

In case #20, we compared the viral sequence of a HCP (HCP 20-1), who tested positive in early October, to a patient contact who tested positive eight days prior. A comprehensive caregiver trace of HCP 20-1 revealed a single patient contact with diagnosed COVID-19 (patient 20-A) within the 14 days prior HCP 20-1’s symptom onset. HCP 20-1 provided direct care to patient 20-A while wearing appropriate PPE and with no reported lapses in PPE. Viral sequencing revealed HCP 20-1 was infected with a virus clustering with the 20G clade whereas patient 20-A was infected with a 20A-clade virus. The sequences of these viruses differed at >20 sites, so we concluded these individuals were unlikely to represent a transmission pair and HCP 20-1 was more likely infected in the outside community (**Figure 1**).

**Figure 1.**
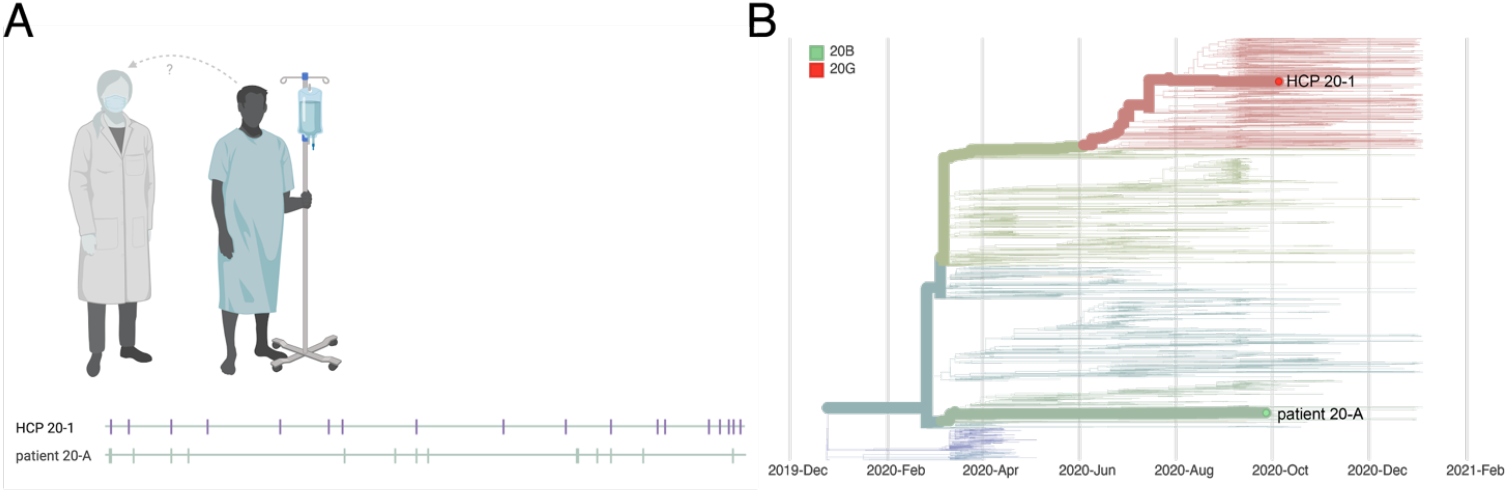
Graphical representation of case #20. A. Virus sequences are aligned against SARS-CoV-2 reference sequence Wuhan-Hu-1 (MN908947.3). Vertical markers denote the location of consensus nucleotide differences between patient viruses and the reference. B. A time-resolved phylogenetic tree built using Nextstrain tools with all Wisconsin sequences available as of 2021-01-15. Viruses involved in this case are denoted with thick branches and labeled tips. Color denotes clade.

In case #16, we investigated infections in three HCP who worked in the same department and tested positive in early September (HCP 16-2), mid-September (HCP 16-1), and late September (HCP 16-3). Contact tracing revealed HCP 16-2 worked for two days prior to symptom onset and may have had unmasked contact with HCP 16-1 during overlapping meal breaks. Contact tracing additionally revealed HCP 16-3 had an exposure event lasting >15 minutes in the outside community prior to testing positive. Viral sequencing in this cluster showed HCP 16-1 and 16-2 were infected with 20G-clade viruses that were identical at the consensus level, while HCP 16-3 was infected with a genetically dissimilar 20A-clade virus. We therefore concluded HCP 16-2 was a likely source of infection for HCP 16-1, while HCP 16-3 was likely infected elsewhere (**Figure 2**).

**Figure 2.**
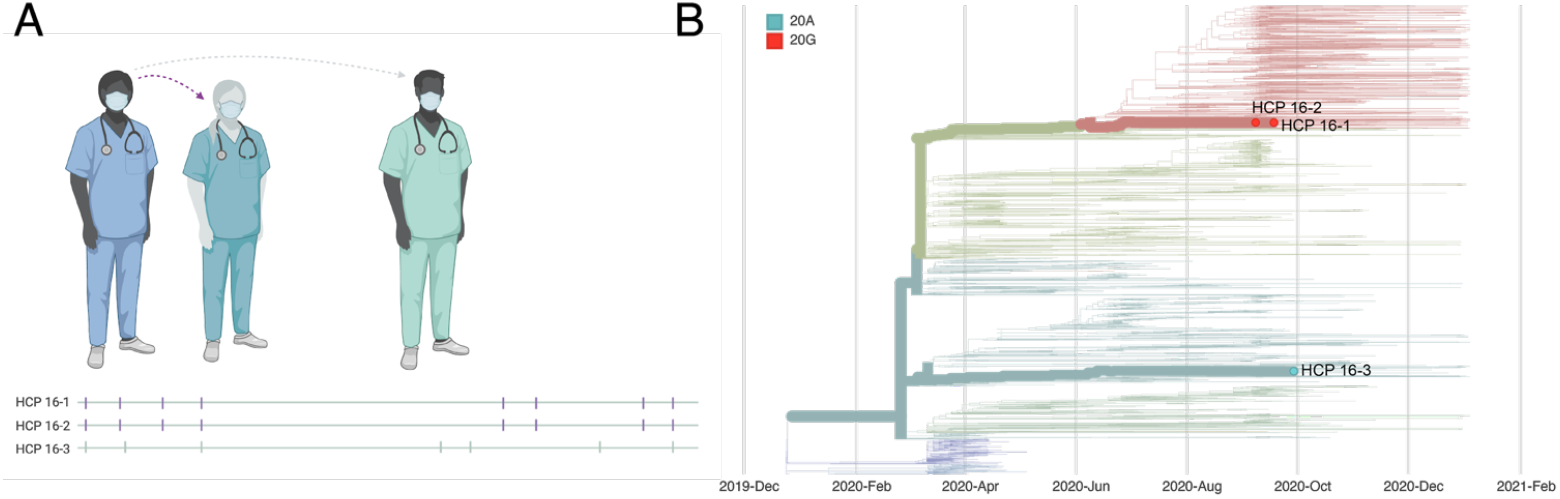
Graphical representation of case #16. A. Virus sequences are aligned against SARS-CoV-2 reference sequence Wuhan-Hu-1 (MN908947.3). Vertical markers denote the location of consensus nucleotide differences between patient viruses and the reference. Purple vertical markers indicate identical virus sequences. B. A time-resolved phylogenetic tree built using Nextstrain tools with all Wisconsin sequences available as of 2021-01-15. Viruses involved in this case are denoted with thick branches and labeled tips. Color denotes clade.

Case #10 involved a HCP (HCP 10-1) who provided care for 15 patients diagnosed with COVID-19 in the 14 days prior to HCP 10-1’s symptom onset. HCP 10-1 provided direct care to each of these patients while wearing appropriate PPE with no reported lapses in PPE. We generated consensus sequences from HCP 10-1 and nine patient contacts. There was insufficient viral RNA (vRNA) in the remaining six patient contacts to generate high-quality consensus sequences for comparison. The virus isolated from patient 10-G was identical to the virus from HCP 10-1. Given the known epidemiological association between these two individuals (HCP 10-1 provided direct patient care to patient 10-G), the time separating sample collections (late July & early August), and identical viral sequences, we concluded patient 10-G is a likely source of infection for HCP 10-1 (**Figure 3**).

**Figure 3.**
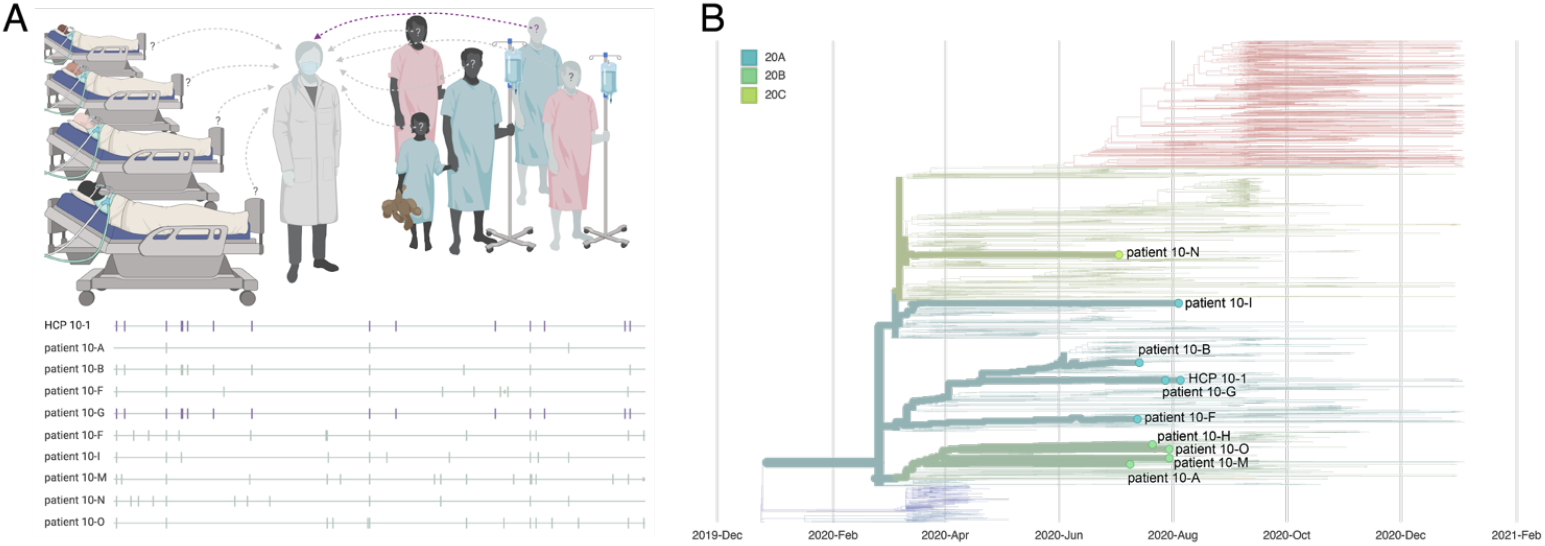
Graphical representation of case #10. A. Virus sequences are aligned against SARS-CoV-2 reference sequence Wuhan-Hu-1 (MN908947.3). Vertical markers denote the location of consensus nucleotide differences between patient viruses and the reference. Purple vertical markers indicate identical virus sequences. B. A time-resolved phylogenetic tree built using Nextstrain tools with all Wisconsin sequences available as of 2021-01-15. Viruses involved in this case are denoted with thick branches and labeled tips. Color denotes clade.

The center where we conducted this case series implemented a number of changes to their institutional infection control guidelines based on these sequencing results ^13^. The recommendations for extended reuse of medical grade face masks were clarified and now instruct HCP to consider barrier mask replacement after three days of wear, to inspect the barrier mask prior to each use and to replace if soiled or damaged, and to always ask for PPE when needed. N-95s or powered air-purifying respirators (PAPR) are now universally required on inpatient units housing COVID-19-confirmed and suspected patients. In addition, medical-grade face masks, instead of cloth masks, are now required for HCP in all clinical areas, and not just direct patient care areas. This final recommendation was based on likely HCP-to-HCP transmission involving a HCP who was not directly involved in patient care of COVID-19 patients (case #14 in **Supplementary File 1**).

## Discussion

HCP across the hospital are involved in caring for people with COVID-19, whether or not they work on an actual COVID-19 ward. With shifting guidelines and PPE shortages that persist today, it is critical to assess the risk that HCP treating people with known SARS-CoV-2 infection will become infected themselves. Here we used viral genome sequencing to assess the risk that HCP in a large academic medical system treating COVID-19 patients would acquire nosocomial infections. Although others have written about the potential utility of sequencing as an infection control asset within clinical settings, few have demonstrated the practical application of such efforts ^3–5^. Our results suggest that caring for COVID-19 patients accounted for a minority of HCP infections (n=4), even while evaluating high-risk infections where epidemiological data alone could not be used to rule out healthcare-associated transmission. In contrast, HCP at this institution were much more likely to acquire SARS-CoV-2 from infected coworkers (n=10) or in the outside community (n=58). This result suggests that infection control procedures, consistently followed, offer significant protection to HCP caring for COVID-19 patients in the United States. A similar conclusion was drawn by recent studies evaluating healthcare-associated infections in the Netherlands and in the UK, suggesting this conclusion may hold across healthcare systems ^1,2^.

Our results suggest that the infection control measures and PPE in place at the institution where we conducted this case series ^13^ can protect against symptomatic SARS-CoV-2 infection, even in settings where the density of individuals infected with SARS-CoV-2 is relatively high. This provides some confidence that PPE and similar infection control measures, when used consistently and correctly, will provide adequate protection in other congregate settings, like schools. The impact of reopening primary schools remains unclear, yet the long-term consequences of remote learning and physical distancing on children, particularly those from disadvantaged backgrounds, are of increasing concern ^26^. In addition to reducing rates of community spread, it follows that schools should prioritize making PPE and training on its appropriate use available to all staff ^27^. Similar precautions are needed in other congregate settings as well, such as correctional facilities and factories, to protect inmates and workers.

The effectiveness of hospital- and clinic-like PPE and infection control guidelines used in other congregate settings should similarly be evaluated following implementation. As the cost of viral sequencing continues to drop and sequencing technologies continue to improve, the integration of rapid pathogen sequence information with traditional epidemiological investigation approaches is possible at the level of individual hospitals, clinics, school systems, and correctional facilities. Such systems would allow for the rapid reconstruction of transmission networks. These systems would also inform feedback loops for schools and institutions to continually assess and improve the efficacy of infection control and PPE procedures in protecting HCP, teachers, children, workers, and individuals who are incarcerated. We should work to build such systems now because we can be certain pathogenic viruses will continue to emerge unpredictably.

Sampling and contact tracing of nosocomial outbreaks is often coordinated by local hospitals and/or departments of health while expertise in viral sequencing, bioinformatics, and phylogenetics can more often be found in academic laboratories. Successful application of precision epidemiology requires the integration of these areas. This is possible now at academic medical institutions like ours, but presents more of a challenge at smaller, rural, and private patient care centers. Federal support should be provided to help establish and maintain these collaborations in the current pandemic and in anticipation of future outbreaks.

Importantly, we were only able to evaluate samples for which residual swab was available and for which we were able to generate high-quality consensus sequences. Given this limitation, we were often able to exclude patient contacts and co-workers as likely sources of infection in HCP, but we were rarely able to pinpoint the exact source of infection, especially when it occurred outside of the healthcare setting. In addition, this study only examined SARS-CoV-2 infections in HCP from a single academic medical center so our conclusions may not be broadly generalizable. However, another recent study evaluated healthcare-associated infections in the Netherlands and similarly found no evidence for widespread nosocomial transmission of SARS-CoV-2, suggesting our conclusions may hold across institutions and healthcare systems ^1^.

Further, we were not able to differentiate between routes of infection (airborne, droplet, contact) with the limited epidemiological data available to us in this study.

Here we demonstrated how rapid whole-genome sequencing of current SARS-CoV-2 outbreaks in hospitals can be used retrospectively to reconstruct the likely source of HCP infection and prospectively to adjust and improve infection control practices and guidelines. The approach we describe here need not be limited to investigation of pandemic virus outbreaks. Key concepts from genome sequencing and routine pathogen surveillance can be applied to any nosocomial pathogen and inform changes to infection control practices. Overall, while we do find examples of patient-to-HCP and HCP-to-HCP spread, the majority of HCP infections appeared to be acquired through community exposure, emphasizing the importance of ongoing measures to reduce community spread through mask-wearing, physical distancing, robust testing programs, and rapid distribution of vaccines.

## Methods

### Sample approvals and sample selection criteria

From 12 March 2020 to 10 January 2021, ∼1,172 HCP tested positive for SARS-CoV-2 at a major academic medical institution in the Upper Midwest. In the case of infections where epidemiological data alone could not rule out healthcare-associated infection, we sequenced viral genomes from all available residual specimens from the individuals involved in these outbreaks. These clusters included at least one HCP, SARS-CoV-2-positive patient contacts, and occasionally extended to epidemiologically-linked hospital staff and household contacts. Relevant patient contacts were identified in the Epic electronic medical record using a comprehensive caregiver trace. This function identifies all patient records accessed by a HCP being traced.

### Summary of infection control measures to prevent transmission of SARS-CoV-2 at our institution

Detailed descriptions of all infection control measures implemented to prevent transmission of SARS-CoV-2 at the medical institution evaluated here can be found in a recent report by Lepak et al ^13^. Briefly, these guidelines include a universal testing policy for all patients admitted to the hospital, negative air pressure in all locations where SARS-CoV-2 patients are treated, a limit of one visitor or primary support person per patient per day (required to undergo screening prior to entry), establishment of an employee testing site with required employee self-monitoring for signs and symptoms of infection, maintenance of a log of persons entering the room of a confirmed or suspected COVID-19 patient for contact tracing purposes, and establishment of a designated respiratory care unit in the emergency department, among others. PPE guidelines include universal masking and face-shield use with any patient care contact, enhanced PPE requirements for HCP working in the intensive care unit or performing aerosol-generating procedures, and formal donning and doffing protocols and required training for all HCP, among others.

### vRNA isolation

All samples were isolated using a Maxwell isolation instrument and subsequently processed using a modified ARTIC tiled amplicon approach ^28,29^. Nasopharyngeal swabs received in transport medium (VTM) were briefly centrifuged at 14,000 r.p.m. for 30 seconds at room temperature to ensure all residual sample sediments at the bottom of the tube. Viral RNA (vRNA) was extracted from 100μl of VTM using the Viral Total Nucleic Acid Purification kit (Promega, Madison, WI, USA) on a Maxwell RSC 48 instrument and was eluted in 50 μL of nuclease-free H_2_O.

### Complementary DNA (cDNA) generation

Complementary DNA (cDNA) was synthesized using a modified ARTIC Network approach ^28,29^. Briefly, vRNA was reverse transcribed with SuperScript IV Reverse Transcriptase (Invitrogen, Carlsbad, CA, USA) using random hexamers and dNTPs. Reaction conditions were as follows: 1μL of random hexamers and 1µL of dNTPs were added to 11 μL of sample RNA, heated to 65°C for 5 minutes, then cooled to 4°C for 1 minute. Then 7 μL of a master mix (4 μL 5x RT buffer, 1 μL 0.1M DTT, 1µL RNaseOUT RNase Inhibitor, and 1 μL SSIV RT) was added and incubated at 42°C for 10 minutes, 70°C for 10 minutes, and then 4°C for 1 minute.

### Multiplex PCR to generate SARS-CoV-2 genomes

A SARS-CoV-2-specific multiplex PCR for Nanopore sequencing was performed, similar to amplicon-based approaches as previously described ^28,29^. In short, primers for 96 overlapping amplicons spanning the entire genome with amplicon lengths of 500bp and overlapping by 75 to 100bp between the different amplicons were used to generate cDNA. cDNA (2.5μL) was amplified in two multiplexed PCR reactions using Q5 Hot-Start DNA High-fidelity Polymerase (New England Biolabs, Ipswich, MA, USA) using the conditions previously described ^28,29^. Samples were amplified through 25 cycles of PCR and each resulting multiplex sample was pooled together before ONT library prep.

### Library preparation and sequencing

Amplified PCR product was purified using a 1:1 concentration of AMPure XP beads (Beckman Coulter, Brea, CA, USA) and eluted in 30μL of water. PCR products were quantified using Qubit dsDNA high-sensitivity kit (Invitrogen, USA) and were diluted to a final concentration of 1 ng/μl. A total of 5ng for each sample was then made compatible for deep sequencing using the one-pot native ligation protocol with Oxford Nanopore kit SQK-LSK109 and its Native Barcodes (EXP-NBD104 and EXP-NBD114) ^29^. Specifically, samples were end-repaired using the NEBNext Ultra II End Repair/dA-Tailing Module (New England Biolabs, Ipswich, MA, USA).

Samples were then barcoded using 2.5µL of ONT Native Barcodes and the Ultra II End Repair Module. After barcoding, samples were pooled directly into a 1:1 concentration of AMPure XP beads (Beckman Coulter, Brea, CA, USA) and eluted in 30µL of water. Samples were then tagged with ONT sequencing adaptors according to the modified one-pot ligation protocol ^29^. Up to 24 samples were pooled prior to being run on the appropriate flow cell (FLO-MIN106) using the 72hr run script.

### Processing raw ONT data

Sequencing data was processed using the ARTIC bioinformatics pipeline (https://github.com/artic-network/artic-ncov2019), with a few modifications. Briefly, we have modified the ARTIC pipeline so that it demultiplexes raw fastq files using qcat as each fastq file is generated by the GridION (https://github.com/nanoporetech/qcat). Once a barcode reaches 100k reads, it will trigger the rest of the ARTIC bioinformatics workflow which will map to the Severe acute respiratory syndrome coronavirus 2 isolate Wuhan-Hu-1 reference (Genbank: MN908947.3) using minimap2. This alignment will then be used to generate consensus sequences and variant calls using medaka (https://github.com/nanoporetech/medaka). The entire ONT analysis pipeline is available at https://github.com/gagekmoreno/SARS-CoV-2-in-Southern-Wisconsin.

### Consensus sequence analysis – clade and lineage generation

Following the generation of consensus sequences via the above ARTIC pipeline, samples were excluded from downstream analysis if gaps in the consensus sequence totaled ≥20% of the genome. Each sample’s consensus sequence within a report (or as many as possible), were visually inspected in Geneious Prime (https://www.geneious.com) and/or in Nextstrain’s Nextclade online tool (https://clades.nextstrain.org/). When inspecting in Geneious, we aligned all sequences to a standard reference sequence – an early Wuhan sequence (Genbank: MN908947.3). Using these alignments as well as exported Nextclade CSV files, we were able to quickly identify consensus intrahost single nucleotide variant (iSNV) differences. We additionally used Nextstrain’s Nextclade tool to assign clades. We used Pangolin’s command-line tool to assign sequences to Pangolin lineages (https://github.com/cov-lineages/pangolin).

As noted in the results, for us to conclude person A was a likely source of infection for person B, persons A and B must have had known contact or epidemiological links, they must have been tested within 0-14 days of each other, and they must be infected with identical or near-identical (≤1 consensus iSNV difference) viruses.

### Consensus sequence analysis – Southeast Wisconsin Phylogenetic tree

Wisconsin-centric time-resolved and divergence phylogenetic trees (seen in **Supplementary File 1**) were built using the standard Nextstrain tools and scripts ^30^.

### Report generation

For each sample set, we drafted these results into a PDF report to be shared with the hospital infection control team. These reports were drafted in a standardized format. In each report, we outlined the purpose for the report, the samples involved including any associated metadata, a brief description of the methods used to generate the sequence data, a screen grab of a Nextclade alignment, a table summarizing the iSNV differences, phylogenetic trees if applicable, and finally any overall conclusions that could be drawn regarding the likely and unlikely sources of infection. A summary of each of these reports can be found in **Supplementary File 1**.

### Data availability

Variant identities, alignments, phylogenetic trees, Nextclade CSVs, and Pangolin lineage reports can be found for each case on the GitHub accompanying this manuscript <u>here</u>. An interactive view of the Wisconsin Nextstrain phylogenetic tree can be found <u>here</u>. Figures 1A, 2A, and 3A were created with <u>BioRender.com</u>.

### Study approvals

The University of Wisconsin-Madison Institutional Review Board deemed this study quality improvement, rather than research, and considered it exempt from review. Data and metadata were collected as part of routine infection control policy in nosocomial outbreaks and all data were deidentified prior to analysis.

## Data Availability

Variant identities, alignments, phylogenetic trees, Nextclade CSVs, and Pangolin lineage reports can be found for each case on the GitHub accompanying this manuscript here (https://github.com/katarinabraun/SARSCoV2_sequencing_healthcare-association_infections). An interactive view of the Wisconsin Nextstrain phylogenetic tree can be found here (https://nextstrain.org/community/gagekmoreno/Wisconsin-SARS-CoV-2/ncov/wisconsin/2021-1-8?c=clade_membership&r=location). Figures 1A, 2A, and 3A were created with BioRender.com.

## Acknowledgements

We gratefully acknowledge Anna Heffron for assisting with sample transport. We also thank all healthcare workers and infection control teams for their ongoing dedication to patient and community health and wellness. This project was funded in part through COVID-19 Response grants from the Wisconsin Partnership Program at the University of Wisconsin School of Medicine and Public Health to T.C.F. and D.H.O. N.S. is supported by the National Institute of Allergy and Infectious Diseases Institute (NIAID) Grant 1DP2AI144244-01.

