## Supplementary_File_1 for "Viral sequencing reveals US healthcare personnel rarely become infected with SARS-CoV-2 through patient contact"

### Infection control report summary

Supplementary File 1

#### Likely sources of infection in HCP: definitions

**Outside community**. Among the sequences available for comparison, the likely source of infection was not a patient and was not a co-worker/employee.

**Patient source**. The most likely source of infection in the HCP was a patient source.

**Employee source**. The most likely source of infection in the HCP was a co-worker/employee.

**Combined patient and employee cluster**. A patient to HCP transmission event likely started this cluster and was followed by HCP-to-HCP transmission. However, we are unable to pinpoint the first HCP to become infected and/or are unable to distinguish ongoing sources of transmission as patient-to-HCP and HCP-to-HCP are both possible.

**Inconclusive**. No consensus sequence available, and/or there were no appropriate comparator sequences available, and/or epidemiological information were insufficient to interpret sequence data

#### Summary table

| Likely source of infection in HCP | Number of cases |
|---|---|
| Outside community | 58 (60.4%) |
| Patient source (via employee-patient interactions) | 4 (4.2%) |
| Employee source (via employee-employee interactions) | 10 (10.4%) |
| Combined patient and employee cluster | 12 (12.5) |
| Inconclusive | 12 (12.5) |
| Total | 96 |

Total number of patient comparator samples = 140 (96 consensus sequences).

Each case included in the above table is summarized below. For each case, we include the likely source of infection for all involved healthcare personnel (HCP). Next, we provide essential information for each associated sample in the form of a table, including sample collection date, GISAID identifier, Nextstrain clade, and Pangolin lineage. We report clades using the updated Nextstrain clade naming strategy as outlined by Bedford, Hodcroft, and Neher in Virological.org [1]. We report lineages according to the Pangolin nomenclature as outlined by Rambaut and colleagues [3]. A description of each Pangolin lineage can be found at cov-lineages [2]. Next, we provide a very brief overview of any known epidemiological interactions among the involved individuals. The level of epidemiological detail associated with each case is variable, but we have included all known information here. We include a simple alignment showing the consensus sequences mapped against the Wuhan-Hu-1 reference sequence. Consensus-level differences amongst the reference and the sample sequences are denoted with a vertical black line. Particular variant identities for each sample can be found on the GitHub accompanying this manuscript. Finally, we include a time-resolved phylogenetic tree, built using Nextstrain algorithms, for each case. These trees include all sequences which are publicly available in the GISAID database from the state of Wisconsin. We highlight the samples involved in each case using bolded branches and nodes. An interactive view of this tree can be found here.

[1] https://virological.org/t/updated-nextstain-sars-cov-2-clade-naming-strategy/581
[2] https://cov-lineages.org/lineages.html
[3] Rambaut A, Holmes EC, O'Toole Á, Hill V, McCrone JT, Ruis C, du Plessis L, Pybus OG. A dynamic nomenclature proposal for SARS-CoV-2 lineages to assist genomic epidemiology. Nat Microbiol. 2020 Nov;5(11):1403-1407. doi: 10.1038/s41564-020-0770-5. Epub 2020 Jul 15. PMID: 32669681.

### Report #1. 2020-04-15.

#### Likely source of HCP infection

HCP 1. Outside community.

#### Samples

| Sample type | Sample collection date | GISAID identifier | Clade (Nextstrain) | Lineage (Pangolin) |
|---|---|---|---|---|
| HCP 1 | April 2020 | hCoV-19/USA/IA-UW-121/2020 | 20A | B.1 |
| patient A | April 2020 | hCoV-19/USA/WI-UW-120/2020 | 20A | B.1.19 |
| patient B | April 2020 | hCoV-19/USA/WI-UW-122/2020 | 20A | B.1.19 |

#### Epidemiological information

In the 14 days before symptom onset, HCP 1 interacted with patients A and B per comprehensive caregiver trace (see Methods, “Sample approvals and sample selection criteria” for further information). HCP 1 reported wearing appropriate personal protective equipment (PPE) while providing care to these patients.

#### Alignment

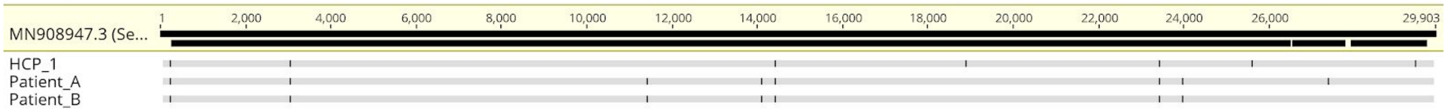

#### Phylogeny

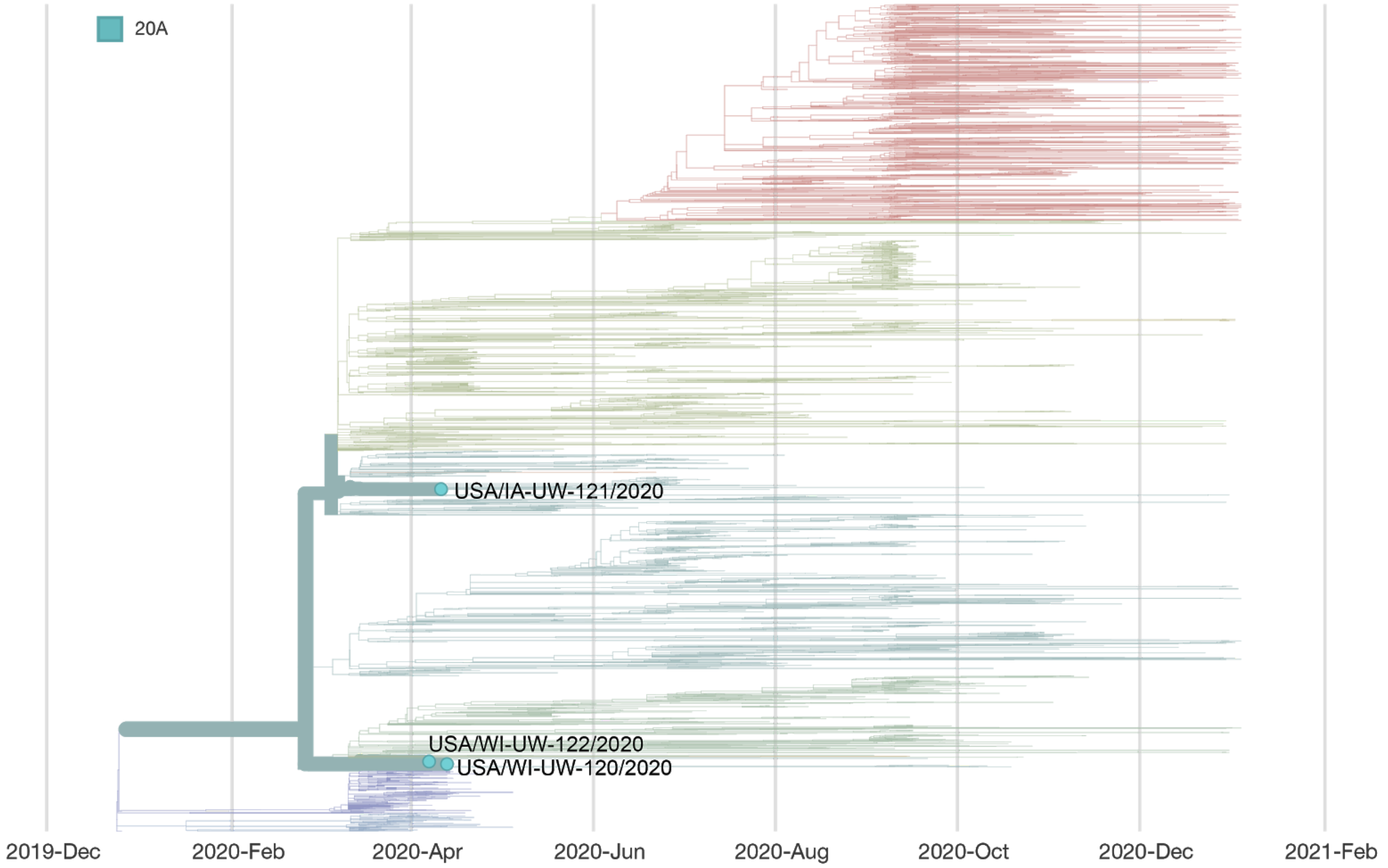


### Report #2. 2020-04-16.

#### Likely source of HCP infection

HCP 1. Outside community (household contact).
HCP 2. Outside community.
HCP 3. Outside community.

#### Samples

| **Sample type** | **Sample collection date** | **GISAID identifier** | **Clade** (Nextstrain) | **Lineage** (Pangolin) |
|---|---|---|---|---|
| HCP 1 | April 2020 | hCoV-19/USA/WI-UW-119/2020 | 20A | B.1.19 |
| HCP 2 | April 2020 | hCoV-19/USA/WI-UW-259/2020 | 20A | B.1.19 |
| HCP 3 | April 2020 | hCoV-19/USA/WI-UW-260/2020 | 20C | B.1 |
| Household contact (HCP 1) | April 2020 | hCoV-19/USA/WI-UW-120/2020 | 20A | B.1.19 |
| patient A | March 2020 | hCoV-19/USA/WI-UW-118/2020 | 20C | B.1 |

| | | | | |
|---|---|---|---|---|
| patient B | March 2020 | hCoV-19/USA/WI-UW-110/2020 | 20A | B.1.139 |

#### Epidemiological information

HCP 1 tested positive for SARS-CoV-2 after providing care for two SARS-CoV-2 positive patients, patients A and B. HCP 1 also had a household contact who tested positive for SARS-CoV-2 16 days before HCP 1. In the week following HCP 1's positive test, two additional HCP, HCP 2 and HCP 3 tested positive for SARS-CoV-2. HCPs 1, 2, and 3 all work in the same department, but we do not know whether these individuals had any high-risk contact with HCP 1 before their positive test results.

#### Alignment

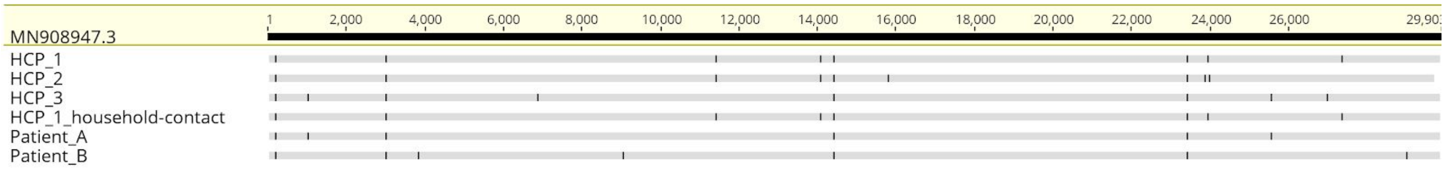


#### Phylogeny

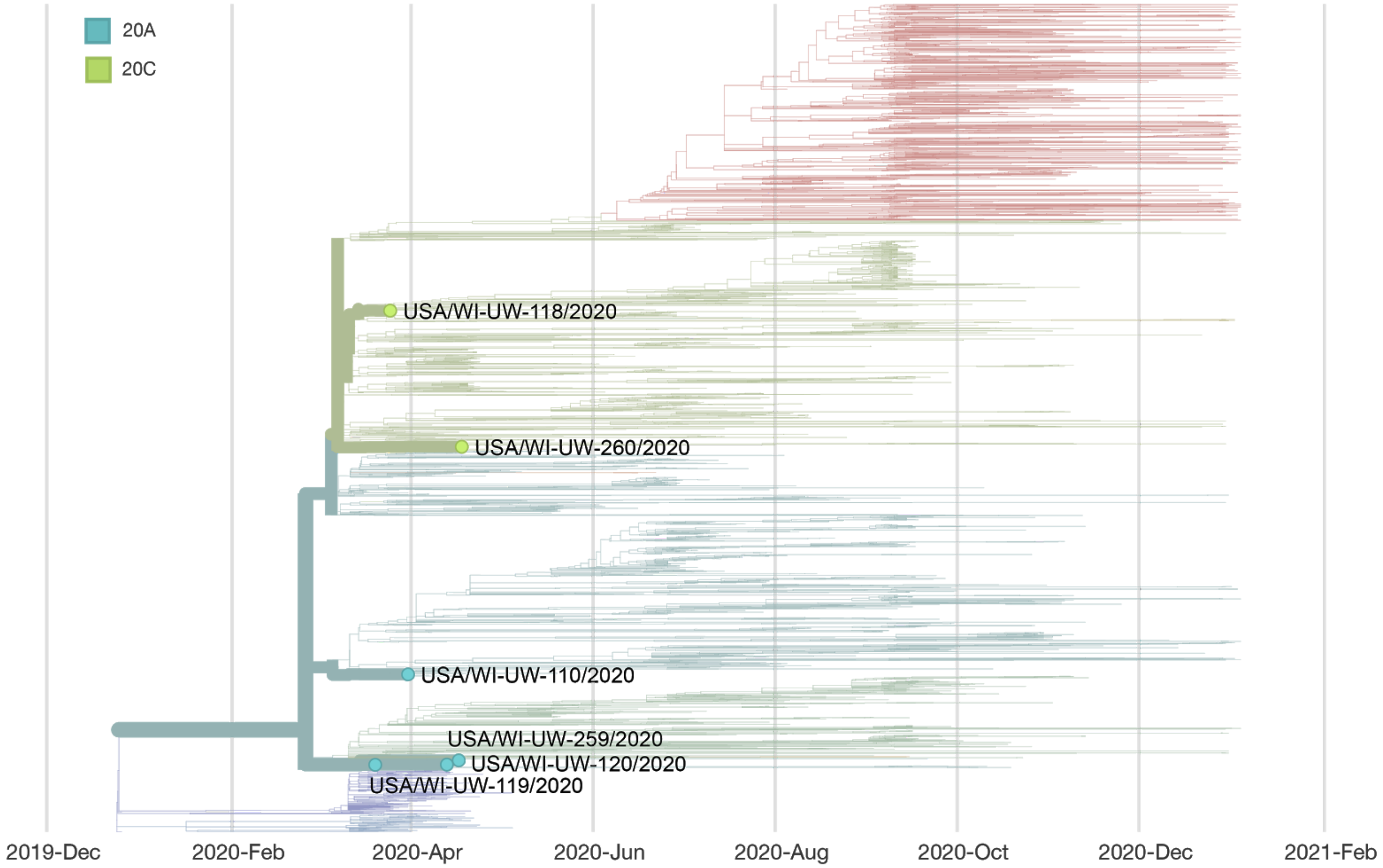


#### Notes

This case was published as an independent case report in Emerging Infectious Disease {32758345}.

### Report #3. 2020-06-04.

#### Likely source of HCP infection

HCP 1. Outside community.

#### Samples

| **Sample type** | **Sample collection date** | **GISAID identifier** | **Clade** (Nextstrain) | **Lineage** (Pangolin) |
|---|---|---|---|---|
| HCP 1 | May 2020 | hCoV-19/USA/WI-UW-388/2020 | 20A | B.1.139 |
| HCP-C | Sample was not available | | | |
| patient A | May 2020 | hCoV-19/USA/WI-UW-386/2020 | 20A | B.1.139 |
| patient B | May 2020 | hCoV-19/USA/WI-UW-390/2020 | 20A | B.1.139 |
| patient C | May 2020 | hCoV-19/USA/WI-UW-389/2020 | 20A | B.1.139 |
| patient D | May 2020 | hCoV-19/USA/WI-UW-387/2020 | 20A | B.1.139 |

#### Epidemiological information

*HCP 1 did not have direct contact with any of the patients included here*. In this case, a household contact (HCP-C) of HCP 1 also works in healthcare, but in a different healthcare facility than HCP 1. The healthcare facility employing HCP-C was experiencing a COVID-19 outbreak at the time that HCP 1 tested positive. Patients A-D were patient samples collected from the HCP-C outbreak. A sample from the HCP-C was not available for comparison. Given the similarity in viral sequences between HCP 1 and all four patients from the outside healthcare facility, it is likely HCP 1 was exposed/infected via their household contact, who was likely exposed through patient contact.

#### Alignment

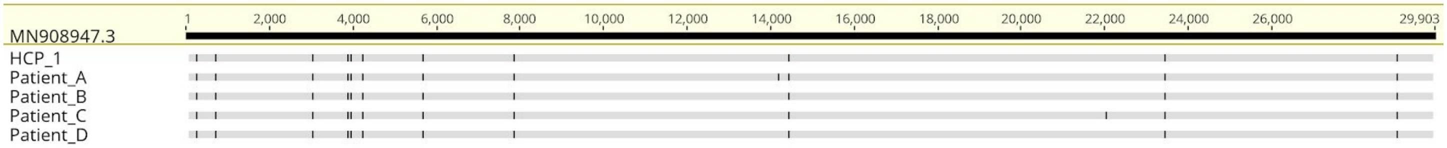

#### Phylogeny

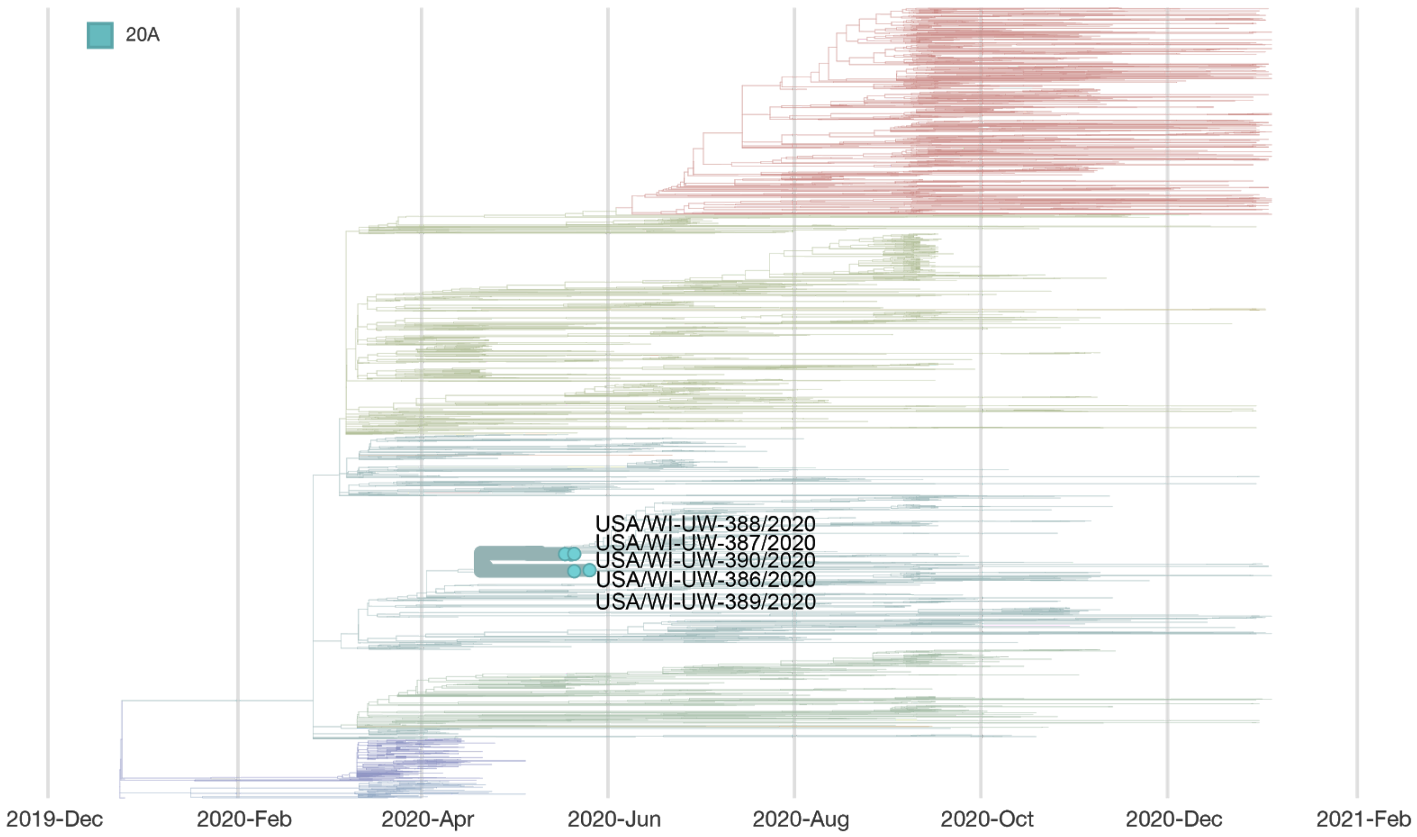


### Report #4. 2020-06-04.

#### Likely source of HCP infection

HCP 1. Patient source (patient D).

#### Samples

| **Sample type** | **Sample collection date** | **GISAID identifier** | **Clade** (Nextstrain) | **Lineage** (Pangolin) |
|---|---|---|---|---|
| HCP 1 | May 2020 | hCoV-19/USA/WI-UW-391/2020 | 20A | B.1.139 |
| patient A | May 2020 | hCoV-19/USA/WI-UW-392/2020 | 20A | B.1.276 |
| patient B | May 2020 | hCoV-19/USA/WI-UW-393/2020 | 20A | B.1.139 |
| patient C | May 2020 | N/A - no consensus sequence | | |
| patient D | May 2020 | hCoV-19/USA/WI-UW-389/2020 | 20A | B.1.139 |

#### Epidemiological information

In the two weeks before symptom onset, HCP 1 provided direct care to patients A-D. HCP 1 wore appropriate PPE while providing care and was also present during patient D’s treatment with the Aerobika nebulizer.

#### Alignment

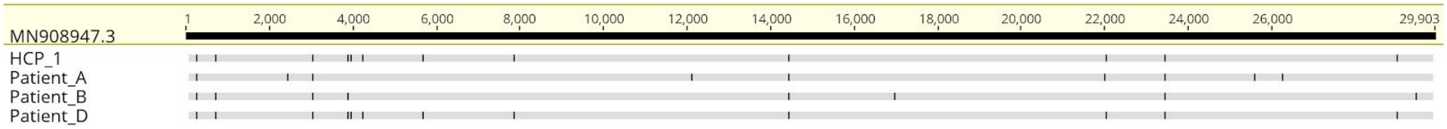


#### Phylogeny

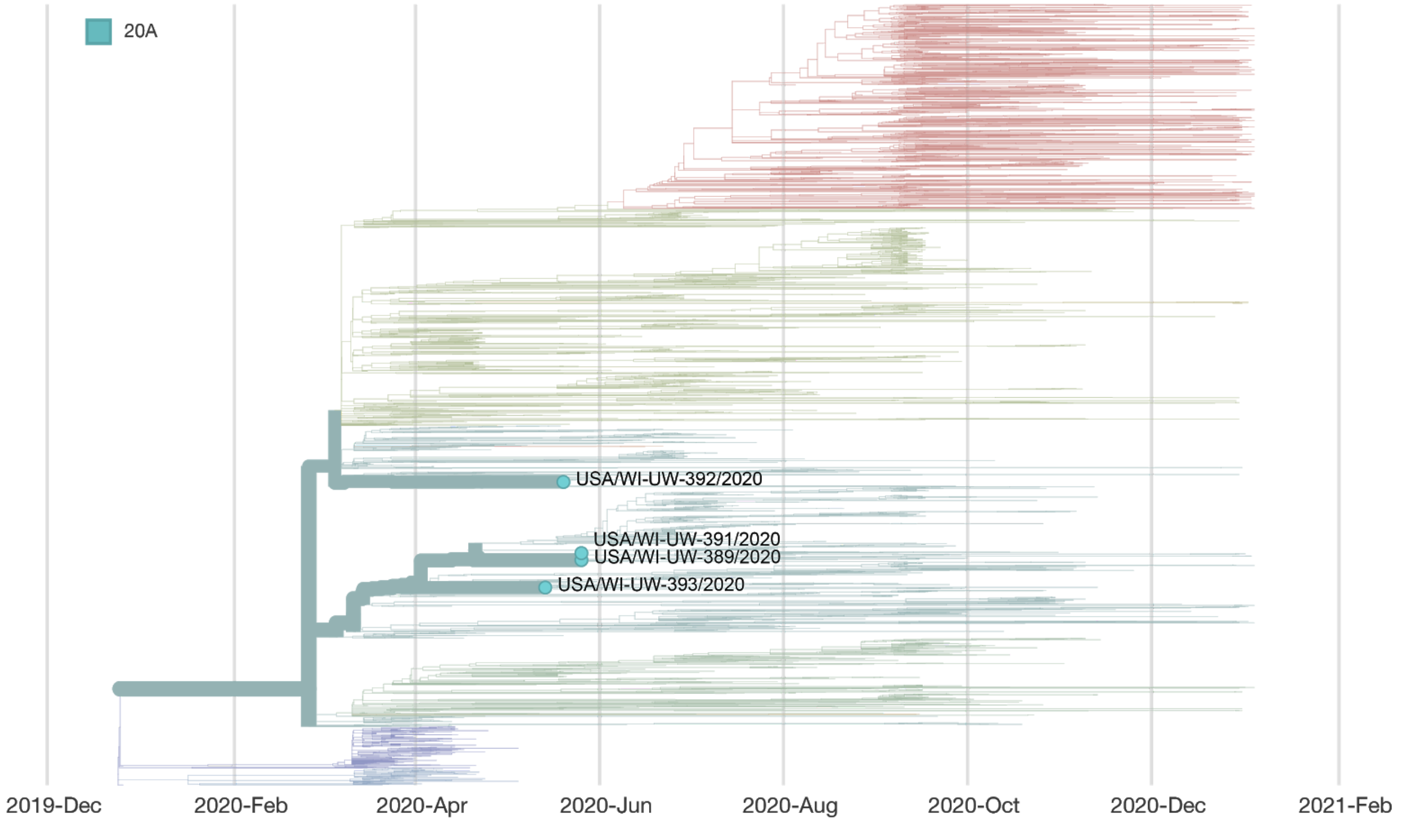


#### Notes

The Aerobika nebulizer was added to the list of aerosol-generating procedures requiring enhanced PPE based on this case.

### Report #5. 2020-07-08.

#### Likely source of HCP infection

HCP 1. Outside community.
HCP 2. Outside community (likely source was HCP 1, but infection took place outside of the workplace).

#### Samples

| **Sample type** | **Sample collection date** | **GISAID identifier** | **Clade** (Nextstrain) | **Lineage** (Pangolin) |
|---|---|---|---|---|
| HCP 1 | June 2020 | hCoV-19/USA/WI-UW-588/2020 | 20C | B.1 |

| | | | | |
|---|---|---|---|---|
| HCP 2 | June 2020 | hCoV-19/USA/WI-UW-610/2020 | 20C | B.1 |
| patient A | June 2020 | hCoV-19/USA/FL-UW-473/2020 | 20A | B.1.162 |
| patient B | June 2020 | hCoV-19/USA/WI-UW-469/2020 | 20A | B.1.162 |
| patient C | June 2020 | hCoV-19/USA/WI-UW-497/2020 | 20A | B.1.139 |
| patient D | June 2020 | hCoV-19/USA/WI-UW-493/2020 | 20A | B.1.139 |
| patient E | June 2020 | hCoV-19/USA/WI-UW-479/2020 | 20A | B.1.139 |
| patient F | June 2020 | N/A - no consensus sequence | | |

#### Epidemiological information

HCP 1 had direct contact while wearing appropriate PPE with patients A-E. HCP 1 and HCP 2 had unmasked interactions (>15 mins) with each other outside of the workplace. HCP 2 did not have contact with any SARS-CoV-2 positive patients in the 14 days before symptom onset.

#### Alignment

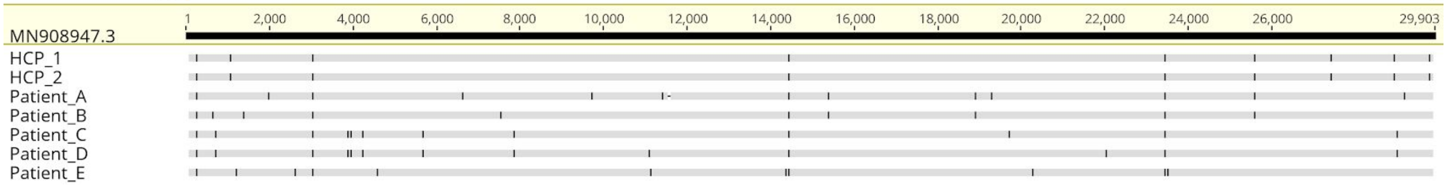


#### Phylogeny

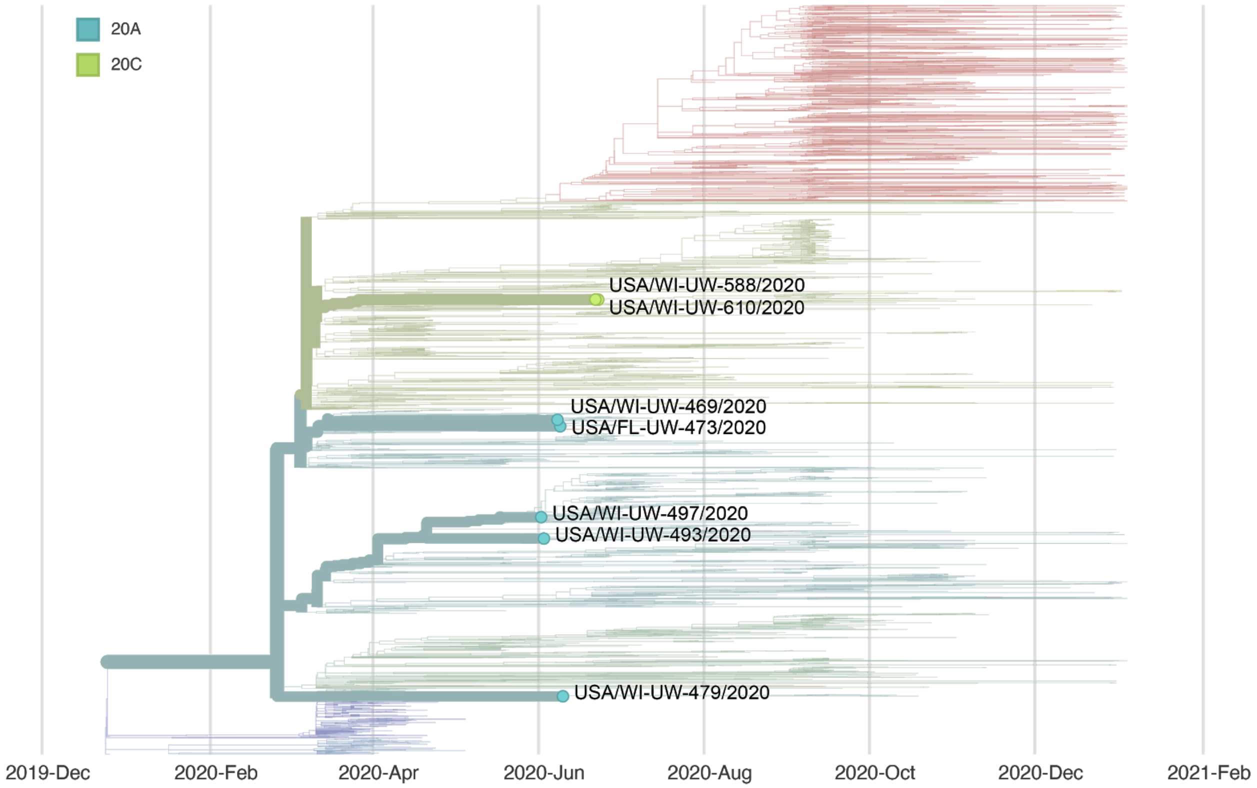

### Report #6. 2020-08-13.

#### Likely source of HCP infection

HCP 1. Outside community (based on epidemiological risk factors).
HCP 2. Employee source (likely source was HCP 1).

#### Samples

| Sample type | Sample collection date | GISAID identifier | Clade (Nextstrain) | Lineage (Pangolin) |
|---|---|---|---|---|
| HCP 1 | June 2020 | hCoV-19/USA/WI-UW-520/2020 | 20A | B.1.139 |
| HCP 2 | June 2020 | hCoV-19/USA/WI-UW-726/2020 | 20A | B.1.139 |

#### Epidemiological information

HCP 1 had a high-risk exposure event in the community before testing positive. This event was indoors, unmasked, and lasted longer than 15 minutes. HCP 1 works in the same department as HCP 2. Neither HCP 1 nor HCP 2 provided direct care to patients diagnosed with COVID-19 in the 14 days before their symptom onset. HCP 2 reported wearing a mask around all coworkers except while eating in the breakroom. HCP 2 reports removing their mask while eating, but maintaining a 6-foot physical distance from others during this time.

#### Alignment

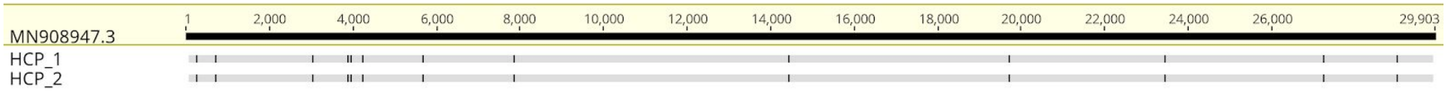


#### Phylogeny

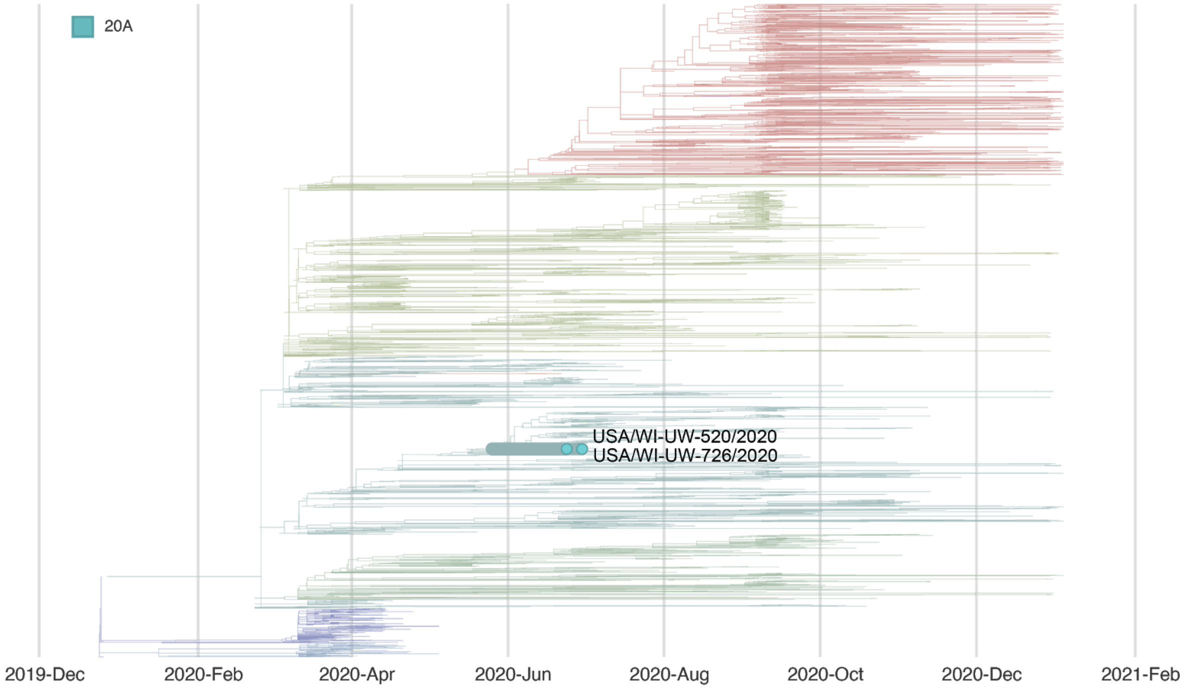

### Report #7. 2020-08-04.

#### Likely source of HCP infection

HCP 1. Outside community.

#### Samples

| **Sample type** | **Sample collection date** | **GISAID identifier** | **Clade** (Nextstrain) | **Lineage** (Pangolin) |
|---|---|---|---|---|
| HCP 1 | July 2020 | hCoV-19/USA/WI-UW-973/2020 | 20C | B.1.2 |
| patient A | July 2020 | N/A - no consensus sequence | | |
| patient B | July 2020 | N/A - no consensus sequence | | |
| patient C | July 2020 | N/A - no consensus sequence | | |
| patient D | July 2020 | N/A - no consensus sequence | | |
| patient E | July 2020 | N/A - no consensus sequence | | |
| patient F | July 2020 | hCoV-19/USA/WI-UW-852/2020 | 20A | B.1.139 |
| patient G | July 2020 | N/A - no consensus sequence | | |
| patient H | July 2020 | hCoV-19/USA/WI-UW-870/2020 | 20A | B.1.139 |
| patient I | July 2020 | hCoV-19/USA/WI-UW-847/2020 | 20A | B.1.139 |
| patient J | July 2020 | N/A - no consensus sequence | | |
| patient K | July 2020 | N/A - no consensus sequence | | |
| patient L | July 2020 | hCoV-19/USA/WI-UW-927/2020 | 20A | B.1.139 |
| patient M | July 2020 | hCoV-19/USA/WI-UW-878/2020 | 20C | B.1.369 |
| patient N | July 2020 | N/A - no consensus sequence | | |
| patient O | July 2020 | hCoV-19/USA/WI-UW-886/2020 | 20A | B.1.162 |
| patient P | July 2020 | hCoV-19/USA/WI-UW-880/2020 | 20A | B.1.139 |
| patient Q | July 2020 | hCoV-19/USA/WI-UW-894/2020 | 20B | B1.1 |
| patient R | July 2020 | N/A - no consensus sequence | | |
| patient S | July 2020 | hCoV-19/USA/WI-UW-895/2020 | 20C | B.1.1369 |
| patient T | July 2020 | hCoV-19/USA/WI-UW-876/2020 | 20C | B.1.369 |
| patient U | July 2020 | hCoV-19/USA/WI-UW-931/2020 | 20C | B.1.369 |
| patient V | July 2020 | N/A - no consensus sequence | | |

| | | | | |
|---|---|---|---|---|
| patient W | July 2020 | N/A - no consensus sequence | | |
| patient X | July 2020 | hCoV-19/USA/WI-UW-898/2020 | 20A | B.1.369 |
| patient Y | July 2020 | hCoV-19/USA/WI-UW-883/2020 | 20C | B.1 |
| patient Z | July 2020 | N/A - no consensus sequence | | |
| patient AA | July 2020 | N/A - no consensus sequence | | |

#### Epidemiological information

HCP 1 collected nasopharyngeal specimens from patients with suspected COVID-19. HCP 1 wore appropriate PPE while collecting these specimens and reported no breach in PPE. Patients A-AA were collected in the 14 days before symptom onset in HCP 1.

#### Alignment

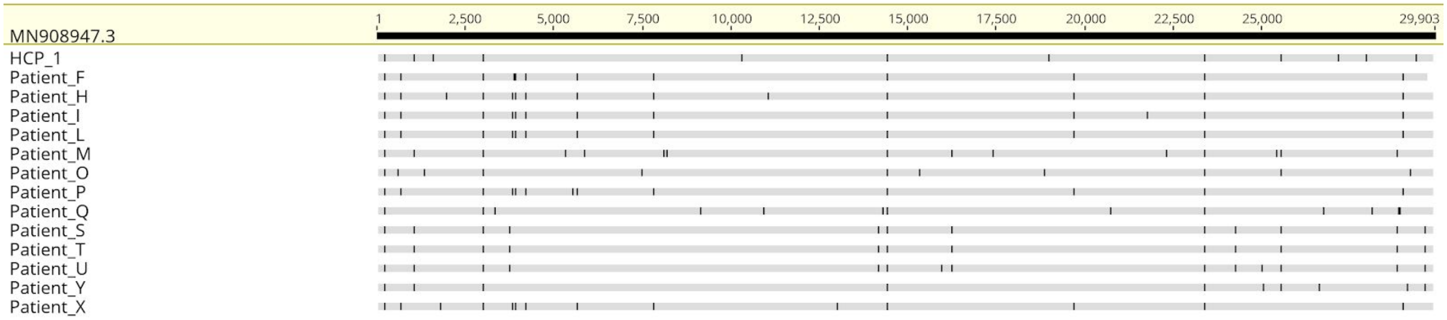


#### Phylogeny

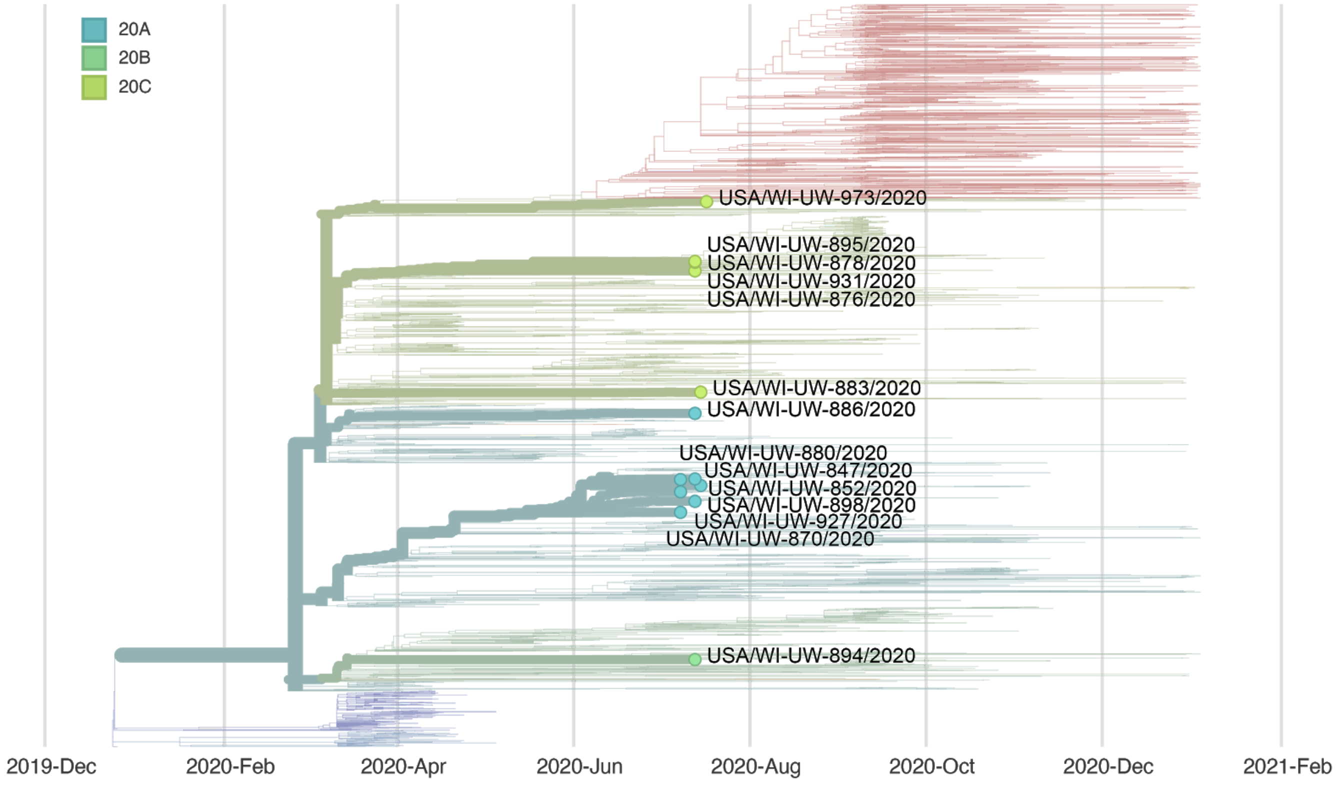

### Report #8. 2020-08-18.

#### Likely source of HCP infection

HCP 1. Outside community.

#### Samples

| Sample type | Sample collection date | GISAID identifier | Clade (Nextstrain) | Lineage (Pangolin) |
|---|---|---|---|---|
| HCP 1 | July 2020 | hCoV-19/USA/WI-UW-1022/2020 | 20B | B.1.1.73 |
| patient A | July 2020 | N/A - no consensus sequence | | |
| patient B | Unknown | N/A - no consensus sequence | | |
| patient C | July 2020 | N/A - no consensus sequence | | |
| patient D | July 2020 | hCoV-19/USA/WI-UW-923/2020 | 20A | B.1.240 |
| patient E | June 2020 | hCoV-19/USA/WI-UW-614/2020 | 20C | B.1.330 |
| patient F | July 2020 | hCoV-19/USA/WI-UW-875/2020 | 20C | B.1.369 |
| patient G | July 2020 | hCoV-19/USA/WI-UW-889/2020 | 20A | B.1.139 |
| patient H | July 2020 | hCoV-19/USA/WI-UW-1100/2020 | 20A | B.1.139 |

#### Epidemiological information

HCP 1 likely did not have direct interactions with any of the patients listed here. HCP 1 did, however, perform cleaning duties in the rooms of each of these patients in the 14 days before symptom onset.

#### Alignment

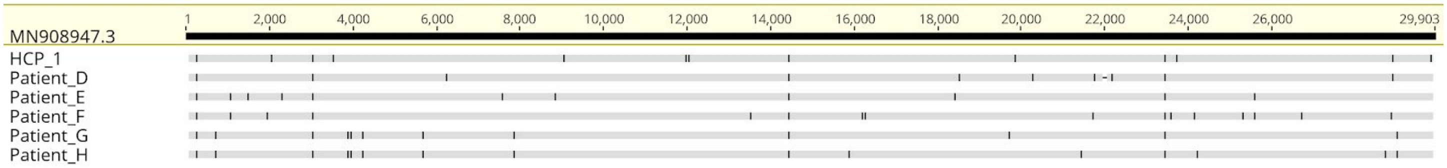

#### Phylogeny

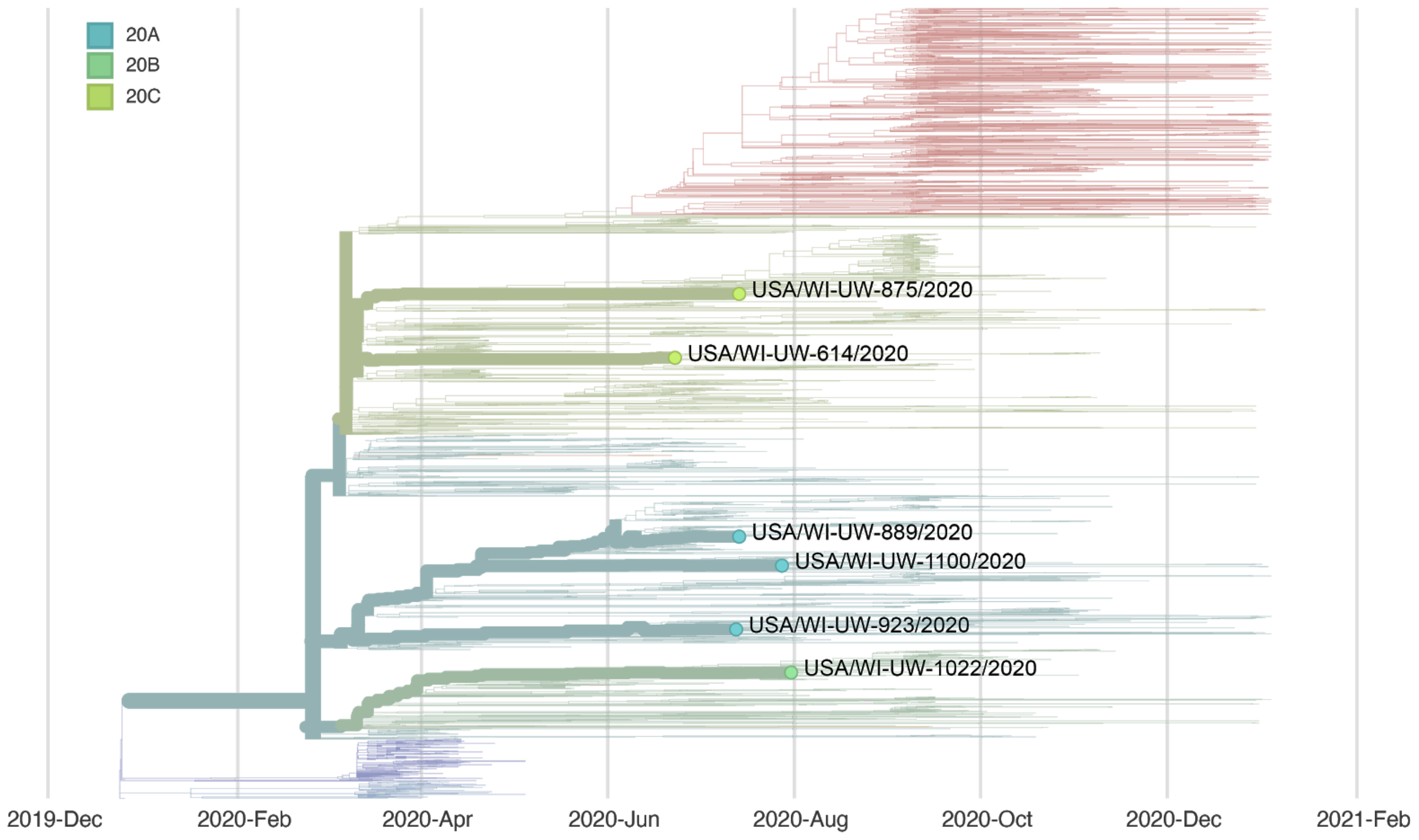


### Report #9. 2020-08-21.

#### Likely source of HCP infection

HCP 1. Outside community.

#### Samples

| **Sample type** | **Sample collection date** | **GISAID identifier** | **Clade** (Nextstrain) | **Lineage** (Pangolin) |
|---|---|---|---|---|
| HCP 1 | July 2020 | hCoV-19/USA/WI-UW-965/2020 | 20B | B.1.1.73 |
| patient A | July 2020 | hCoV-19/USA/WI-UW-765/2020 | 20A | B.1.139 |
| patient B | July 2020 | hCoV-19/USA/WI-UW-792/2020 | 20A | B.1.139 |
| patient C | July 2020 | hCoV-19/USA/WI-UW-761/2020 | 20C | B.1.3 |
| patient D | July 2020 | hCoV-19/USA/WI-UW-781/2020 | 20A | B.1.139 |
| patient E | July 2020 | hCoV-19/USA/WI-UW-794/2020 | 20A | B.1.139 |
| patient F | July 2020 | hCoV-19/USA/WI-UW-796/2020 | 20C | B.1.294 |
| patient G | July 2020 | hCoV-19/USA/WI-UW-790/2020 | 20A | B.1.139 |

| | | | | |
|---|---|---|---|---|
| patient H | July 2020 | hCoV-19/USA/WI-UW-768/2020 | 20A | B.1.139 |
| patient I | July 2020 | hCoV-19/USA/WI-UW-803/2020 | 20A | B.1.162 |
| patient J | July 2020 | N/A - no consensus sequence | | |
| patient K | July 2020 | hCoV-19/USA/MO-UW-771/2020 | 20C | B.1.370 |
| patient L | July 2020 | N/A - no consensus sequence | | |
| patient M | July 2020 | N/A - no consensus sequence | | |
| patient N | July 2020 | hCoV-19/USA/WI-UW-784/2020 | 20A | B.1 |
| patient O | July 2020 | hCoV-19/USA/WI-UW-787/2020 | 20A | B.1.139 |
| patient P | July 2020 | N/A - no consensus sequence | | |
| patient Q | July 2020 | hCoV-19/USA/WI-UW-786/2020 | 20A | B.1.112 |
| patient R | July 2020 | hCoV-19/USA/WI-UW-766/2020 | 20C | B.1.370 |
| patient S | July 2020 | hCoV-19/USA/WI-UW-798/2020 | 20A | B.1 |

#### Epidemiological information

HCP 1 collected nasopharyngeal specimens from patients with suspected COVID-19. HCP 1 wore appropriate PPE while collecting these specimens and reported no breach in PPE. Patients A-S were collected in the 14 days before symptom onset in HCP 1.

#### Alignment

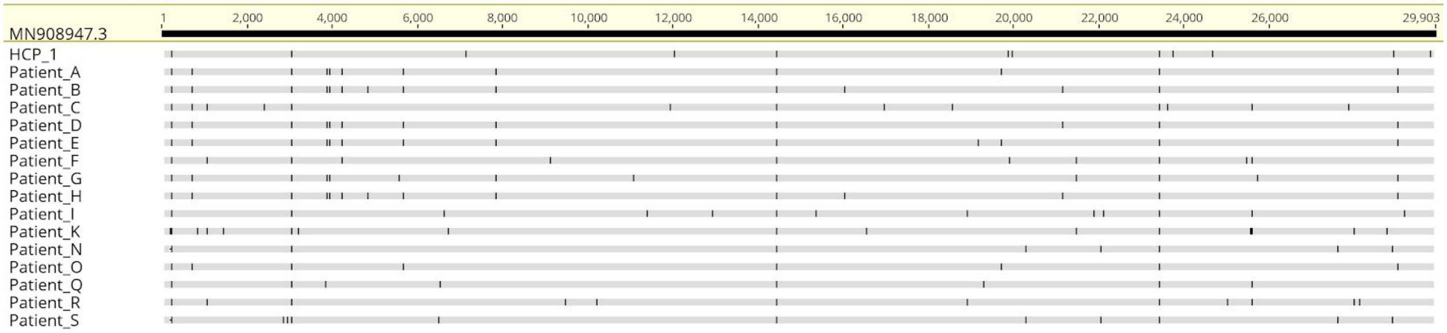

#### Phylogeny

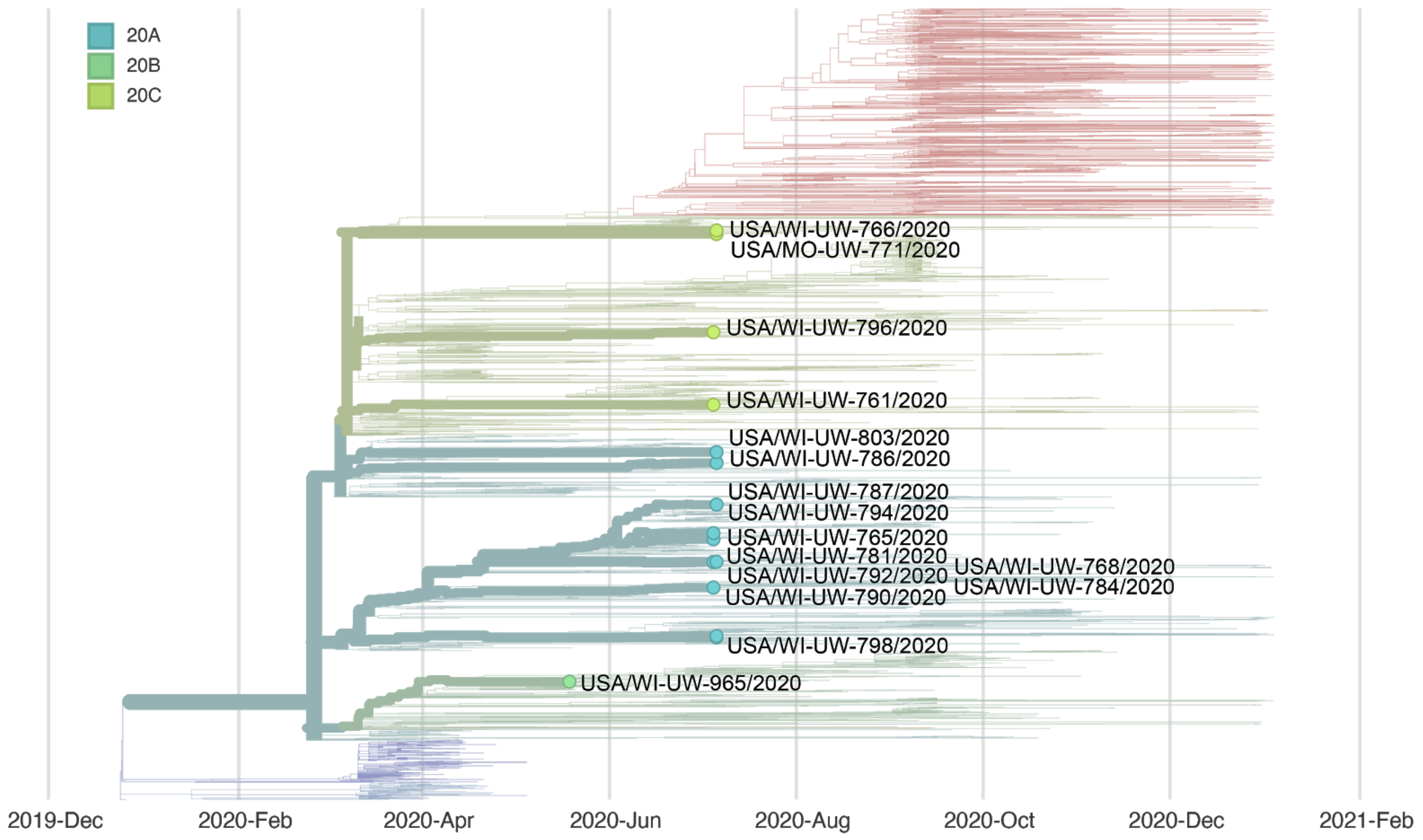


### Report #10. 2020-08-26.

#### Likely source of HCP infection

HCP 1. Patient source (patient G).

#### Samples

| **Sample type** | **Sample collection date** | **GISAID identifier** | **Clade** (Nextstrain) | **Lineage** (Pangolin) |
|---|---|---|---|---|
| HCP 1 | August 2020 | hCoV-19/USA/WI-UW-1125/2020 | 20A | B.1.139 |
| patient A | August 2020 | hCoV-19/USA/WI-UW-867/2020 | 20A | B.1.255 |
| patient B | July 2020 | hCoV-19/USA/WI-UW-889/2020 | 20A | B.1.139 |
| patient C | July 2020 | N/A - no consensus sequence | | |
| patient D | July 2020 | N/A - no consensus sequence | | |
| patient E | August 2020 | N/A - no consensus sequence | | |
| patient F | July 2020 | hCoV-19/USA/WI-UW-923/2020 | 20A | B.1.240 |
| patient G | July 2020 | hCoV-19/USA/WI-UW-1100/2020 | 20A | B.1.139 |
| patient H | July 2020 | hCoV-19/USA/WI-UW-998/2020 | 20B | B.1.1.73 |

| | | | | |
|---|---|---|---|---|
| patient I | August 2020 | hCoV-19/USA/WI-UW-1035/2020 | 20A | B.1.162 |
| patient J | July 2020 | N/A - no consensus sequence | | |
| patient K | July 2020 | N/A - no consensus sequence | | |
| patient L | July 2020 | N/A - no consensus sequence | | |
| patient M | July 2020 | hCoV-19/USA/WI-UW-1054/2020 | 20B | B.1.1.73 |
| patient N | July 2020 | hCoV-19/USA/WI-UW-805/2020 | 20C | B.1.330 |
| patient O | July 3030 | hCoV-19/USA/WI-UW-1090/2020 | 20B | B.1.1.73 |

#### Epidemiological information

HCP 1 had direct contact while wearing appropriate PPE with patients A-O in the 14 days before symptom onset.

#### Alignment

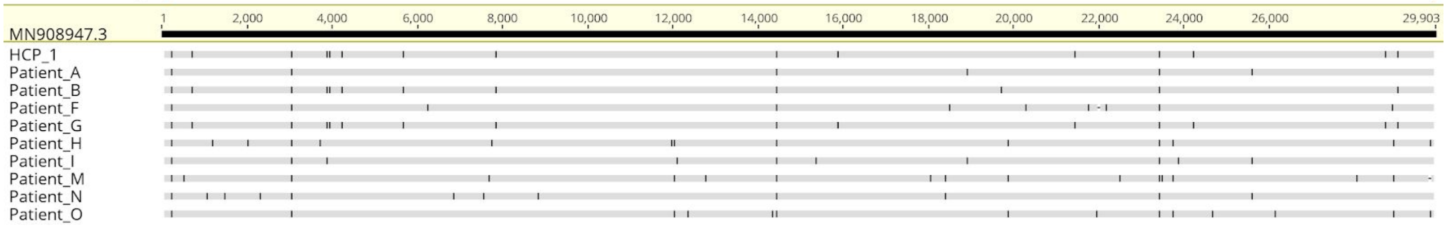


#### Phylogeny

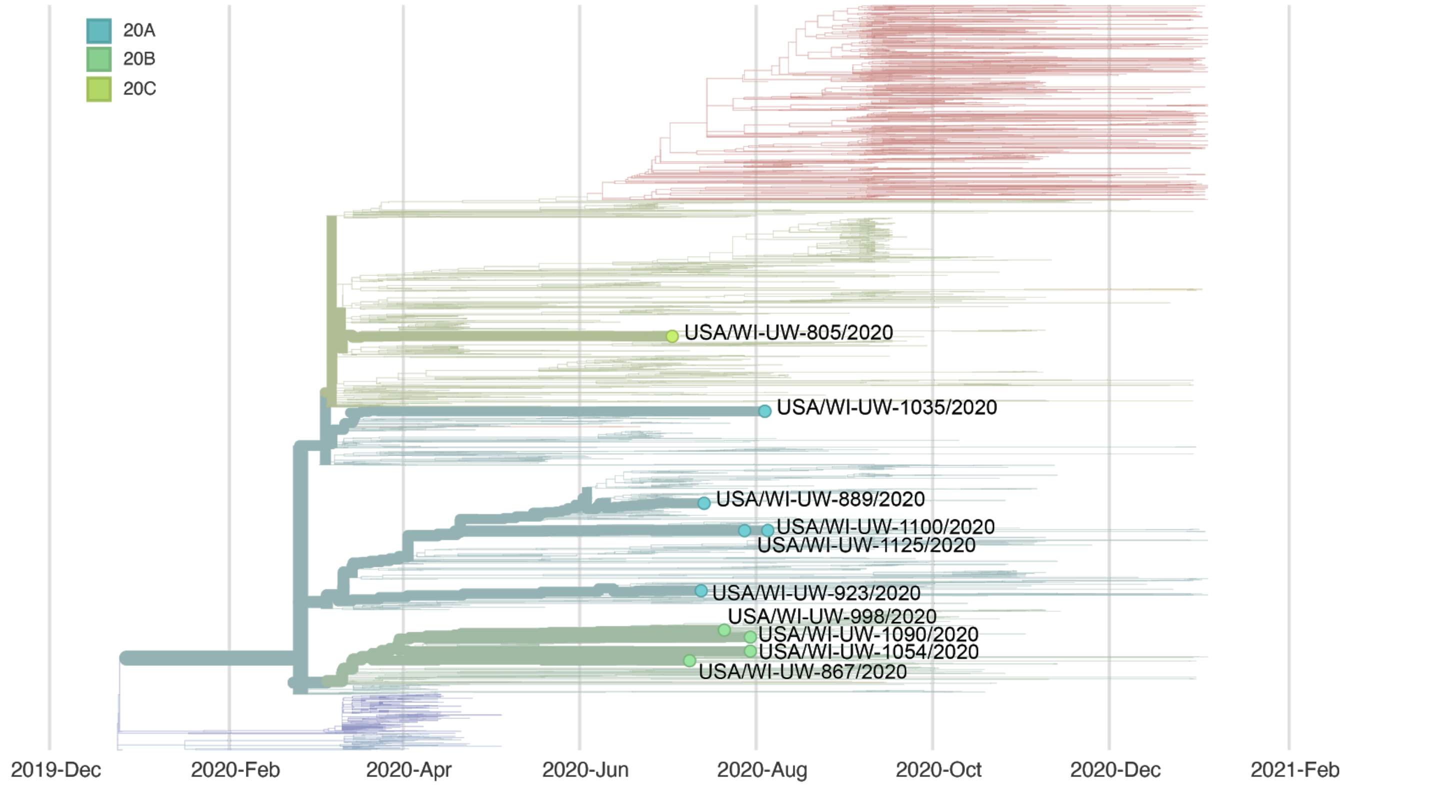

### Report #11. 2020-09-11.

#### Likely source of HCP infection

HCP 1. Outside community.

#### Samples

| **Sample type** | **Sample collection date** | **GISAID identifier** | **Clade** (Nextstrain) | **Lineage** (Pangolin) |
|---|---|---|---|---|
| HCP 1 | August 2020 | hCoV-19/USA/WI-UW-1213/2020 | 20C | B.1.2 |
| patient A | August 2020 | N/A - no consensus sequence | | |
| patient B | August 2020 | hCoV-19/USA/WI-UW-1163/2020 | 20B | B.1.1.130 |
| patient C | August 2020 | hCoV-19/USA/WI-UW-1240/2020 | 20C | B.1.2 |
| patient D | August 2020 | hCoV-19/USA/WI-UW-1170/2020 | 20A | B.1.5 |

#### Epidemiological information

HCP 1 had direct contact while wearing appropriate PPE with patients A-D in the 14 days before symptom onset. HCP 1 denied any lapses in PPE and did not interact with coworkers without a surgical mask on.

#### Alignment

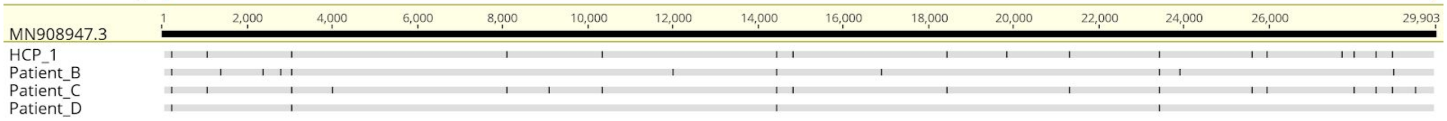

#### Phylogeny

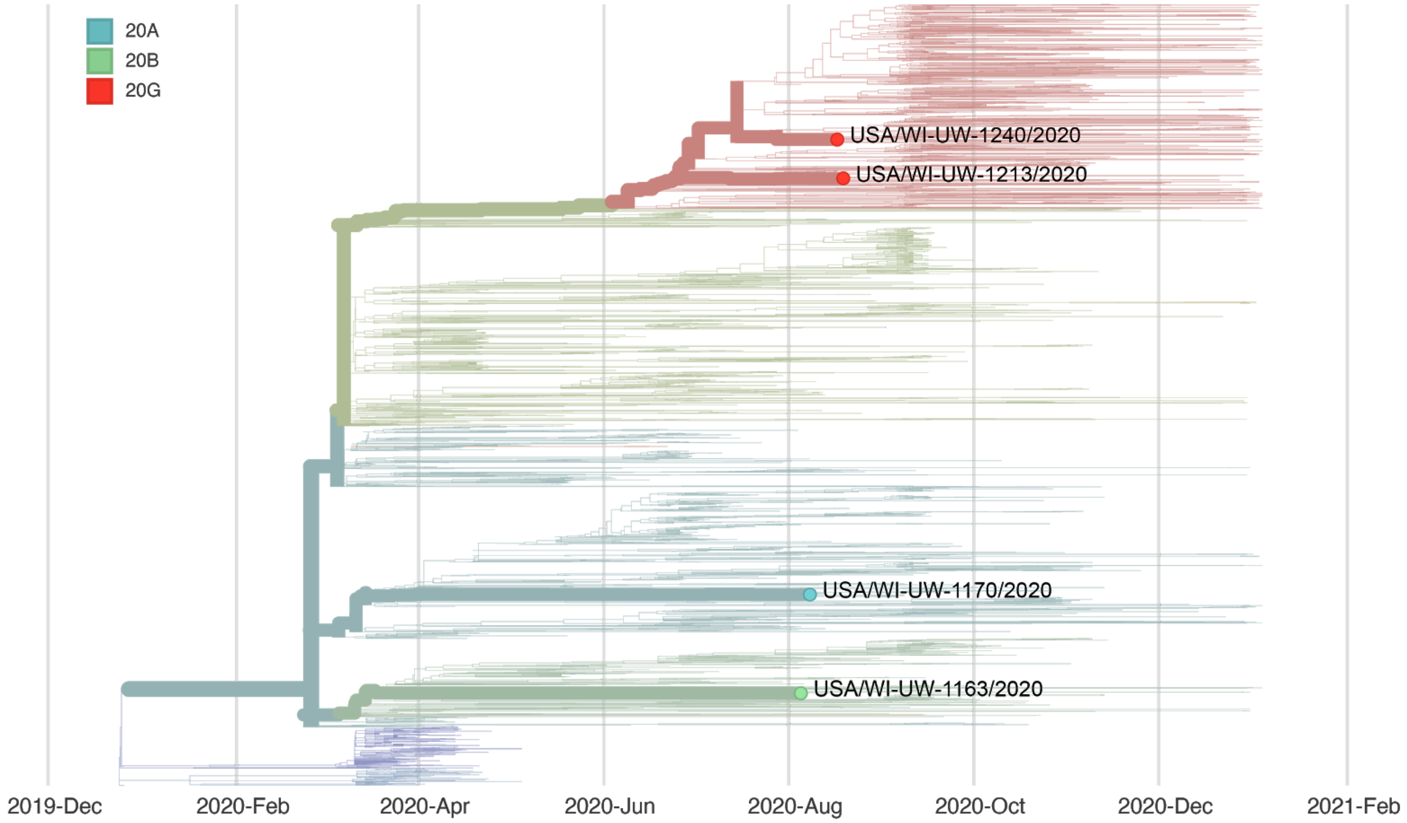


### Report #12. 2020-09-11.

#### Likely source of HCP infection

HCP 1. Inconclusive.
HCP 2. Outside community.

#### Samples

| **Sample type** | **Sample collection date** | **GISAID identifier** | **Clade** (Nextstrain) | **Lineage** (Pangolin) |
|---|---|---|---|---|
| HCP 1 | August 2020 | N/A - no consensus sequence | | |
| HCP 2 | August 2020 | hCoV-19/USA/WI-UW-1278/2020 | 20A | B.1.139 |
| patient A | July 2020 | hCoV-19/USA/WI-UW-1100/2020 | 20A | B.1.139 |
| patient B | July 2020 | N/A - no consensus sequence | | |
| patient C | July 2020 | N/A - no consensus sequence | | |
| patient D | August 2020 | N/A - no consensus sequence | | |

| | | | | |
|---|---|---|---|---|
| patient E | July 2020 | hCoV-19/USA/WI-UW-850/2020 | 20C | B.1.2 |
| patient F | August 2020 | hCoV-19/USA/WI-UW-1248/2020 | 20B | B.1.1.244 |
| patient G | August 2020 | hCoV-19/USA/WI-UW-1204/2020 | 20A | B.1.139 |
| patient H | August 2020 | N/A - no consensus sequence | | |

#### Epidemiological information

HCP 1 and HCP 2 are household contacts and one or both of these HCP provided direct patient care to patients A-H while wearing appropriate PPE in the two weeks before their symptom onset.

#### Alignment

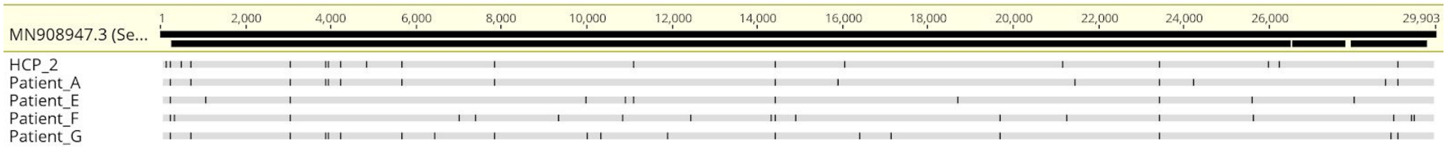


#### Phylogeny

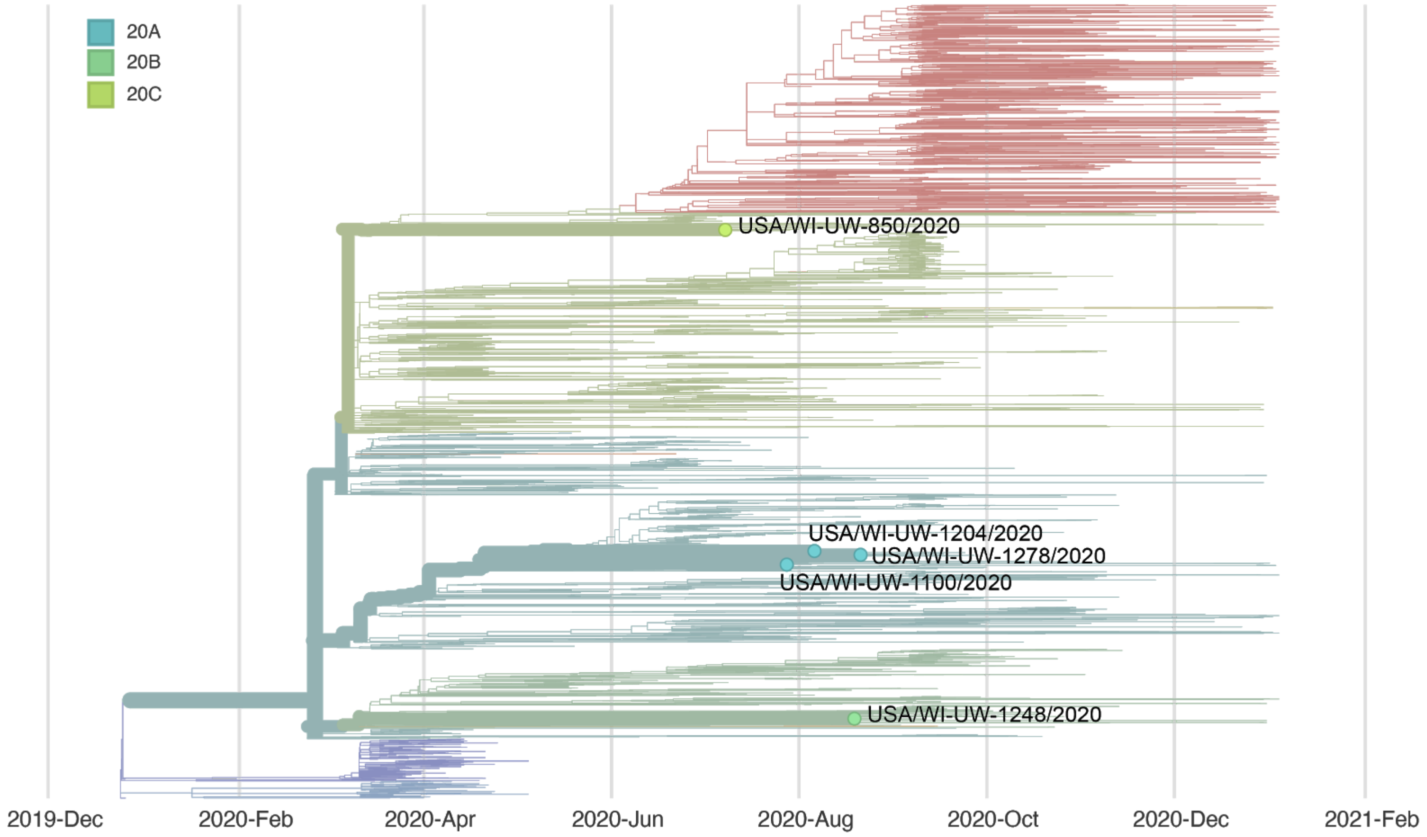

### Report #13. 2020-09-14.

#### Likely source of HCP infection

HCP 1. Outside community.

#### Samples

| **Sample type** | **Sample collection date** | **GISAID identifier** | **Clade** (Nextstrain) | **Lineage** (Pangolin) |
|---|---|---|---|---|
| HCP 1 | August 2020 | hCoV-19/USA/WI-UW-1305/2020 | 20A | B.1.139 |
| patient A | July 2020 | hCoV-19/USA/WI-UW-1100/2020 | 20A | B.1.139 |
| patient B | August 2020 | hCoV-19/USA/WI-UW-1231/2020 | 20C | B.1.337 |
| patient C | August 2020 | N/A - no consensus sequence | | |
| patient D | August 2020 | N/A - no consensus sequence | | |

#### Epidemiological information

HCP 1 provided direct care while wearing appropriate PPE to patients A-D in the 14 days before symptom onset.

#### Alignment

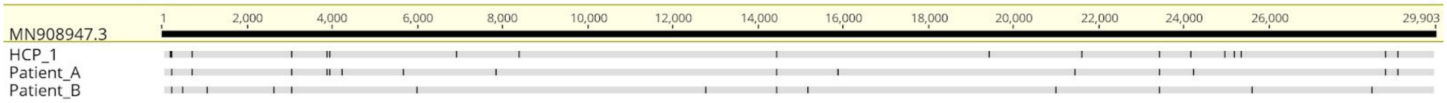


#### Phylogeny

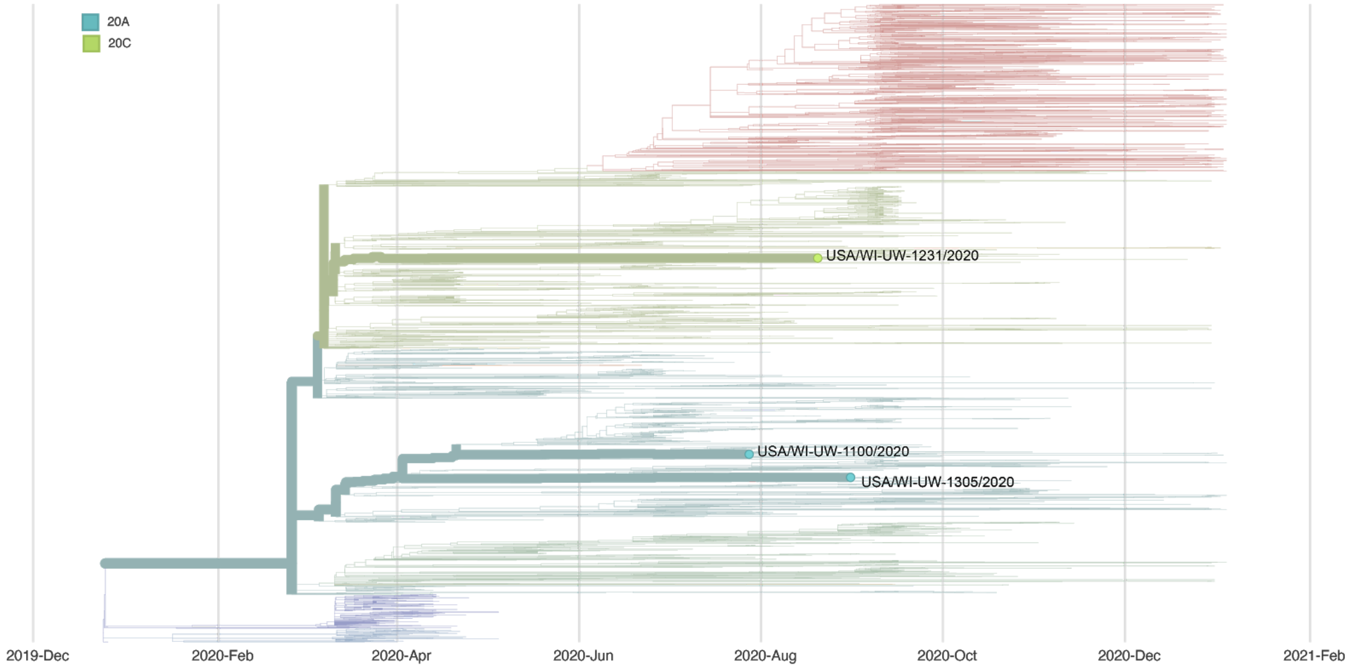

### Report #14. 2020-09-22.

#### Likely source of HCP infection

HCP 1. Outside community.
HCP 2. Combined patient and employee cluster.
HCP 3. Inconclusive.
HCP 4. Combined patient and employee cluster.
HCP 5. Outside community.
HCP 6. Employee source (HCP 9 is a likely source of infection, although the HCP 6 → HCP 9 is also possible).
HCP 7. Outside community.
HCP 8. Inconclusive.
HCP 9. Outside community.
HCP 10. Inconclusive.
HCP 11. Combined patient and employee cluster.
HCP 12. Combined patient and employee cluster.
HCP 13. Combined patient and employee cluster.
HCP 14. Inconclusive.
HCP 15. Inconclusive.
HCP 16. Combined patient and employee cluster.
HCP 17. Combined patient and employee cluster.
HCP 18. Inconclusive.
HCP 19. Inconclusive.
HCP 20. Combined patient and employee cluster.
HCP 21. Inconclusive.

Patients A and C are the patients involved in this combined patient and employee cluster.

#### Samples

| **Sample type** | **Sample collection date** | **GISAID identifier** | **Clade** (Nextstrain) | **Lineage** (Pangolin) |
|---|---|---|---|---|
| HCP 1 | September 2020 | hCoV-19/USA/WI-UW-1348/2020 | 20G | B.1.2 |
| HCP 2 | September 2020 | hCoV-19/USA/WI-UW-1344/2020 | 20G | B.1.2 |
| HCP 3 | September 2020 | N/A - no consensus sequence | | |
| HCP 4 | September 2020 | hCoV-19/USA/WI-UW-1412/2020 | 20G | B.1.2 |
| HCP 5 | September 2020 | hCoV-19/USA/WI-UW-1325/2020 | 20G | B.1.5 |
| HCP 6 | September 2020 | hCoV-19/USA/WI-UW-1480/2020 | 20B | B.1.1.251 |
| HCP 7 | September 2020 | hCoV-19/USA/WI-UW-1477/2020 | 20G | B.1.369 |
| HCP 8 | September 2020 | N/A - no consensus sequence | | |
| HCP 9 | September 2020 | hCoV-19/USA/WI-UW-1475/2020 | 20B | B.1.1.251 |
| HCP 10 | September 2020 | N/A - no consensus sequence | | |

| | | | | |
|---|---|---|---|---|
| HCP 11 | September 2020 | hCoV-19/USA/WI-UW-1441/2020 | 20G | B.1.2 |
| HCP 12 | September 2020 | hCoV-19/USA/WI-UW-1445/2020 | 20G | B.1.2 |
| HCP 13 | September 2020 | hCoV-19/USA/WI-UW-1452/2020 | 20G | B.1.2 |
| HCP 14 | September 2020 | N/A - no consensus sequence | | |
| HCP 15 | September 2020 | N/A - no consensus sequence | | |
| HCP 16 | September 2020 | hCoV-19/USA/WI-UW-1459/2020 | 20G | B.1.2 |
| HCP 17 | September 2020 | hCoV-19/USA/WI-UW-1436/2020 | 20G | B.1.2 |
| HCP 18 | September 2020 | N/A - no consensus sequence | | |
| HCP 19 | September 2020 | N/A - no consensus sequence | | |
| HCP 20 | September 2020 | hCoV-19/USA/WI-UW-1461/2020 | 20G | B.1.2 |
| HCP 21 | September 2020 | N/A - no consensus sequence | | |
| patient A | September 2020 | hCoV-19/USA/WI-UW-1406/2020 | 20G | B.1.2 |
| patient B | September 2020 | hCoV-19/USA/WI-UW-1499/2020 | 20G | B.1.2 |
| patient C | September 2020 | hCoV-19/USA/WI-UW-1900/2020 | 20G | B.1.2 |
| patient D | September 2020 | hCoV-19/USA/WI-UW-1941/2020 | 20G | B.1.2 |

#### Epidemiological information

These HCP work in the same department. HCP 2, 4, 11, 12, 13, 16, and 20 provided direct care to patient A and/or patient C. HCP 17 and HCP 1 did not provide direct care to patient A or C.

#### Alignment

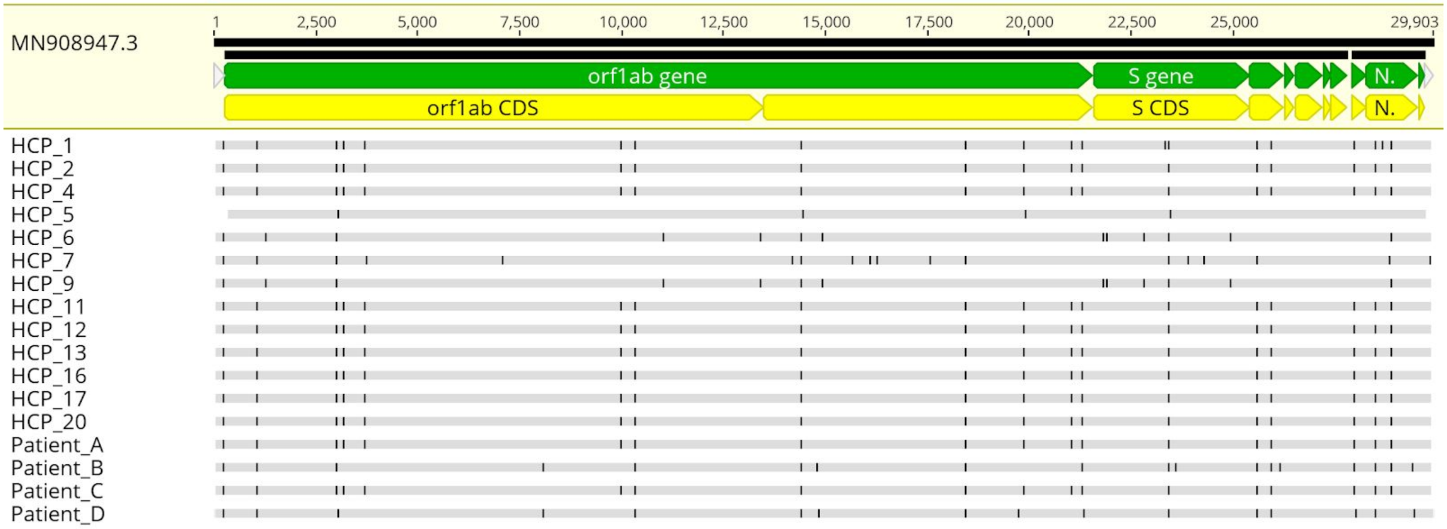

#### Phylogeny

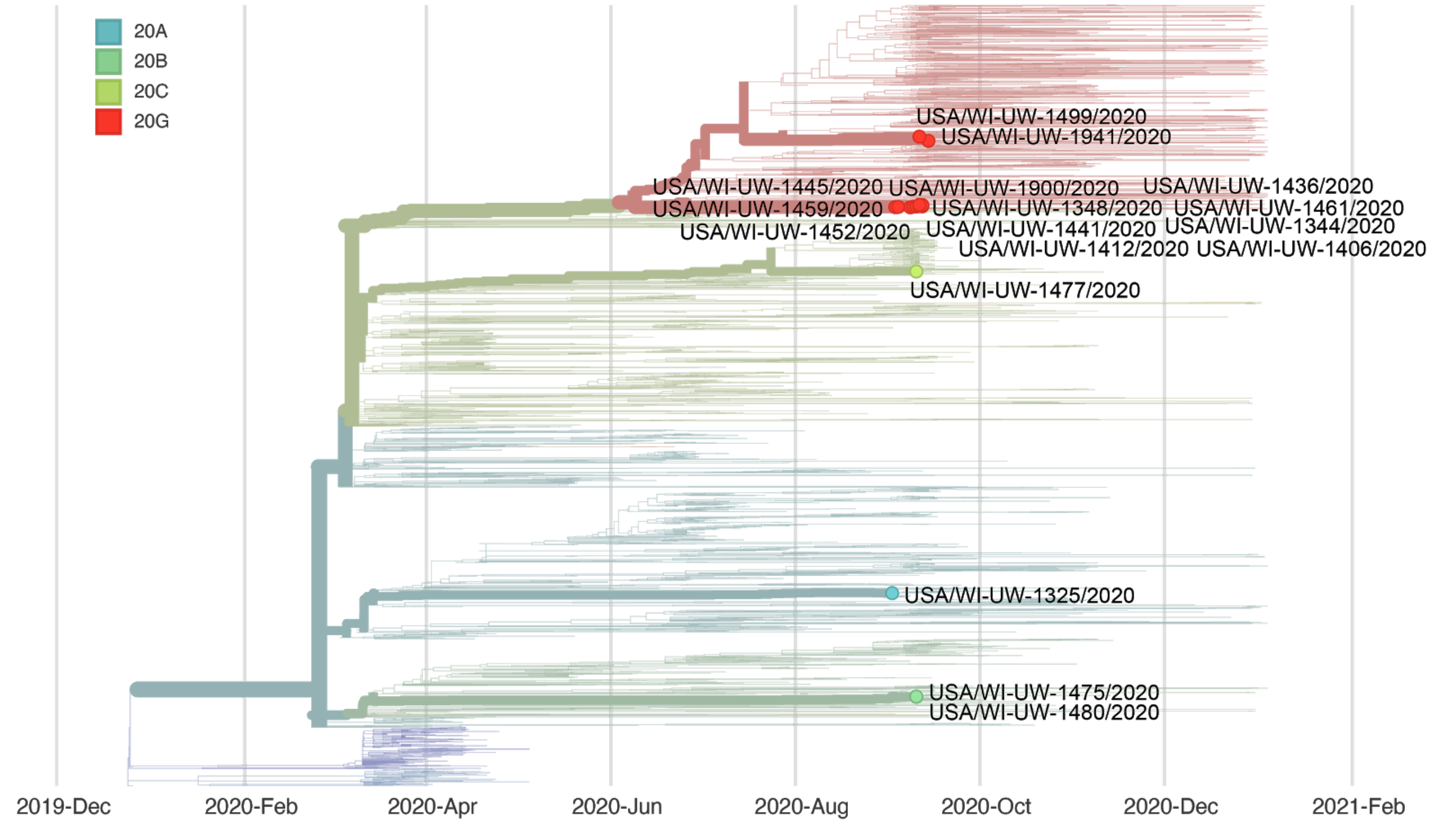


### Report #15. 2020-09-18.

#### Likely source of HCP infection

HCP 1. Outside community.

#### Samples

| **Sample type** | **Sample collection date** | **GISAID identifier** | **Clade** (Nextstrain) | **Lineage** (Pangolin) |
|---|---|---|---|---|
| HCP 1 | September 2020 | hCoV-19/USA/WI-UW-1347/2020 | 20G | B.1.2 |
| patient A | August 2020 | hCoV-19/USA/WI-UW-1220/2020 | 20G | B.1.2 |

#### Epidemiological information

HCP 1 provided direct care to patient A while wearing appropriate PPE and with no reported lapses in PPE use.

#### Alignment

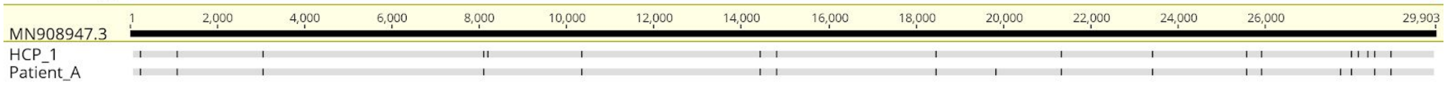

#### Phylogeny

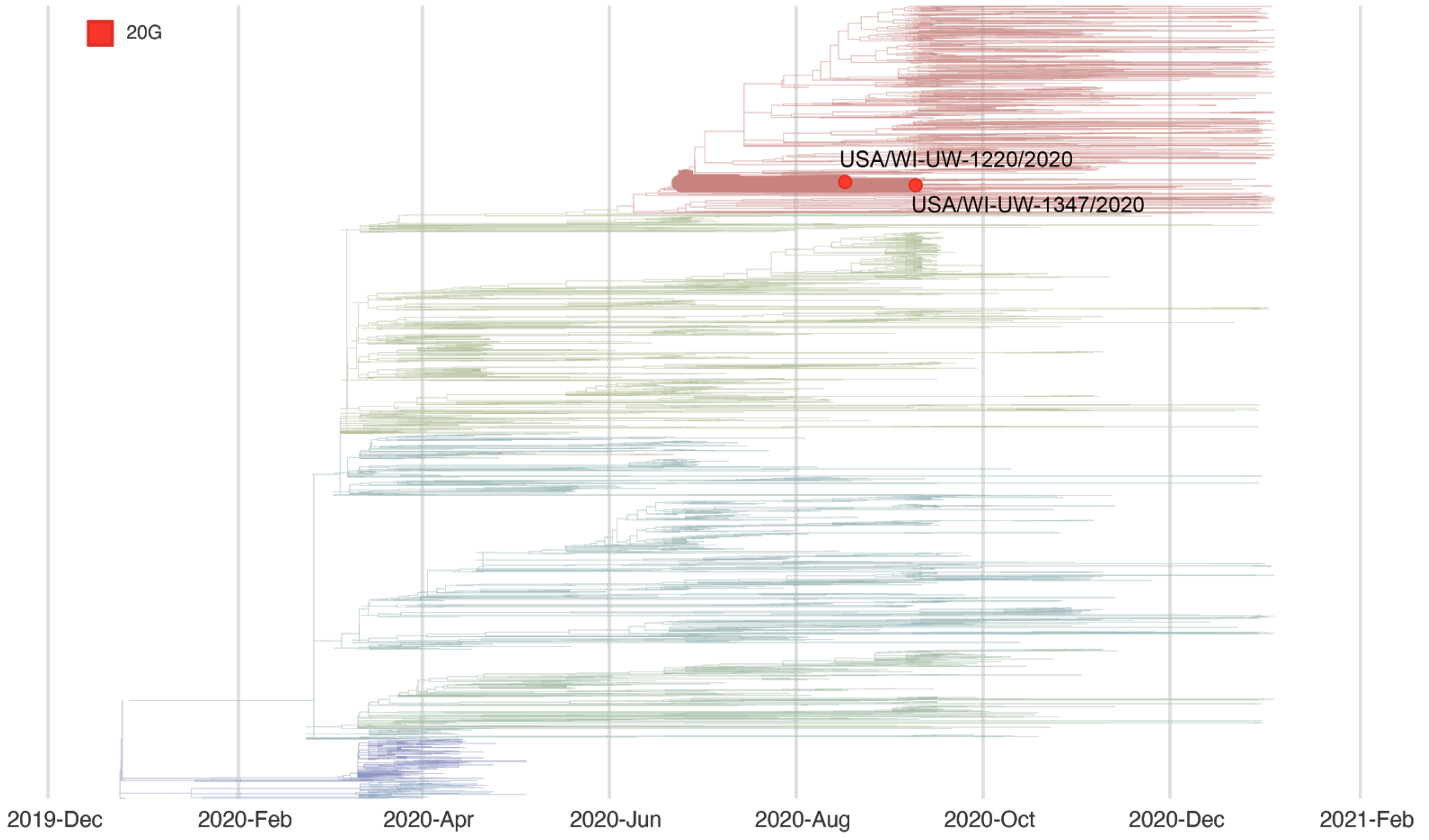


### Report #16. 2020-10-29.

#### Likely source of HCP infection

HCP 1. Employee source (HCP 2).
HCP 2. Inconclusive.
HCP 3. Outside community.

#### Samples

| **Sample type** | **Sample collection date** | **GISAID identifier** | **Clade** (Nextstrain) | **Lineage** (Pangolin) |
|---|---|---|---|---|
| HCP 1 | September 2020 | hCoV-19/USA/WI-UW-1901/2020 | 20G | B.1.2 |
| HCP 2 | September 2020 | hCoV-19/USA/WI-UW-1350/2020 | 20G | B.1.2 |
| HCP 3 | September 2020 | hCoV-19/USA/WI-UW-1898/2020 | 20A | B.1 |

#### Epidemiological information

Contact tracing revealed HCP 2 worked for two days prior to symptom onset and may have had unmasked contact with HCP 1 during overlapping meal breaks. Contact tracing additionally revealed HCP 3 had a high-risk exposure even lasting >15 minutes in the outside community prior to testing positive.

#### Alignment

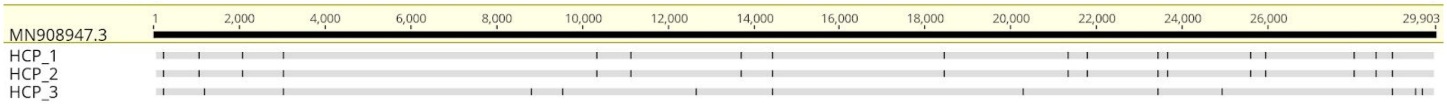


#### Phylogeny

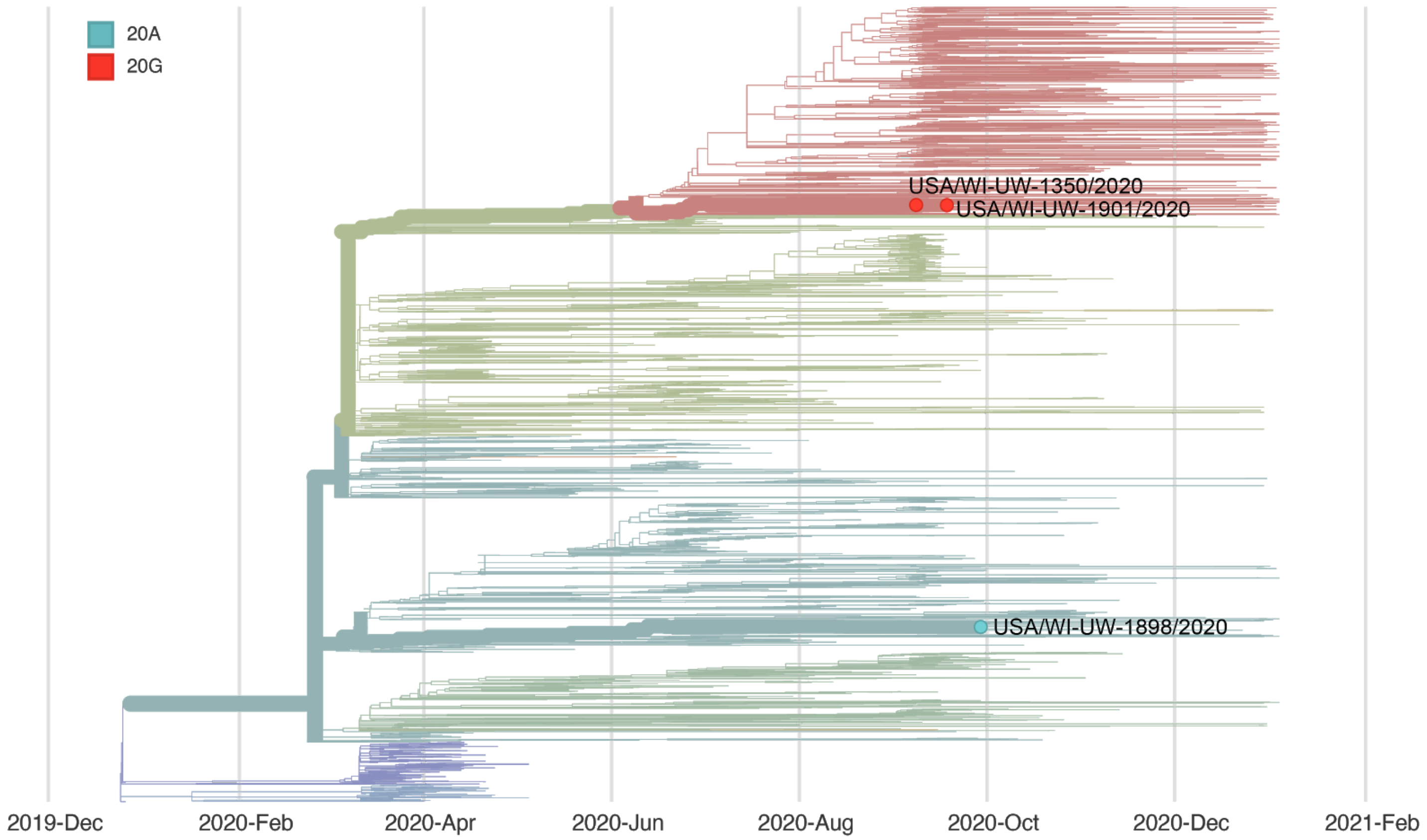


### Report #17. 2020-10-29.

#### Likely source of HCP infection

HCP 1. Outside community.

#### Samples

| Sample type | Sample collection date | GISAID identifier | Clade (Nextstrain) | Lineage (Pangolin) |
|---|---|---|---|---|
| HCP 1 | September 2020 | hCoV-19/USA/WI-UW-1895/2020 | 20C | B.1.369 |
| patient A | September 2020 | N/A - no consensus sequence | | |
| patient B | July 2020 | hCoV-19/USA/WI-UW-774/2020 | 20A | B.1.139 |
| patient C | August 2020 | hCoV-19/USA/WI-UW-1198/2020 | 20A | B.1.139 |

| | | | | |
|---|---|---|---|---|
| patient D | August 2020 | hCoV-19/USA/WI-UW-1166/2020 | 20A | B.1.139 |
| patient E | August 2020 | N/A - no consensus sequence | | |
| patient F | September 2020 | N/A - no consensus sequence | | |
| patient G | August 2020 | N/A - no consensus sequence | | |
| patient H | August 2020 | hCoV-19/USA/WI-UW-1301/2020 | 20C | B.1 |
| patient I | August 2020 | N/A - no consensus sequence | | |

#### Epidemiological information

HCP 1 provided direct care to patients A-I while wearing appropriate PPE and with no reported lapses in PPE use. HCP 1 also had a household contact with confirmed SARS-CoV-2 infection who had symptoms onset prior to HCP 1's symptom onset.

#### Alignment

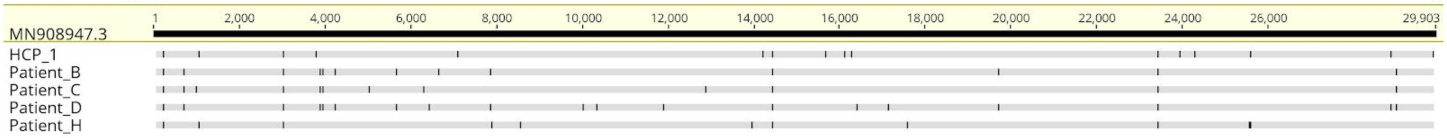


#### Phylogeny

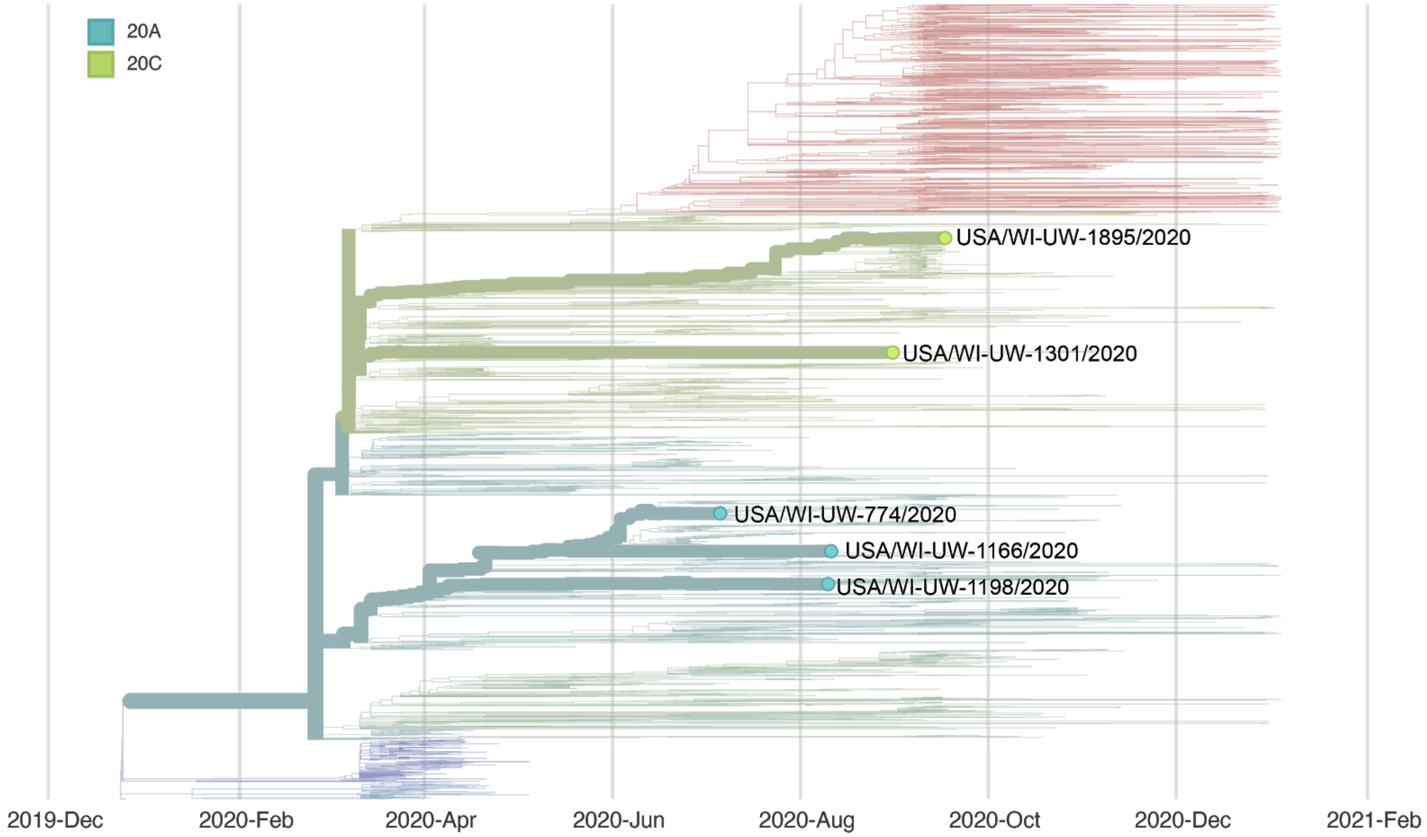

#### Report #18. 2020-10-29.

##### Likely source of HCP infection

HCP 1. Outside community.
HCP 2. Outside community.
HCP 3. Outside community.

##### Samples

| **Sample type** | **Sample collection date** | **GISAID identifier** | **Clade** (Nextstrain) | **Lineage** (Pangolin) |
|---|---|---|---|---|
| HCP 1 | September 2020 | hCoV-19/USA/WI-UW-1475/2020 | 20B | B.1.1.251 |
| HCP 2 | September 2020 | hCoV-19/USA/WI-UW-1896/2020 | 20B | B.1.1.251 |
| HCP 3 | September 2020 | hCoV-19/USA/WI-UW-1894/2020 | 20B | B.1.1.251 |

##### Epidemiological information

HCP 1 worked with HCP 3. HCP 3 was a household contact of HCP 2. None of these HCP had direct interactions with patients with known SARS-CoV-2 infection.

##### Alignment

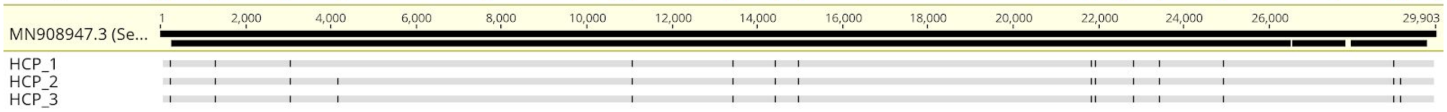


##### Phylogeny

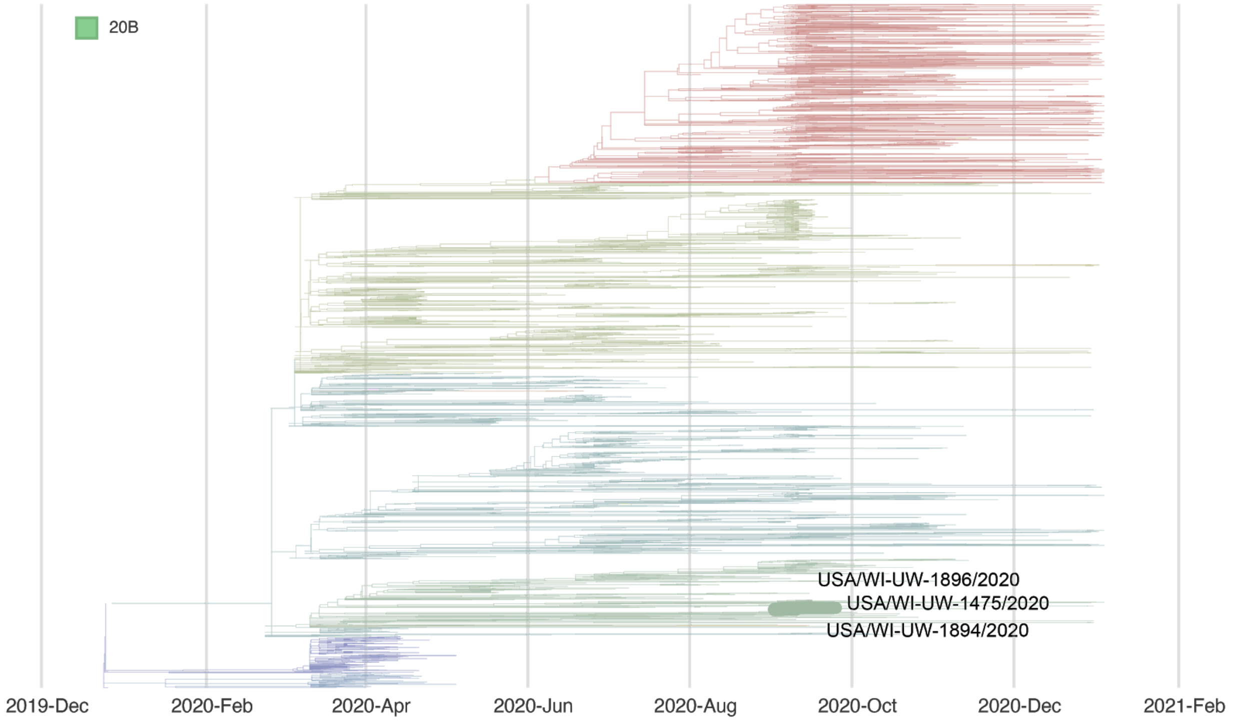

### Report #19. 2020-11-05.

#### Likely source of HCP infection

HCP 1. Outside community.

#### Samples

| **Sample type** | **Sample collection date** | **GISAID identifier** | **Clade** (Nextstrain) | **Lineage** (Pangolin) |
|---|---|---|---|---|
| HCP 1 | October 2020 | hCoV-19/USA/WI-UW-1928/2020 | 19A | B.1 |
| patient A | September 2020 | hCoV-19/USA/WI-UW-1930/2020 | 20G | B.1.2 |

#### Epidemiological information

In the two weeks before symptom onset, HCP 1 provided direct care to patient A. HCP 1 wore appropriate PPE while providing care and reported no lapses in PPE use.

#### Alignment

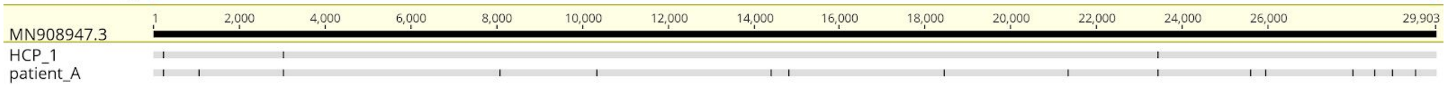


#### Phylogeny

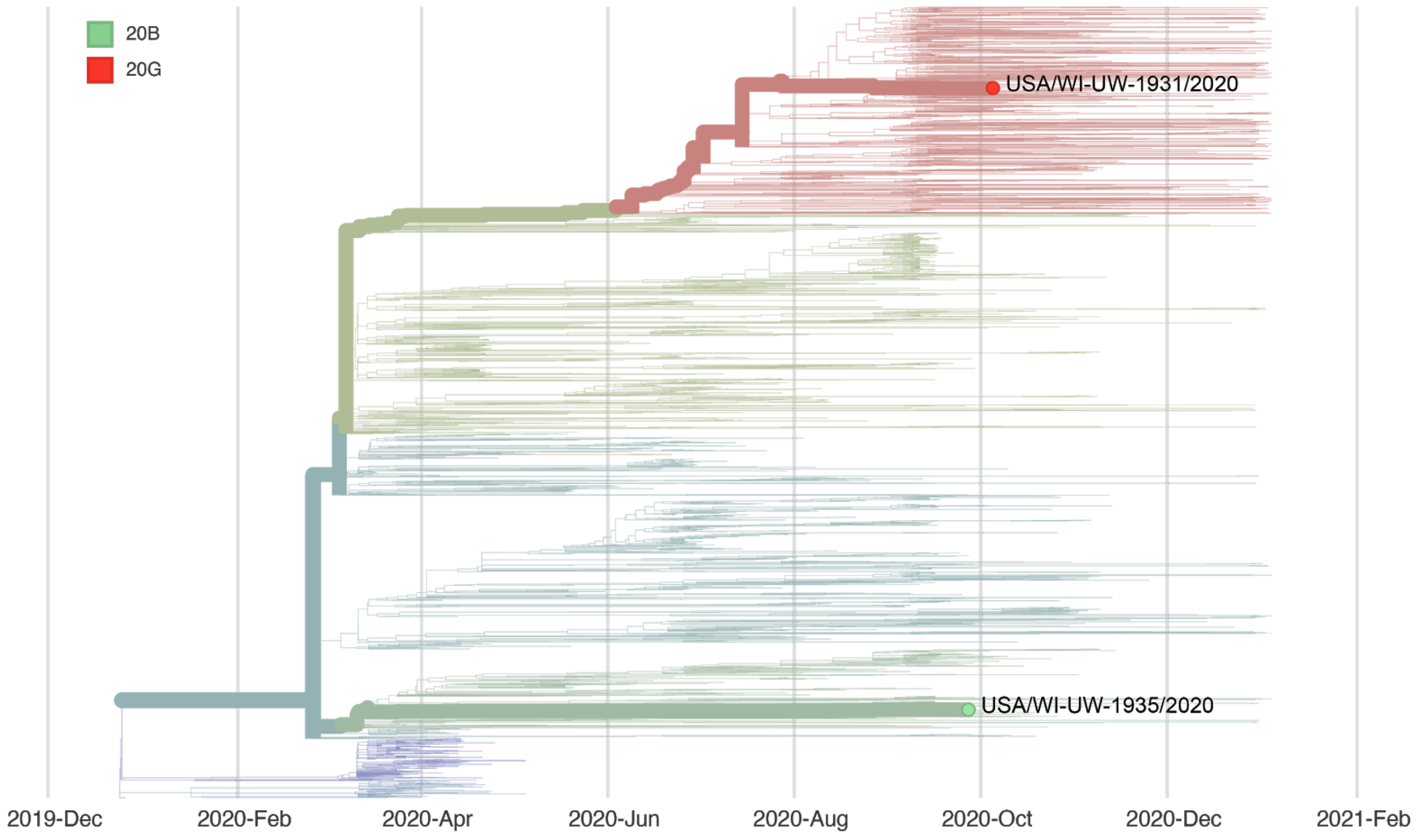

### Report #20. 2020-11-05.

#### Likely source of HCP infection

HCP 1. Outside community.

#### Samples

| **Sample type** | **Sample collection date** | **GISAID identifier** | **Clade** (Nextstrain) | **Lineage** (Pangolin) |
|---|---|---|---|---|
| HCP 1 | October 2020 | hCoV-19/USA/WI-UW-1931/2020 | 20G | B.1.2 |
| patient A | September 2020 | hCoV-19/USA/WI-UW-1935/2020 | 20A | B.1 |

#### Epidemiological information

In the two weeks before symptom onset, HCP 1 provided direct care to patient A. HCP 1 wore appropriate PPE while providing care and reported no lapses in PPE use.

#### Alignment

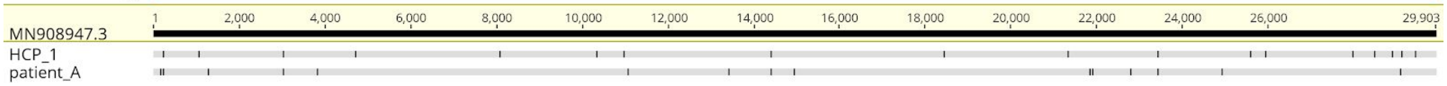


#### Phylogeny

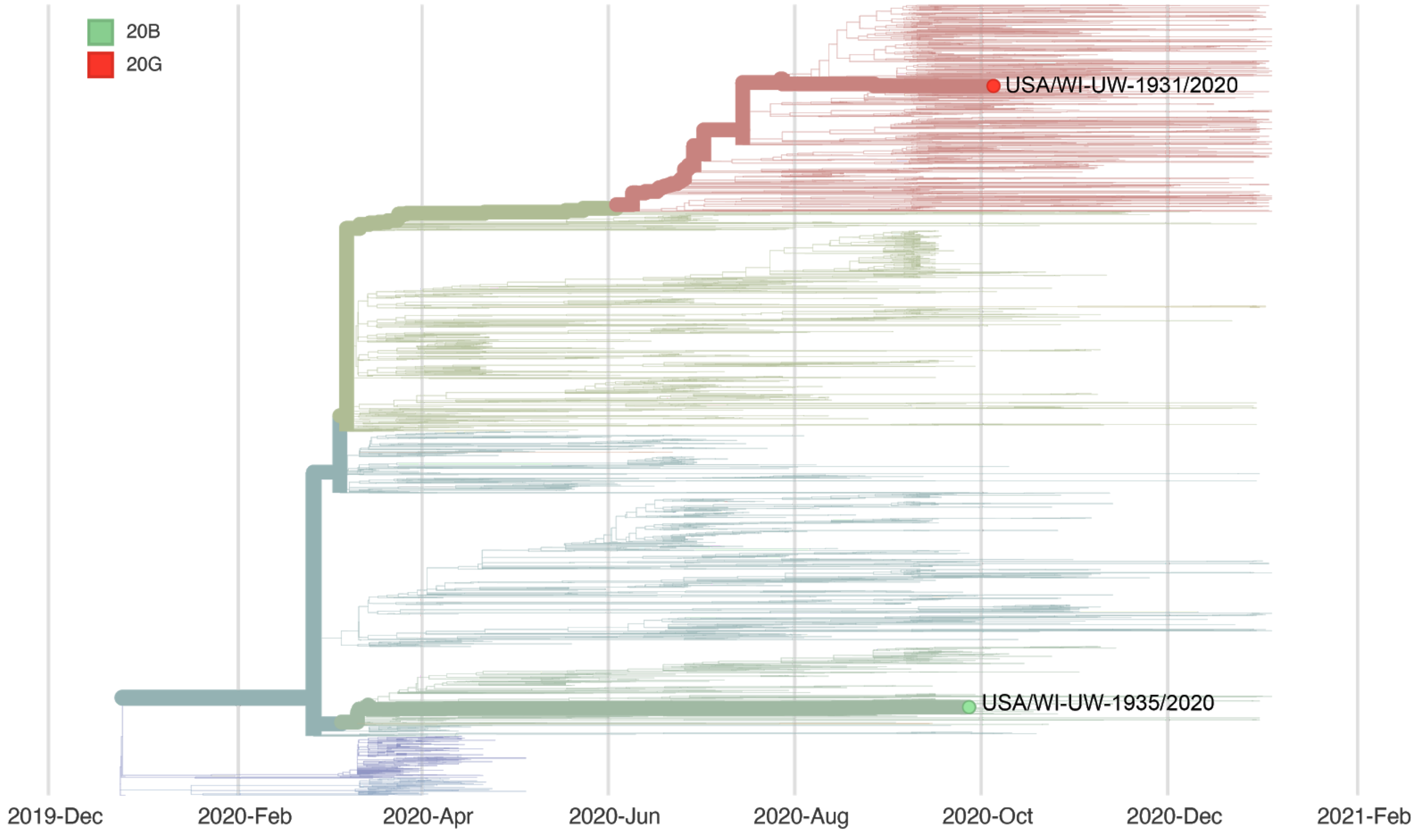

### Report #21. 2020-11-05.

#### Likely source of HCP infection

HCP 1. Outside community.
HCP 2. Outside community.

#### Samples

| Sample type | Sample collection date | GISAID identifier | Clade (Nextstrain) | Lineage (Pangolin) |
|---|---|---|---|---|
| HCP 1 | October 2020 | hCoV-19/USA/WI-UW-1929/2020 | 20G | B.1.2 |
| HCP 2 | October 2020 | hCoV-19/USA/WI-UW-1938/2020 | 20B | B.1.1.73 |
| patient A | September 2020 | N/A - no consensus sequence | | |
| patient B | September 2020 | N/A - no consensus sequence | | |
| patient C | September 2020 | hCoV-19/USA/WI-UW-1934/2020 | 20G | B.1.2 |
| patient D | September 2020 | hCoV-19/USA/IL-UW-1937/2020 | 20G | B.1.2 |
| patient E | October 2020 | N/A - no consensus sequence | | |

#### Epidemiological information

HCP 1 and 2 worked in the same department and both provided direct patient care to patients A-E in the two weeks before their onset of symptoms. HCP 1 and HCP 2 reported no lapses in PPE with each other or with their patients.

#### Alignment

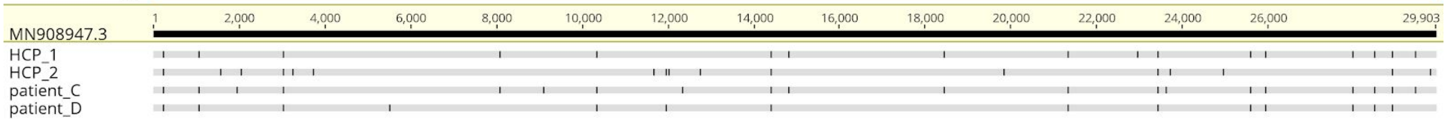

#### Phylogeny

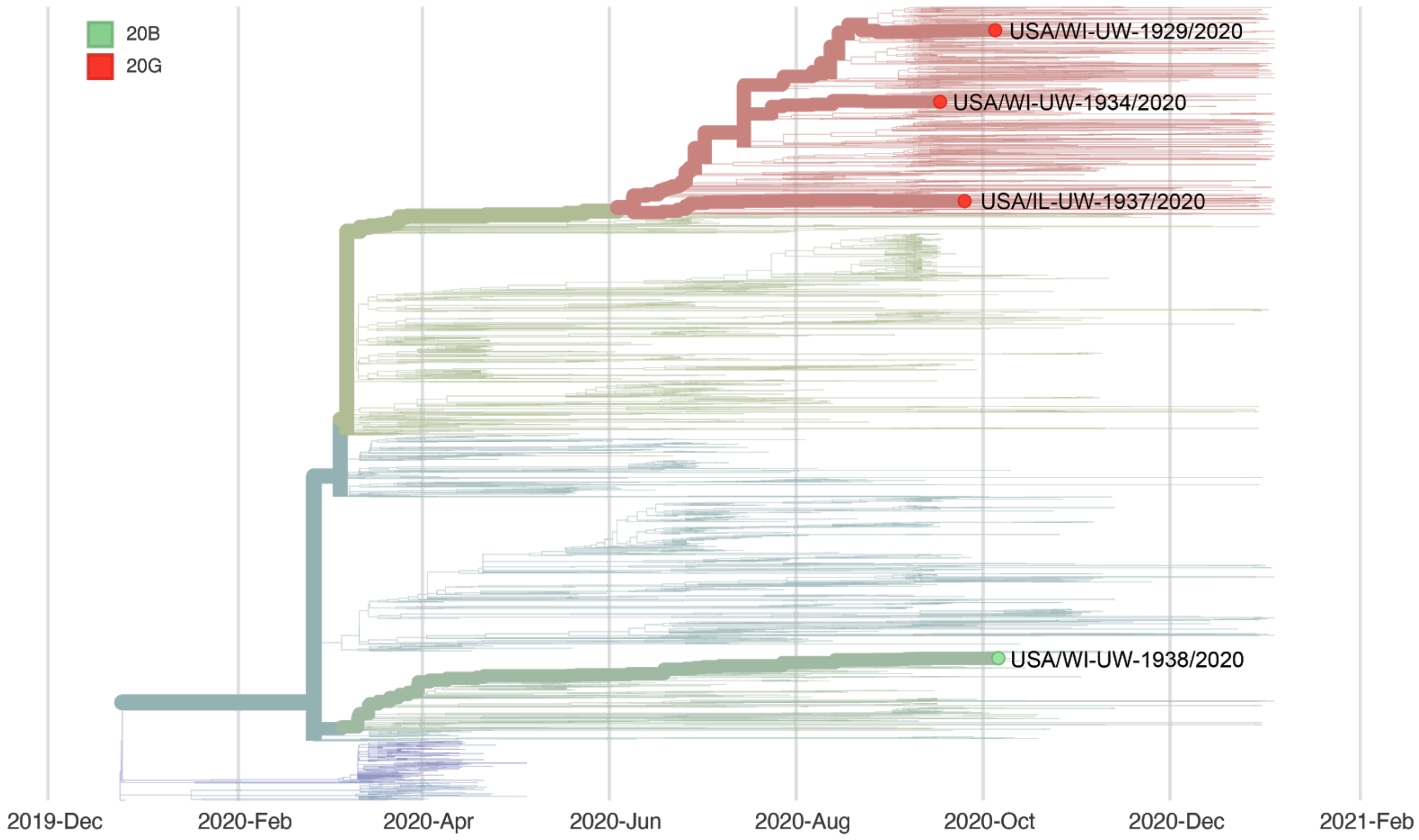


### Report #22. 2020-11-05.

#### Likely source of HCP infection

HCP 1. Outside community.

#### Samples

| **Sample type** | **Sample collection date** | **GISAID identifier** | **Clade** (Nextstrain) | **Lineage** (Pangolin) |
|---|---|---|---|---|
| HCP 1 | October 2020 | hCoV-19/USA/WI-UW-1936/2020 | 20G | B.1.2 |
| patient A | July 2020 | hCoV-19/USA/WI-UW-1100/2020 | 20A | B.1.139 |

#### Epidemiological information

HCP 1 performed a postmortem examination on patient A. At the time the patient expired, they were known to have an active COVID-19 infection.

#### Alignment

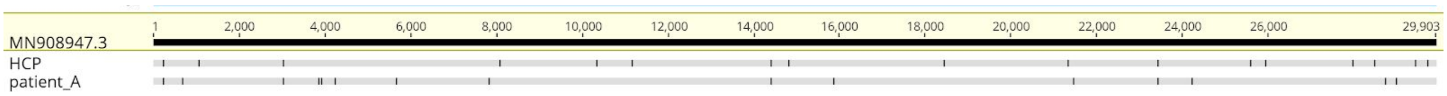


#### Phylogeny

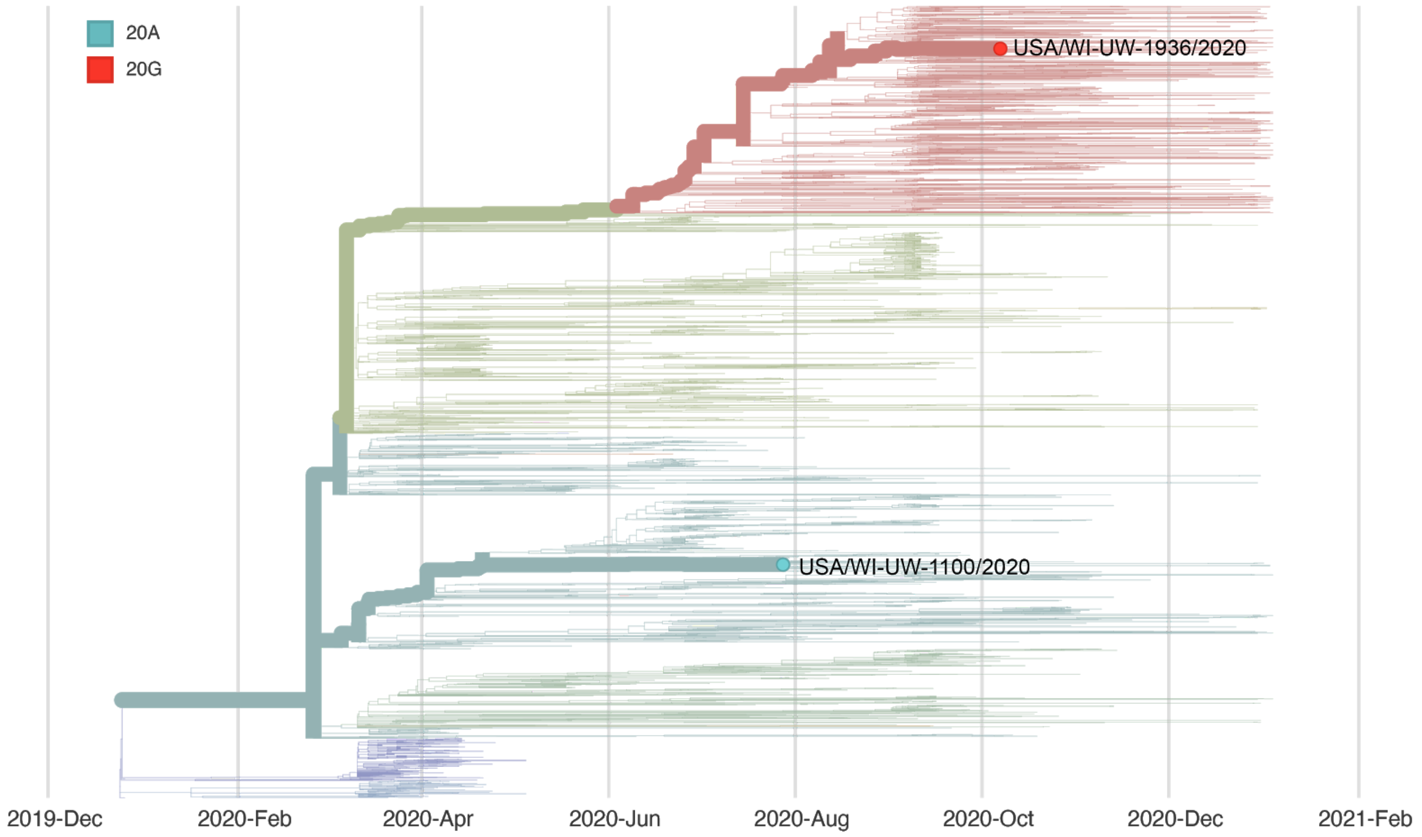


### Report #23. 2020-11-05.

#### Likely source of HCP infection

HCP 1. Outside community (household contact).
HCP 2. Employee source.
HCP 3. Employee source.
HCP 4. Employee source.
HCP 5. Outside community.

#### Samples

| **Sample type** | **Sample collection date** | **GISAID identifier** | **Clade** (Nextstrain) | **Lineage** (Pangolin) |
|---|---|---|---|---|
| HCP 1 | October 2020 | hCoV-19/USA/WI-UW-1933/2020 | 20G | B.1.2 |
| HCP 2 | October 2020 | hCoV-19/USA/WI-UW-1926/2020 | 20G | B.1.2 |

| | | | | |
|---|---|---|---|---|
| HCP 3 | October 2020 | hCoV-19/USA/WI-UW-1939/2020 | 20G | B.1.2 |
| HCP 4 | October 2020 | hCoV-19/USA/WI-UW-1927/2020 | 20G | B.1.2 |
| HCP 5 | October 2020 | hCoV-19/USA/WI-UW-1932/2020 | 20A | B.1.139 |

#### Epidemiological information

All of these HCP work in the same department, with the exception of HCP 1 who is a household contact of HCP 2. HCP 2-5 reported sharing an unmasked meal together prior to testing positive.

#### Alignment

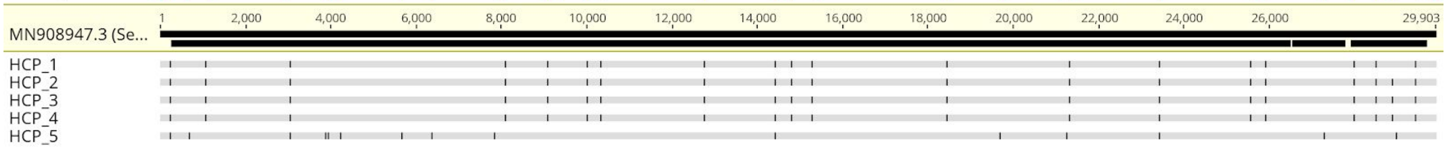


#### Phylogeny

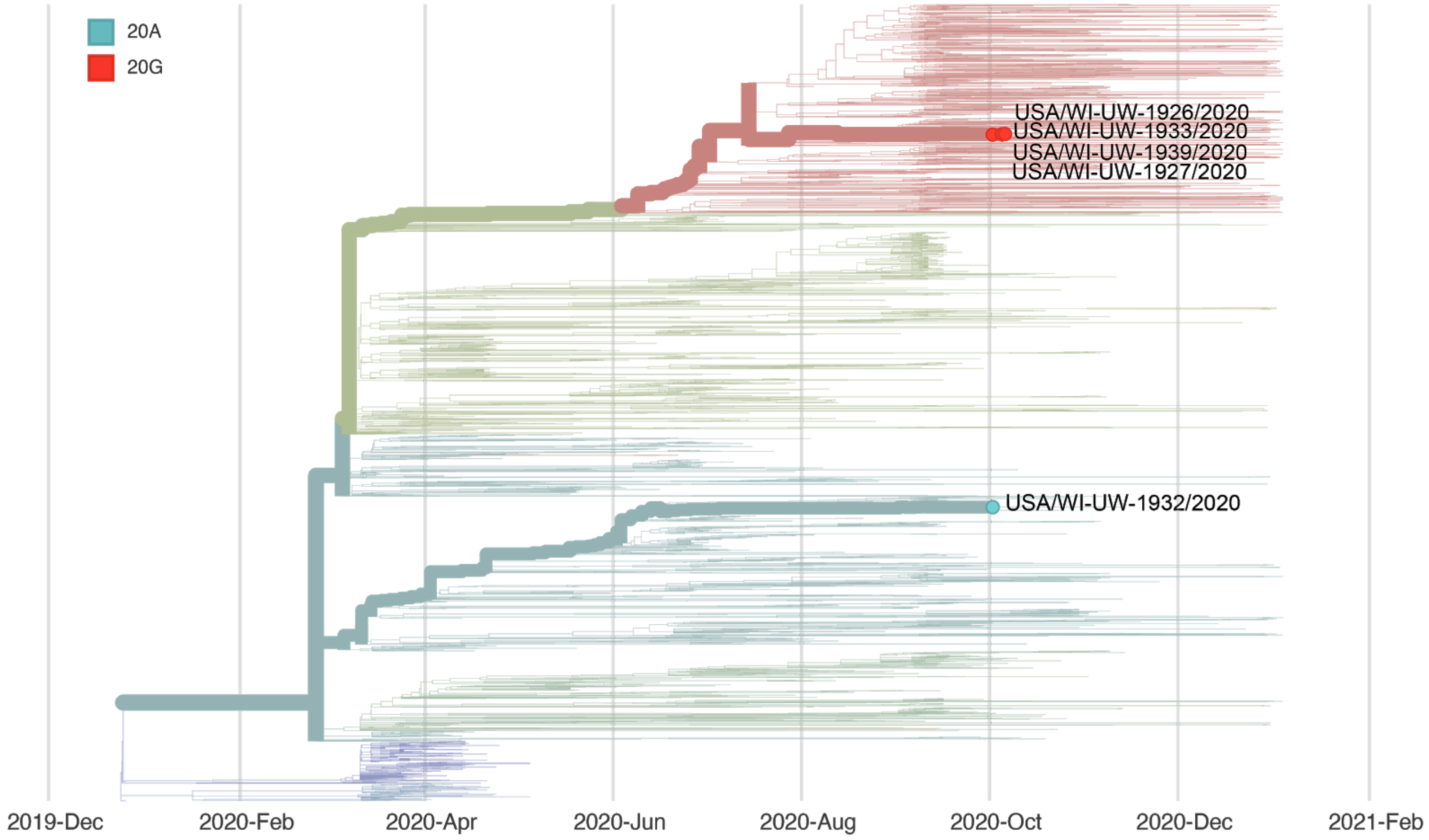


### Report #25. 2020-12.08.

#### Likely source of HCP infection

HCP 1. Patient source (patient A).

#### Samples

| **Sample type** | **Sample collection date** | **GISAID identifier** | **Clade** (Nextstrain) | **Lineage** (Pangolin) |
|---|---|---|---|---|
| HCP 1 | October 2020 | hCoV-19/USA/WI-UW-2326/2020 | 20C | B.1.2 |
| patient A | October 2020 | hCoV-19/USA/WI-UW-2362/2020 | 20C | B.1.2 |
| patient B | October 2020 | hCoV-19/USA/WI-UW-2389/2020 | 20C | B.1.2 |

#### Epidemiological information

In the two weeks before symptom onset, HCP 1 provided direct care to patients A and B.

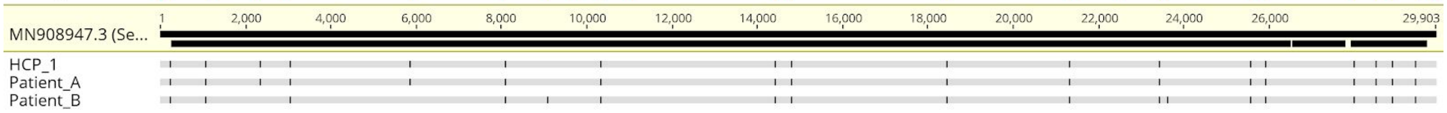


#### Alignment

#### Phylogeny

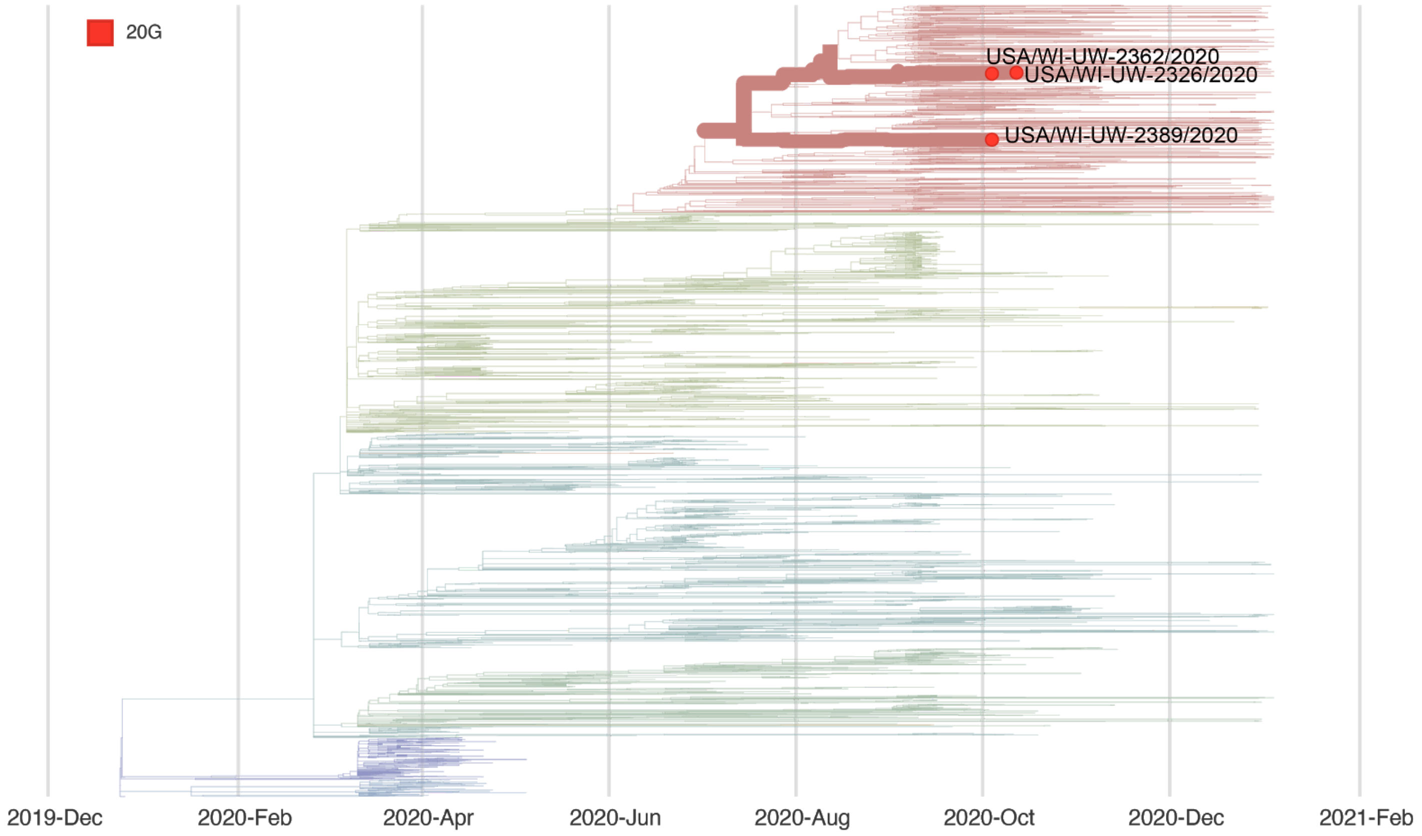

### Report #27. 2020-12.09.

#### Likely source of HCP infection

HCP 1. Outside community.
HCP 2. Outside community.
HCP 3. Outside community.
HCP 4. Outside community.

#### Samples

| **Sample type** | **Sample collection date** | **GISAID identifier** | **Clade** (Nextstrain) | **Lineage** (Pangolin) |
|---|---|---|---|---|
| HCP 1 | October 2020 | hCoV-19/USA/WI-UW-2226/2020 | 20A | B.1 |
| HCP 2 | October 2020 | N/A | 20G | B.1.2 |
| HCP 3 | October 2020 | hCoV-19/USA/WI-UW-2227/2020 | 20G | B.1.2 |
| HCP 4 | October 2020 | hCoV-19/USA/WI-UW-2228/2020 | 20G | B.1.2 |
| patient A | September 2020 | hCov-19/USA/IL-UW-2327/2020 | 20A | B.1 |
| patient B | September 2020 | hCoV-19/USA/WI-UW-1972/2020 | 20G | B.1 |

#### Epidemiological information

HCP 1-4 work in the same department. HCP 1-4 all had direct interactions with patients A and/or B during the 14 days prior to symptom onset.

#### Alignment

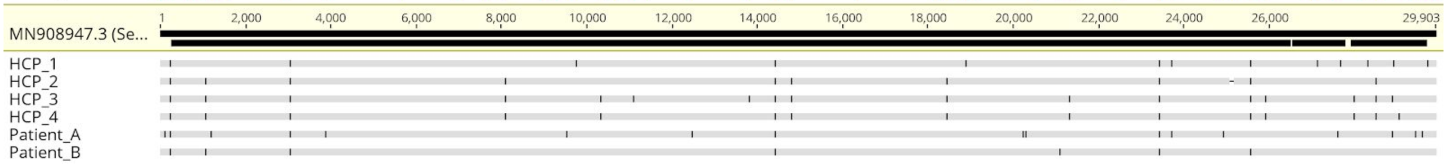

#### Phylogeny

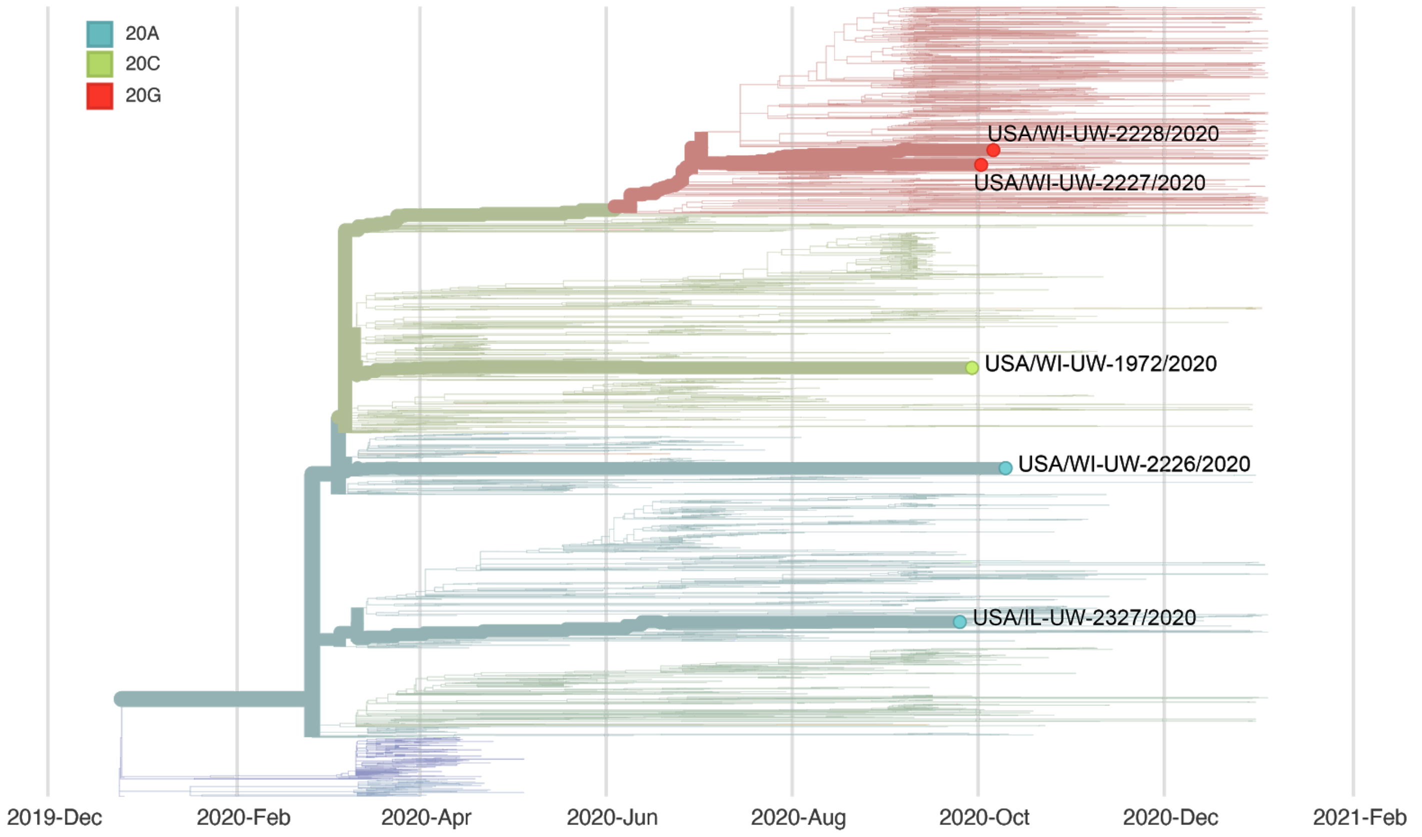


### Report #28. 2020-12.09.

#### Likely source of HCP infection

HCP 1. Outside community.
HCP 2. Outside community.
HCP 3. Outside community.

#### Samples

| **Sample type** | **Sample collection date** | **GISAID identifier** | **Clade** (Nextstrain) | **Lineage** (Pangolin) |
|---|---|---|---|---|
| Patient A | November 2020 | hCoV-19/USA/WI-UW-2265/2020 | 20G | B.1.2 |
| HCP 1 | October 2020 | hCoV-19/USA/WI-UW-2266/2020 | 20G | B.1.2 |
| HCP 2 | October 2020 | hCoV-19/USA/WI-UW-2267/2020 | 20G | B.1.2 |
| HCP 3 | October 2020 | hCoV-19/USA/WI-UW-2268/2020 | 20G | B.1.2 |

#### Epidemiological information

HCP 1-3 provided direct patient care to patient A in the 14 days before symptom onset. All HCP reported appropriate use of PPE with no lapses. HCP 1 and 2 had no known interactions prior to their infections. HCP 2 and HCP 3 both attended a high-risk community event for greater than 15 minutes together.

#### Alignment

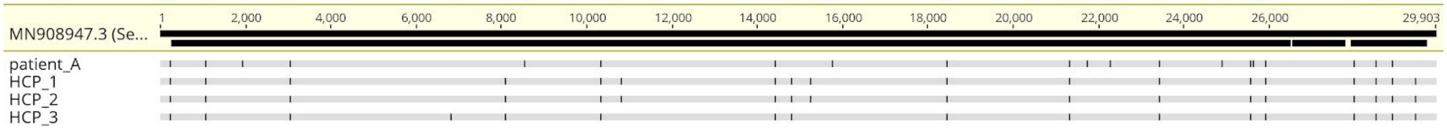


#### Phylogeny

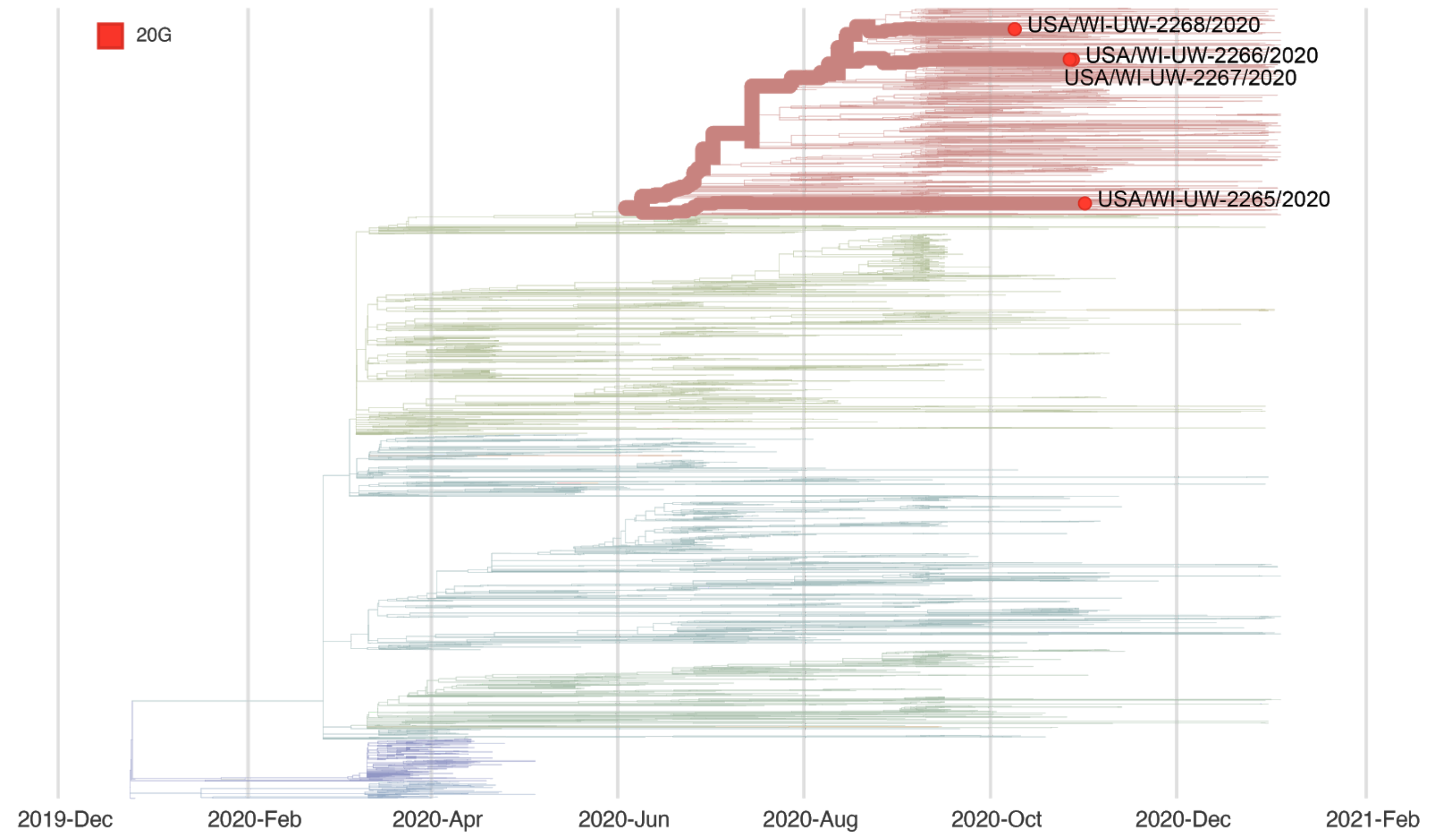


### Report #29. 2020-12.09.

#### Likely source of HCP infection

HCP 1. Employee source (HCP 3).
HCP 2. Outside community.
HCP 3. Outside community.

#### Samples

| **Sample type** | **Sample collection date** | **GISAID identifier** | **Clade** (Nextstrain) | **Lineage** (Pangolin) |
|---|---|---|---|---|
| HCP 1 | November 2020 | hCoV-19/USA/WI-UW-2269/2020 | 20G | B.1.2 |
| HCP 2 | November 2020 | hCoV-19/USA/WI-UW-2270/2020 | 20G | B.1.2 |
| HCP 3 | November 2020 | hCoV-19/USA/WI-UW-2271/2020 | 20G | B.1.2 |

#### Epidemiological information

HCP 1-3 work in the same department. HCP 1 and 3 attended an in person meeting together and reported sitting 6-feet apart while wearing masks. HCP 2 did not attend this meeting. HCP 3 reported symptoms before HCP 1.

#### Alignment

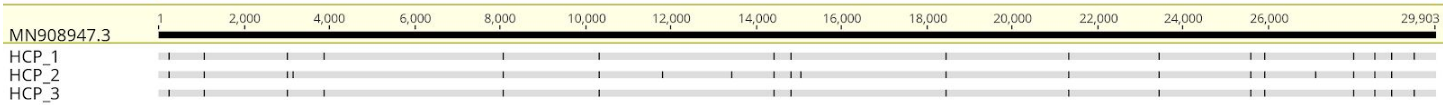


#### Phylogeny

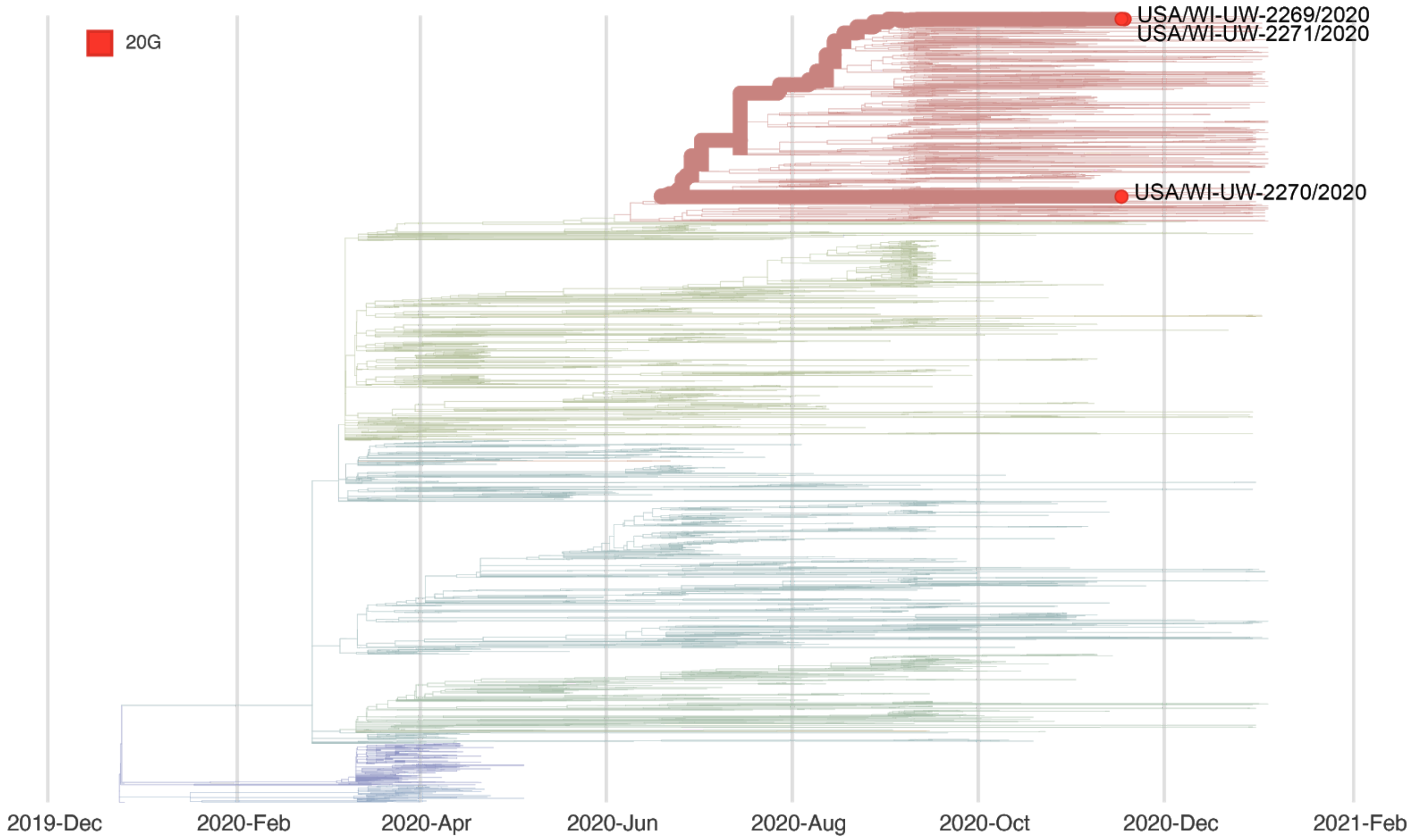

### Report #30. 2020-12.09.

#### Likely source of HCP infection

HCP 1. Patient source (patient C).

#### Samples

| Sample type | Sample collection date | GISAID identifier | Clade (Nextstrain) | Lineage (Pangolin) |
|---|---|---|---|---|
| HCP 1 | November 2020 | hCoV-19/USA/WI-UW-2272/2020 | 20G | B.1.2 |
| patient A | November 2020 | hCov-19/USA/WI-UW-2324/2020 | 20G | B.1.2 |
| patient B | October 2020 | hCoV-19/USA/WI-UW-2273/2020 | 20G | B.1.2 |
| patient C | November 2020 | hCoV-19/USA/WI-UW-2274/2020 | 20G | B.1.2 |
| patient D | November 2020 | hCoV-19/USA/WI-UW-2275/2020 | 20A | B.1.139 |
| patient E | November 2020 | hCoV-19/USA/WI-UW-2276/2020 | 20G | B.1.2 |

#### Epidemiological information

HCP 1 provided direct patient care to patients A-E and reported no lapses in PPE use.

#### Alignment

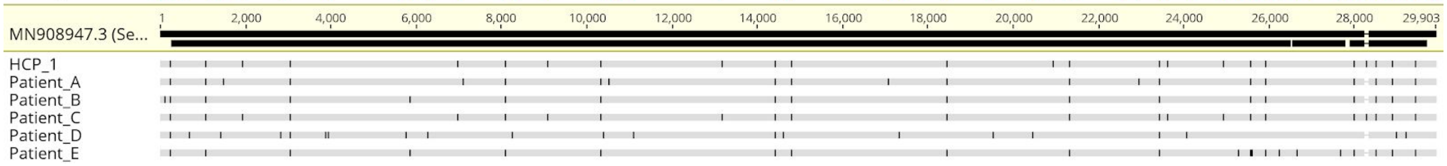

#### Phylogeny

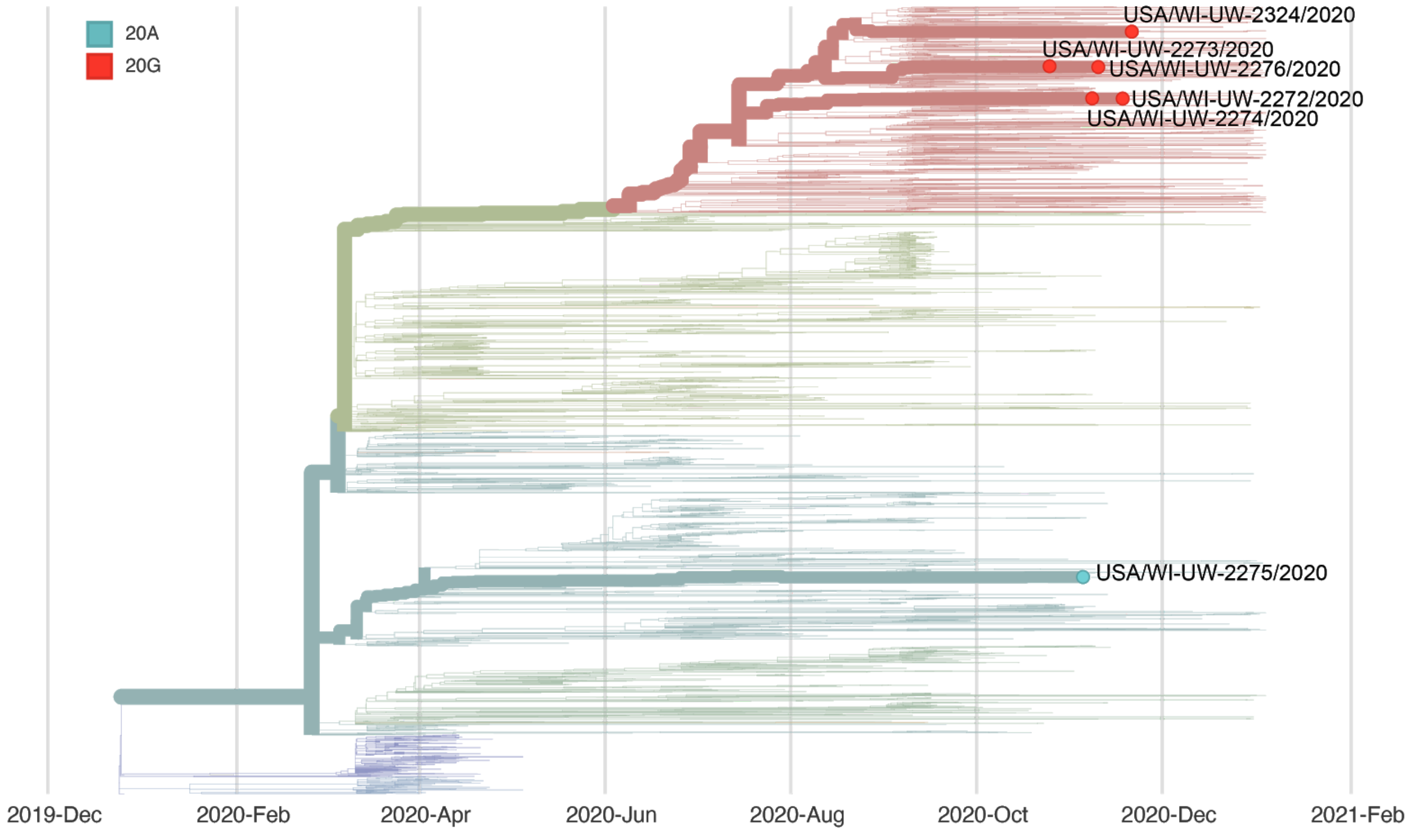


### Report #31. 2020-12.09.

#### Likely source of HCP infection

HCP 1. Outside community.
HCP 2. Outside community.
HCP 3. Outside community.
HCP 4. Employee source (HCP 2).
HCP 5. Employee source (HCP 2).
HCP 6. Outside community.
HCP 7. Outside community.

#### Samples

| **Sample type** | **Sample collection date** | **GISAID identifier** | **Clade** (Nextstrain) | **Lineage** (Pangolin) |
|---|---|---|---|---|
| HCP 1 | November 2020 | hCoV-19/USA/WI-UW-2312/2020 | 20C | B.1.363 |
| HCP 2 | November 2020 | hCoV-19/USA/WI-UW-2277/2020 | 20G | B.1.2 |
| HCP 3 | November 2020 | hCoV-19/USA/WI-UW-2278/2020 | 20G | B.1.2 |
| HCP 4 | November 2020 | hCoV-19/USA/WI-UW-2279/2020 | 20G | B.1.2 |

| | | | | |
|---|---|---|---|---|
| HCP 5 | November 2020 | hCoV-19/USA/WI-UW-2280/2020 | 20G | B.1.2 |
| HCP 6 | November 2020 | hCoV-19/USA/WI-UW-2281/2020 | 20G | B.1.2 |
| HCP 7 | November 2020 | hCoV-19/USA/WI-UW-2282/2020 | 20G | B.1.2 |

##### Epidemiological information

These HCP are all in the same department. Their level of interaction with each other is unclear.

##### Alignment

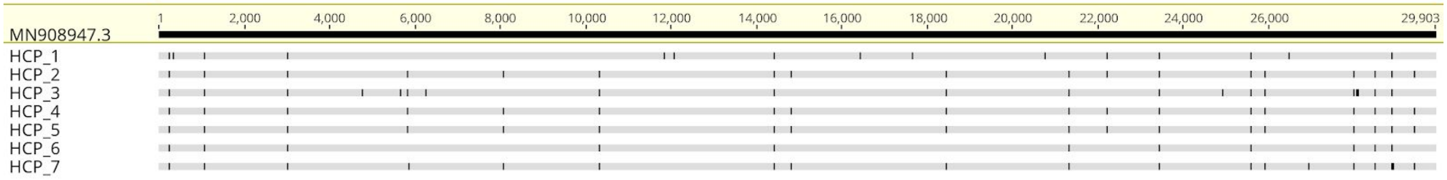


##### Phylogeny

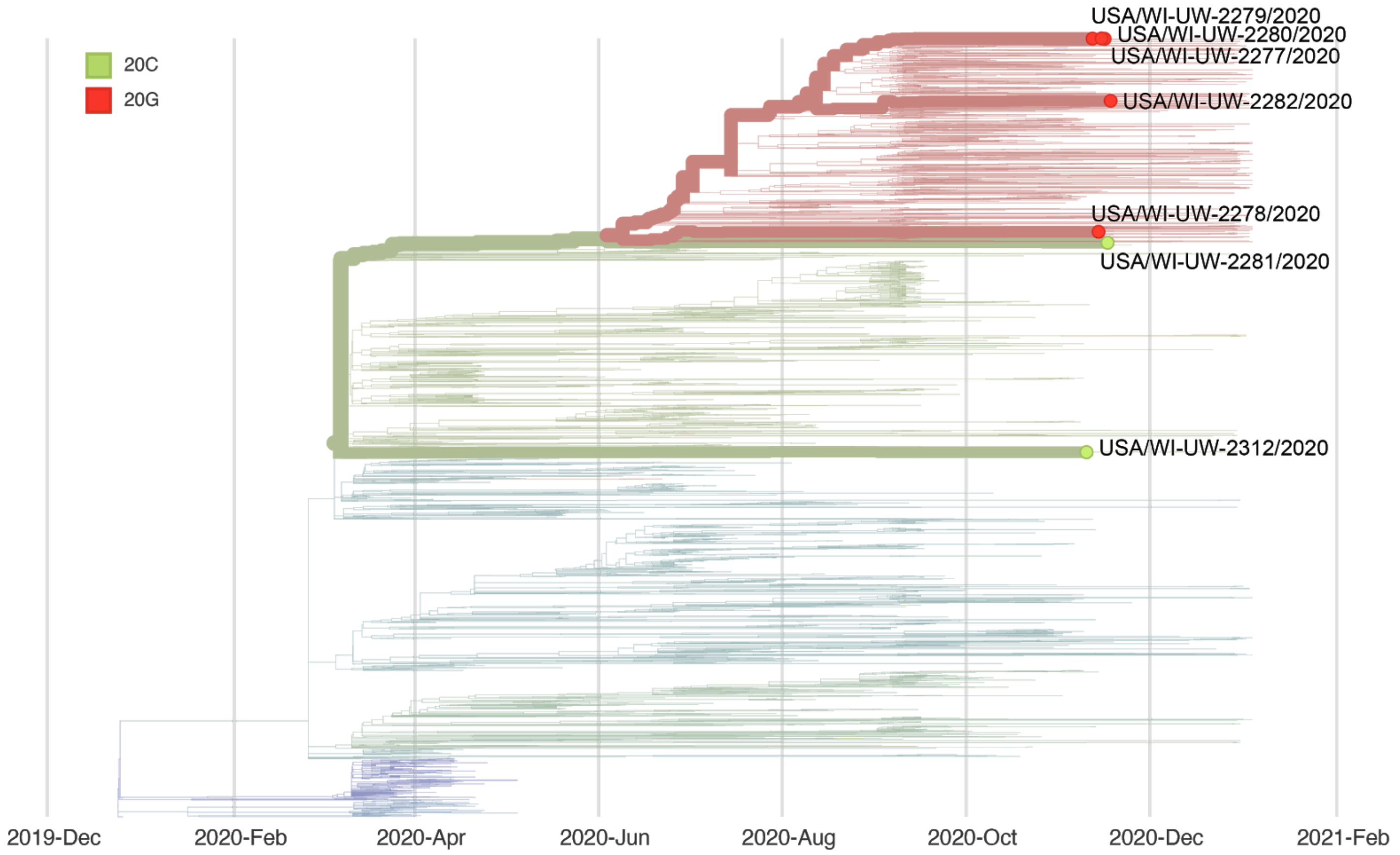


#### Report #32. 2020-12.09.

##### Likely source of HCP infection

HCP 1. Outside community.
HCP 2. Outside community.

HCP 3. Outside community.
HCP 4. Outside community.
HCP 5. Outside community.
HCP 6. Outside community.
HCP 7. Outside community.
HCP 8. Outside community.
HCP 9. Outside community.
HCP 10. Outside community.
HCP 11. Outside community.
HCP 12. Outside community.
HCP 13. Inconclusive (could be HCP 8, but these samples were collected >14 apart).
HCP 14. Employee source (HCP 8 or HCP 13).

#### Samples

| **Sample type** | **Sample collection date** | **GISAID identifier** | **Clade** (Nextstrain) | **Lineage** (Pangolin) |
|---|---|---|---|---|
| HCP 1 | October 2020 | hCoV-19/USA/WI-UW-2283/2020 | 20G | B.1.370 |
| HCP 2 | October 2020 | hCoV-19/USA/WI-UW-2284/2020 | 20G | B.1.2 |
| HCP 3 | October 2020 | hCoV-19/USA/WI-UW-2285/2020 | 20A | B.1.216 |
| HCP 4 | October 2020 | hCoV-19/USA/WI-UW-2286/2020 | 20G | B.1.2 |
| HCP 5 | October 2020 | hCoV-19/USA/WI-UW-2390/2020 | 20G | B.1.2 |
| HCP 6 | October 2020 | hCoV-19/USA/WI-UW-2391/2020 | 20G | B.1 |
| HCP 7 | November 2020 | hCoV-19/USA/WI-UW-2287/2020 | 20G | B.1.2 |
| HCP 8 | November 2020 | hCoV-19/USA/WI-UW-2288/2020 | 20G | B.1.2 |
| HCP 9 | November 2020 | hCoV-19/USA/WI-UW-2289/2020 | 20G | B.1.2 |
| HCP 10 | November 2020 | hCoV-19/USA/WI-UW-2313/2020 | 20G | B.1.2 |
| HCP 11 | November 2020 | hCoV-19/USA/WI-UW-2290/2020 | 20G | B.1.2 |
| HCP 12 | November 2020 | hCoV-19/USA/WI-UW-2291/2020 | 20G | B.1.2 |
| HCP 13 | November 2020 | hCoV-19/USA/WI-UW-2292/2020 | 20G | B.1.2 |
| HCP 14 | November 2020 | hCoV-19/USA/WI-UW-2293/2020 | 20G | B.1.2 |
| patient A | November 2020 | hCoV-19/USA/WI-UW-2325/2020 | 20B | B.1.1.73 |

#### Epidemiological information

HCP 1-14 work in the same department together. One or more of these HCP provided direct patient care to patient A.

#### Alignment

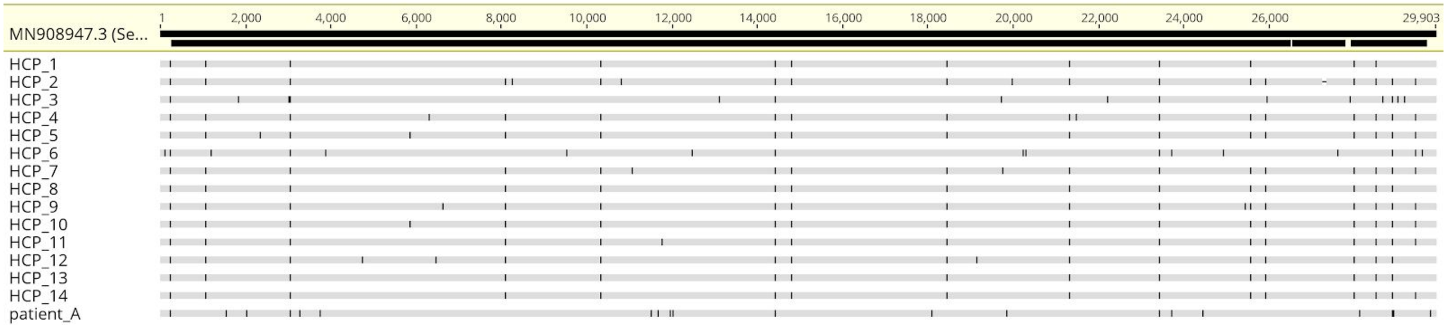


#### Phylogeny

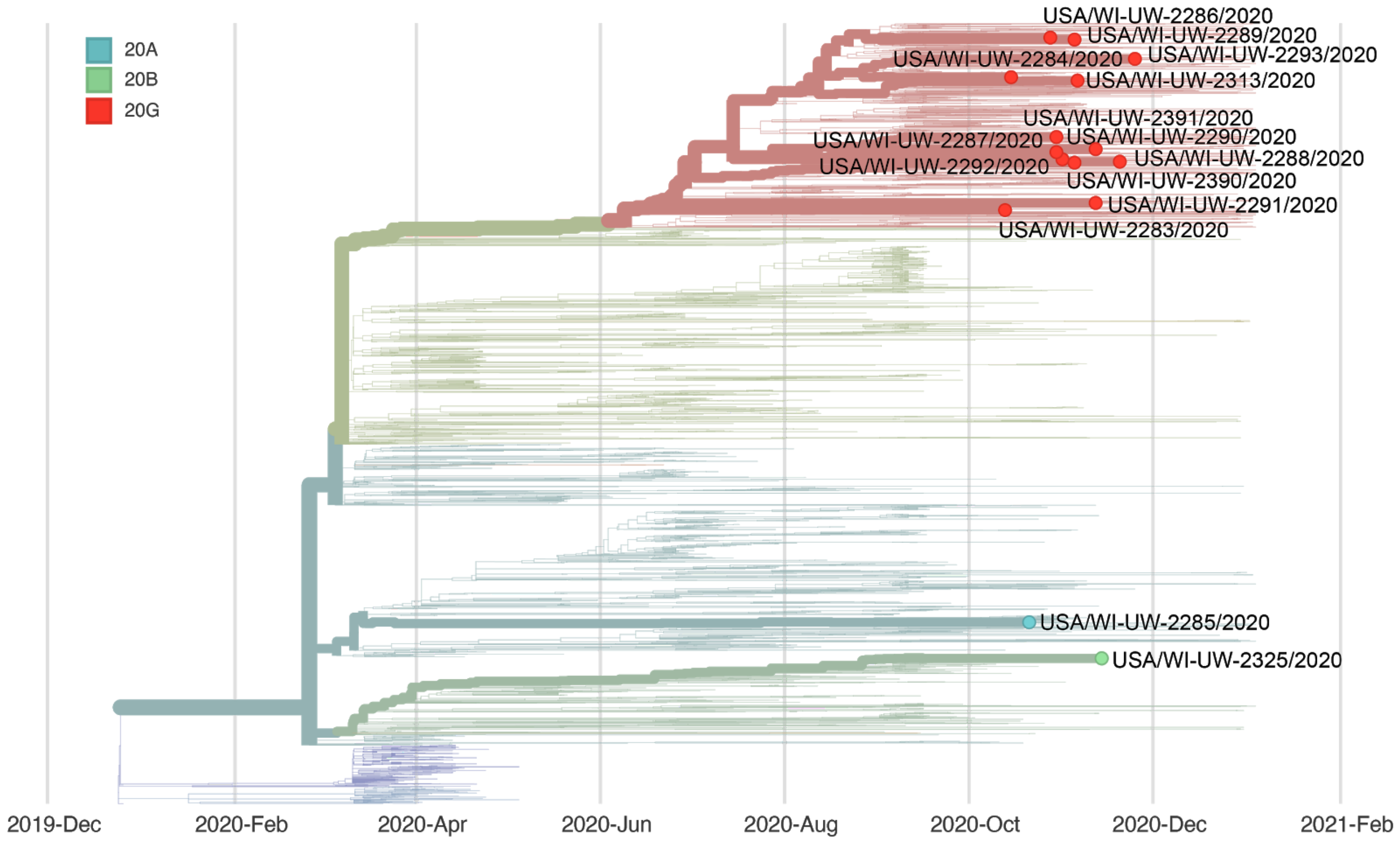


### Report #33. 2021-01-03.

#### Likely source of HCP infection

HCP 1. Combined patient and employee cluster.
HCP 2. Combined patient and employee cluster.
HCP 3. Combined patient and employee cluster.
HCP 4. Outside community.
HCP 5. Combined patient and employee cluster.

Patients A, B and C are the patients involved in this combined patient and employee cluster.

#### Samples

| **Sample type** | **Sample collection date** | **GISAID identifier** | **Clade** (Nextstrain) | **Lineage** (Pangolin) |
|---|---|---|---|---|
| HCP 1 | December 2020 | hCoV-19/USA/WI-UW-2394/2020 | 20G | B.1.2 |
| HCP 2 | December 2020 | hCoV-19/USA/WI-UW-2396/2020 | 20G | B.1.2 |
| HCP 3 | December 2020 | hCoV-19/USA/WI-UW-2451/2020 | 20G | B.1.2 |
| HCP 4 | December 2020 | hCoV-19/USA/WI-UW-2452/2020 | 20G | B.1.2 |
| HCP 5 | December 2020 | hCoV-19/USA/WI-UW-2450/2020 | 20G | B.1.2 |
| patient A | December 2020 | hCoV-19/USA/WI-UW-2392/2020 | 20G | B.1.2 |
| patient B | December 2020 | hCoV-19/USA/WI-UW-2393/2020 | 20G | B.1.2 |
| patient C | December 2020 | hCoV-19/USA/IL-UW-2449/2020 | 20G | B.1.2 |

#### Epidemiological information

HCP 1-5 provided direct care to one or more of these patients, A-C. HCP 1-5 may have also interacted with each other. No lapses in PPE were reported. Patient A had the earliest reported symptom onset.

#### Alignment

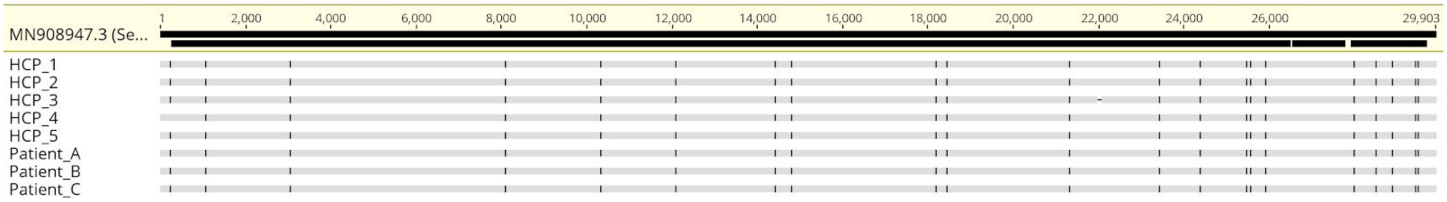

#### Phylogeny

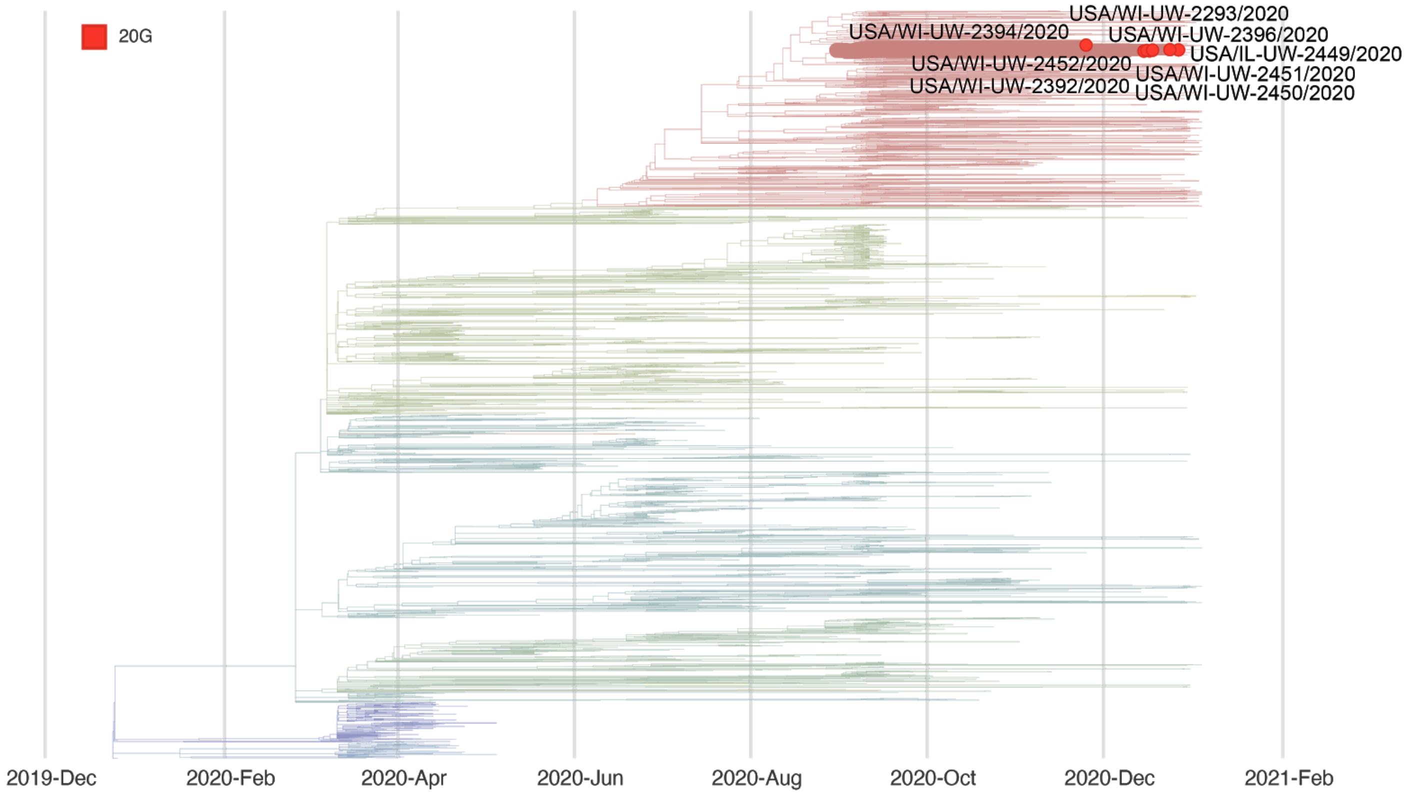


### Report #34. 2021-01-03.

#### Likely source of HCP infection

HCP 1. Outside community.

#### Samples

| **Sample type** | **Sample collection date** | **GISAID identifier** | **Clade** (Nextstrain) | **Lineage** (Pangolin) |
|---|---|---|---|---|
| HCP 1 | December 2020 | hCoV-19/USA/WI-UW-2453/2020 | 20G | B.1.2 |
| patient A | December 2020 | hCoV-19/USA/WI-UW-2395/2020 | 20G | B.1.2 |

#### Epidemiological information

HCP 1 provided direct care to patient A during the 14 days prior to HCP 1's symptom onset. HCP 1 reported no lapse in PPE when providing care to patient A.

#### Alignment

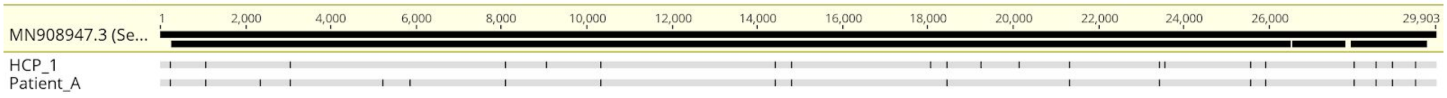

#### Phylogeny

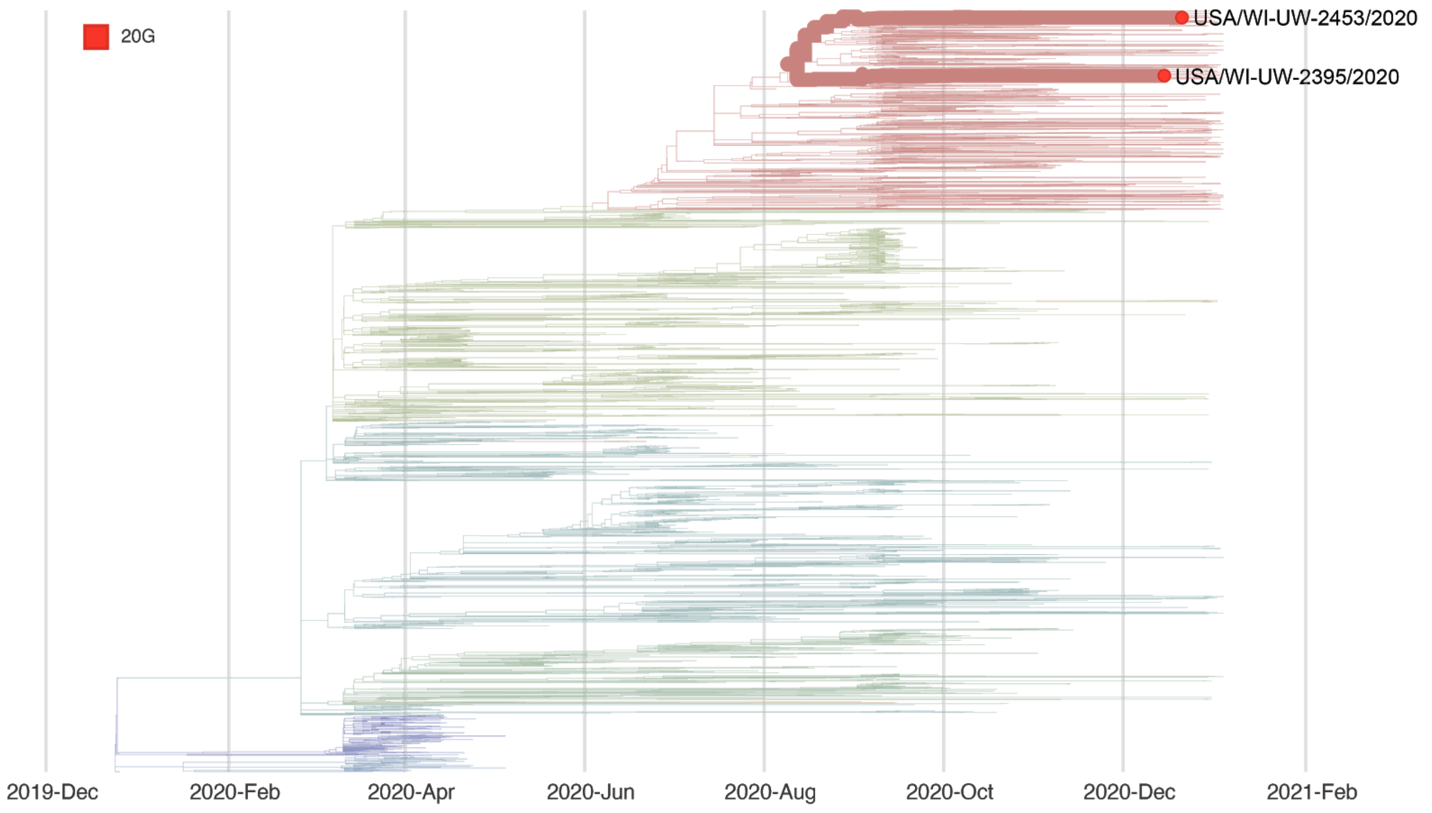

20G
USA/WI-UW-2453/2020
USA/WI-UW-2395/2020
2019-Dec
2020-Feb
2020-Apr
2020-Jun
2020-Aug
2020-Oct
2020-Dec
2021-Feb
